# Integrated multiomics implicates dysregulation of ECM and cell adhesion pathways as drivers of severe COVID-associated kidney injury

**DOI:** 10.1101/2024.03.18.24304401

**Authors:** Nanditha Anandakrishnan, Zhengzi Yi, Zeguo Sun, Tong Liu, Jonathan Haydak, Sean Eddy, Pushkala Jayaraman, Stefanie DeFronzo, Aparna Saha, Qian Sun, Dai Yang, Anthony Mendoza, Gohar Mosoyan, Huei Hsun Wen, Jennifer A. Schaub, Jia Fu, Thomas Kehrer, Rajasree Menon, Edgar A. Otto, Bradley Godfrey, Mayte Suarez-Farinas, Sean Leffters, Akosua Twumasi, Kristin Meliambro, Alexander W. Charney, Adolfo García-Sastre, Kirk N. Campbell, G. Luca Gusella, John Cijiang He, Lisa Miorin, Girish N. Nadkarni, Juan Wisnivesky, Hong Li, Matthias Kretzler, Steve G. Coca, Lili Chan, Weijia Zhang, Evren U. Azeloglu

**Affiliations:** Department of Medicine, Division of Nephrology, Icahn School of Medicine at Mount Sinai, New York, NY; Center for Advanced Proteomics Research, Rutgers University, Newark, New Jersey; Department of Internal Medicine, Division of Nephrology, University of Michigan, Ann Arbor, Michigan, USA; The Charles Bronfman Institute for Personalized Medicine, Icahn School of Medicine at Mount Sinai, New York, NY, USA; Department of Microbiology, Icahn School of Medicine at Mount Sinai, New York, NY 10029, USA; Department of Computational Medicine and Bioinformatics, University of Michigan, Ann Arbor, Michigan, USA; Department of Genetics and Genomic Sciences, Icahn School of Medicine at Mount Sinai, New York, New York, USA; Center for Biostatistics, Department of Population Health Science and Policy, Icahn School of Medicine at Mount Sinai, New York, New York, USA; Division of General Internal Medicine, Icahn School of Medicine at Mount Sinai, One Gustave L. Levy Place, New York, NY 10029, USA; Pamela Sklar Division of Psychiatric Genomics, Icahn School of Medicine at Mount Sinai, New York, New York, USA; Global Health and Emerging Pathogens Institute, Icahn School of Medicine at Mount Sinai, New York, NY 10029, USA; Department of Medicine, Division of Infectious Diseases, Icahn School of Medicine at Mount Sinai, New York, NY 10029, USA; Tisch Cancer Institute, Icahn School of Medicine at Mount Sinai, New York, NY 10029, USA; Department of Pathology, Molecular, and Cell-Based Medicine, Icahn School of Medicine at Mount Sinai, New York, NY 10029, USA; The Icahn Genomics Institute, Icahn School of Medicine at Mount Sinai, New York, NY 10029, USA; Renal Program, James J. Peters Veterans Affairs Medical Center at Bronx, Bronx, NY; Department of Medicine, Division of Data-Driven and Digital Medicine, Icahn School of Medicine at Mount Sinai, New York, NY; Department of Pharmacological Sciences, Icahn School of Medicine at Mount Sinai, New York, NY, USA; Black Family Stem Cell Institute, Icahn School of Medicine at Mount Sinai, New York, NY, 10029, USA

**Keywords:** machine learning, COVID-19, urine proteomics, multiomics, kidney organoids

## Abstract

COVID-19 has been a significant public health concern for the last four years; however, little is known about the mechanisms that lead to severe COVID-associated kidney injury. In this multicenter study, we combined quantitative deep urinary proteomics and machine learning to predict severe acute outcomes in hospitalized COVID-19 patients. Using a 10-fold cross-validated random forest algorithm, we identified a set of urinary proteins that demonstrated predictive power for both discovery and validation set with 87% and 79% accuracy, respectively. These predictive urinary biomarkers were recapitulated in non-COVID acute kidney injury revealing overlapping injury mechanisms. We further combined orthogonal multiomics datasets to understand the mechanisms that drive severe COVID-associated kidney injury. Functional overlap and network analysis of urinary proteomics, plasma proteomics and urine sediment single-cell RNA sequencing showed that extracellular matrix and autophagy-associated pathways were uniquely impacted in severe COVID-19. Differentially abundant proteins associated with these pathways exhibited high expression in cells in the juxtamedullary nephron, endothelial cells, and podocytes, indicating that these kidney cell types could be potential targets. Further, single-cell transcriptomic analysis of kidney organoids infected with SARS-CoV-2 revealed dysregulation of extracellular matrix organization in multiple nephron segments, recapitulating the clinically observed fibrotic response across multiomics datasets. Ligand-receptor interaction analysis of the podocyte and tubule organoid clusters showed significant reduction and loss of interaction between integrins and basement membrane receptors in the infected kidney organoids. Collectively, these data suggest that extracellular matrix degradation and adhesion-associated mechanisms could be a main driver of COVID-associated kidney injury and severe outcomes.

## Introduction

As of March 2024, the World Health Organization has reported over 774 million COVID-19 cases and 7 million deaths worldwide attributed to COVID-19 ^1^. Although COVID-19 is a respiratory illness with a major impact on the lungs, studies have shown that peripheral organs such as the kidneys are also affected ^2–4^. While direct infection of the kidney is still ambiguous ^5–7^, kidney function is highly affected due to COVID-19 infection as evidenced by a higher incidence of acute kidney injury (AKI) and decline in estimated glomerular filtration rate (eGFR) in patients hospitalized with COVID-19 ^8, 9^. More recently, clinical data have shown that post-acute sequelae of COVID (PASC) increases the risk of chronic kidney disease (CKD) in patients recovered from COVID-19 ^10, 11^.

In the last few years, several studies have employed biospecimens such as urine ^9, 12–15^, plasma ^16–18^, and serum ^19, 20^ to predict severity, measure levels of AKI biomarkers, and better understand the pathophysiology of COVID-associated kidney injury. SARS-CoV-2 uses entry receptors such as ACE2 and TMPRSS2 during infection and these markers are expressed in the tubules, podocytes, and glomerular parietal epithelial cells (PEC) in the kidney ^21^. Studies employing induced pluripotent stem cell (iPSC)-derived kidney organoids as an *in vitro* model for SARS-CoV-2 infection have demonstrated direct infection of kidney cell-types through interaction with these receptors ^22–24^. These studies have identified several mechanisms such as tubulointerstitial fibrosis, disrupted renal absorption, and cytokine-induced JAK/STAT/APOL1 signaling to be driving COVID-associated kidney injury. However, there is limited understanding of the impact of COVID-19 on specific kidney cell-types and injury mechanisms associated with these cell types.

In this multicenter study, we leverage urine’s accessibility as a biospecimen and utility in surveying kidney function in combination with machine learning (ML) and quantitative tandem-mass-tag (TMT) urinary proteomics to predict severe outcomes in hospitalized COVID-19 patients. We further integrate urine sediment single-cell RNA sequencing (scRNA-seq) from a subset of samples in the urinary dataset along with a larger independent plasma proteomics dataset to identify complementary dysregulated mechanisms. We hypothesize that integrating single-cell transcriptomics with proteomics will uncover pathophysiological mechanisms that can be assigned to specific cell types. We further incorporate scRNA-seq data from kidney organoids infected with SARS-CoV-2 as a physiologically relevant *in vitro* platform to validate the mechanistic signatures observed in the integrated molecular analytics of clinical biospecimens.

## Methods

### Cohort description

We collected urine samples from 130 COVID-19 PCR-positive hospitalized participants between April 2020 and April 2021 and 26 non-COVID AKI patients between May 2021 and August 2021 from two sites, namely Mount Sinai Hospital in New York, NY and University of Michigan in Ann Arbor, MI, and 13 healthy participants from the Mount Sinai Hospital. Inclusion criteria for sample collection was age > 18 years, hospitalization, adult without capacity if a legally authorized representative (LAR) is available, provided they comply with institutional policy. Patients with Stage 5 CKD or end-stage kidney disease (ESKD) were excluded. Most of the COVID-19 patients were male and Caucasian with a median age of 58 ± 15 years.

This study was approved by the Icahn School of Medicine at Mount Sinai Program Institutional Review Board (IRB) under the study number 20-00523 and the University of Michigan Medical School Institutional Review Boards approval under HUM00004729. Detailed methods can be found in the Supplementary Materials.

## Results

### Isobaric urinary proteomic signature of severe COVID-19

We identified 34,474 unique peptides representing 6,196 unique urinary proteins using quantitative TMT-16 plex mass spectrometry (MS) analysis on 130 COVID-19 urine samples collected at two medical centers. The number of unique proteins identified across thirteen TMT-16 plex batches were found to be consistent (**Figure S1**). COVID-19 samples across multiple TMT-16 plex batches clustered together while displaying a significant separation from the instrument controls (**Figure S2**). Therefore, the samples from both collection sites were pooled for downstream analysis. For constructing a predictive algorithm, COVID-19 samples collected less than eighteen days after hospitalization (n=122, median = 3d, interquartile range (IQR) = 2-5d) were stratified into severe (major adverse events), and mild (no major adverse events) composite outcomes as shown (**Figure 1A**). The samples were then randomly allocated at a 2:1 ratio between the discovery (n=81) and validation (n=41) cohort while maintaining equivalent demographic (race, sex, age) distribution between the two groups (**Table 1, Table S1**). While COVID-19 samples clustered separately from the healthy samples, there was no clear separation based on the whole proteome between severe and mild COVID-19 samples (**Figure 1B**). Differential testing using *limma* ^25^ on the discovery set revealed differentially abundant proteins (DAPs) between the severe and mild outcome cohort indicating that there may be dysregulated pathways leading to severe COVID-19 (**Figure 1C**). Unsupervised clustering of all samples in the discovery set (n=81) using the top 50 DAPs showed very subtle separation of mild and severe outcome cases, however, almost 70% of the severe outcome cases clustered towards the right side of the heatmap (**Figure 1D**).

**Figure 1:**
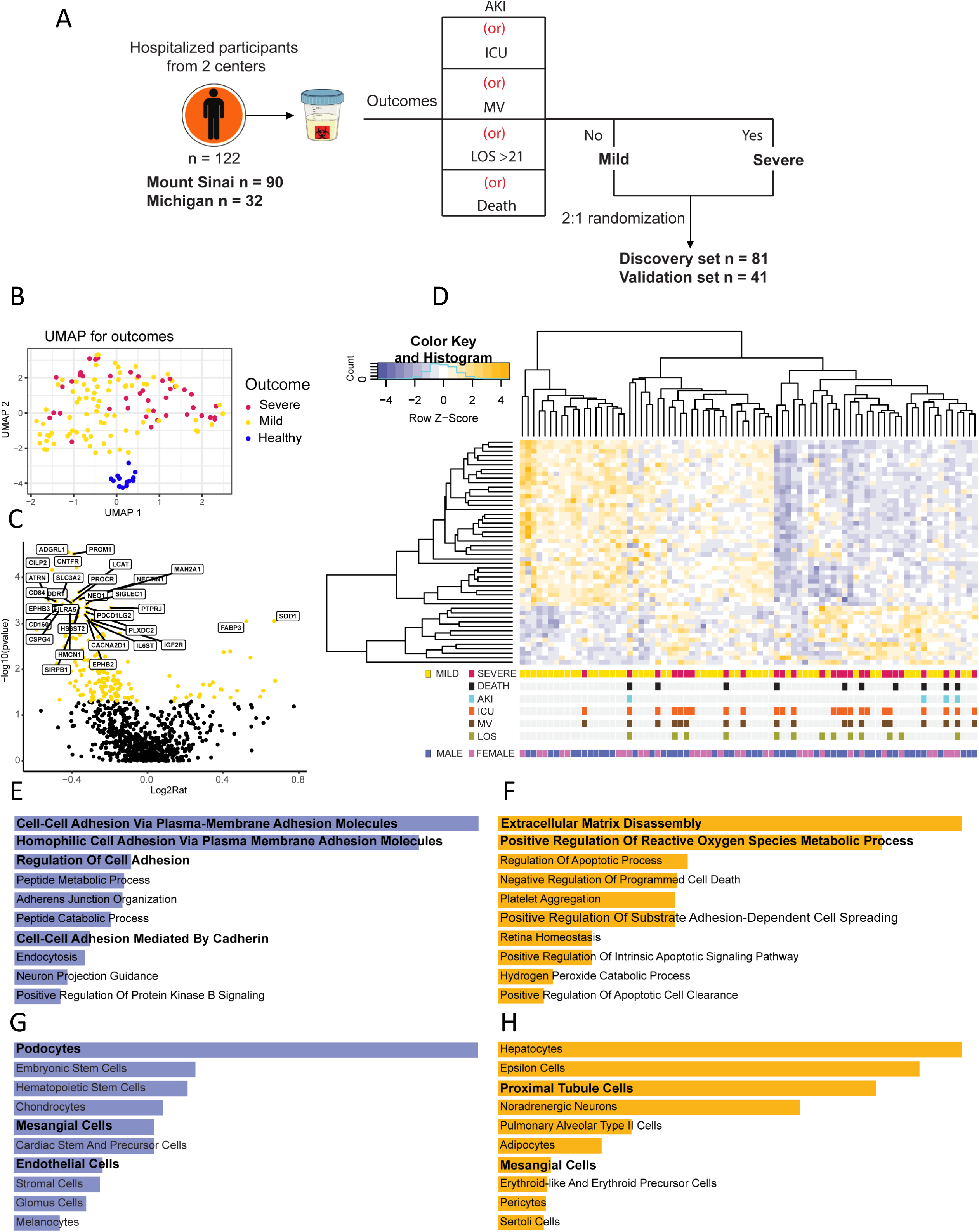
Urinary proteomics reveal differentially abundant proteins (DAPs) between severe and mild COVID-19. **(A)** Schematic representation of stratification of severe and mild outcomes for machine learning (ML) analysis, **(B)** Uniform Manifold Approximation and Projection (UMAP) shows that samples from COVID-19 patients cluster separately from samples from healthy participants, **(C)** Volcano plot of DAPs in severe vs mild outcome groups; **(D)** Unsupervised clustering heatmap of the top 50 DAPs from all the samples in the discovery cohort showing distribution of composite and individual outcomes; Gene ontology biological processes (GOBP) enrichment analysis of **(E)** downregulated and **(F)** upregulated DAPs in severe COVID-19; Cell types associated with **(G)** downregulated and **(H)** upregulated enrichment according to the PanglaoDB database.

**Table 1:** Urine proteomics sample demographics.

|  |  | **Discovery Set** | **Validation Set** |
| --- | --- | --- | --- |
|  | **Number of samples** | 81 | 41 |
| **Site** | **Mount Sinai Hospital** | 60 (74%) | 30 (72%) |
|  | **University of Michigan** | 21 (26%) | 11 (28%) |
| **Sex** | **Male** | 48 (59%) | 25 (64%) |
|  | **Female** | 33 (41%) | 14 (36%) |
| **Race** | **Asian** | 6 (7.4%) | 1 (2.4%) |
|  | **Black** | 22 (27.1%) | 12 (29.2%) |
|  | **Hispanic** | 1 (1.2%) | 0 (0%) |
|  | **White** | 30 (37%) | 19 (46.3%) |
|  | **Other** | 17 (20.9%) | 8 (19.5%) |
|  | **Unknown** | 5 (6.1%) | 1 (2.4%) |
| **Age, years, mean ± SD** | | 57 ± 15 | 58 ± 15 |
| **Composite Outcomes** | **Mild** | 54 (66.67%) | 27 (65.8%) |
|  | **Severe** | 27 (33.33%) | 14 (34.14%) |

Gene ontology biological process (GOBP) enrichment analysis using the DAVID database ^26^ of the top 150 DAPs ranked by absolute value of log2fold-change (log2FC) in COVID-19 versus healthy participants consistently showed significant downregulation of proteolytic and metabolic processes and upregulation of immune processes (**Figure S3**). Enrichment analysis using Enrichr ^27–29^ of DAPs in severe versus mild COVID-19 showed significant upregulation of extracellular matrix (ECM) disassembly, reactive oxygen species and apoptosis associated processes and downregulation of cell adhesion associated processes (**Figure 1E, F, S4**). Downregulated DAPs were associated strongly with kidney cells such as podocytes and mesangial cells in addition to endothelial cells (EC) (**Figure 1G**). Upregulated DAPs were associated with kidney proximal tubular (PT), mesangial cells in addition to hepatocytes and pulmonary alveolar cells (**Figure 1H**).

### Machine learning model predicts major adverse outcomes in COVID-19 patients

To construct the ML model to predict COVID-19 severity, we identified features from the top DAPs obtained from *limma* comparisons using the Boruta feature selection method ^30^. A set of 12 features (i.e., proteins) was identified as predictors of severity, encoded by the following transcripts: ADGRL1, PROM1, PLS3, NECTIN1, EPHB2, SCGB1A1, PLXDC2, LCN1, PDCD1LG2, CILP2, FAM151A and PTPRJ (**Figure 2A, Table S2**). These features were used for random forest model construction within the discovery set with 10-fold cross-validation. The generated receiver operating characteristic (ROC) curves show that the algorithm demonstrated good predictive power for both discovery and validation set with Area Under the Curve (AUC) of 87% and 76% respectively (**Figure 2B-C).** By setting one-cutoff according to ROC curve in discovery set, the model achieved Negative Predictive Power (NPV) of 89% and 83% in the discovery and validation set respectively (**Table 2**). Our model was also able to accurately predict 78% of all severe-outcome patients (True Positive Rate, TPR) (**Figure 2D**). Further, we derived a tertile cutoff to estimate prediction accuracy, achieving a Positive Predictive Power (PPV) of 85% and 75% at a high cutoff (0.653), an NPV of 97% and 88% at the low cutoff (0.247) in the discovery and validation sets, respectively (**Figure 2E**, **Table 3**). While over 80% of the samples in this study were collected less than 5 days after hospitalization, we did not observe any significant correlation between prediction accuracy and the number of days between hospitalization and sample collection (**Figure S5A**). While prediction accuracy for individual outcomes were about 70%, 100% of samples with AKI as an outcome were accurately predicted demonstrating a potential in predicting kidney-specific adverse events (**Figure S5B**).

**Figure 2:**
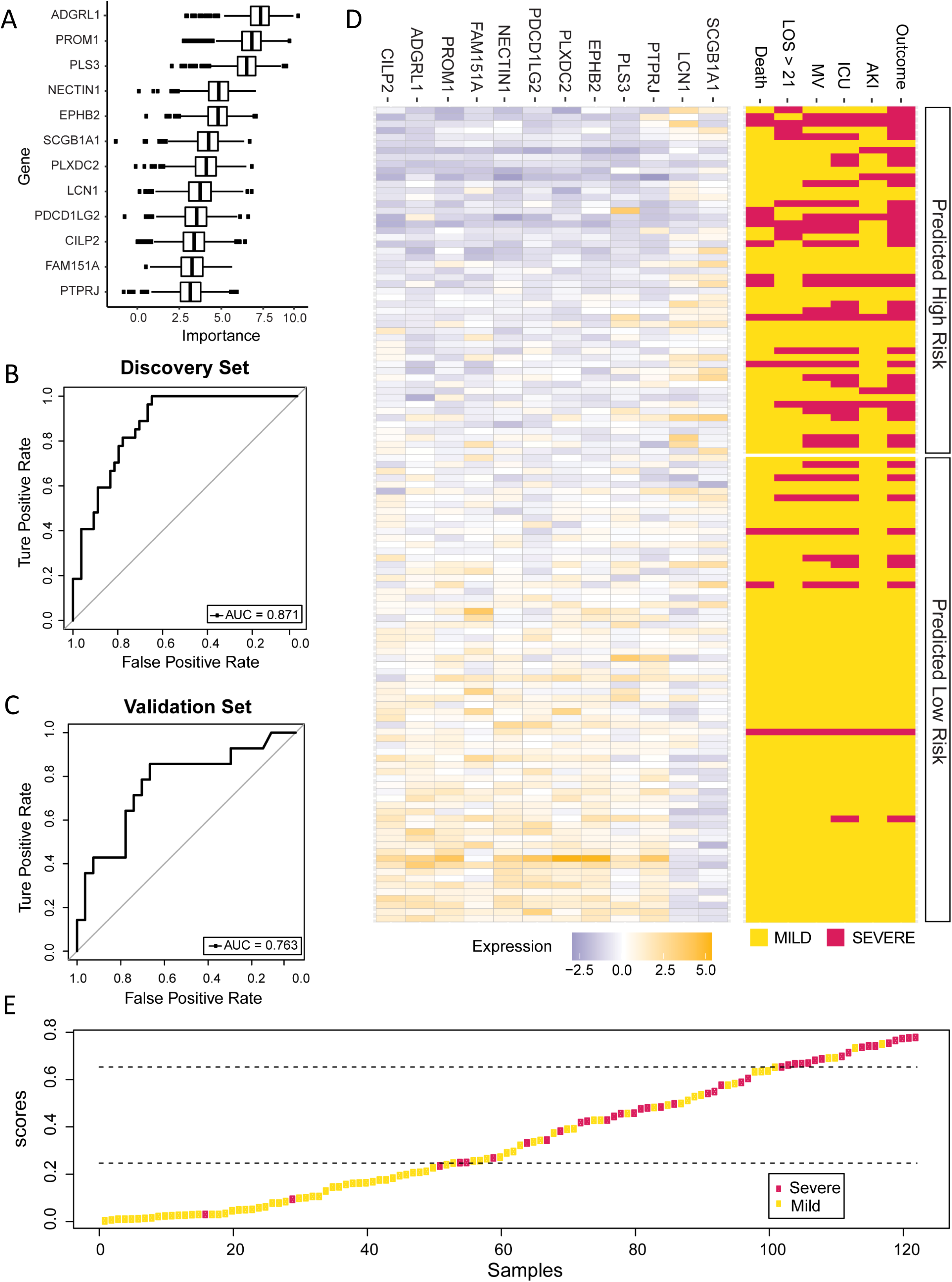
Cross-validated random forest model reliably predicts COVID-19 severity. **(A)** Top-12 features selected using Boruta feature selection method; Receiver Operating Characteristics (ROC) curves of the **(B)** discovery and **(C)** validation sets; **(D)** Expression levels of the top-12 features in individual samples correlated with severe composite outcomes and prediction accuracy. **(E)** A two-tiered cutoff model, where scores < 0.653 or > 0.247 are used for accuracy calculation, further improved prediction accuracy.

**Table 2:** Predictive accuracy of the ML model.

| **Set** | **TPR** | **FPR** | **PPV** | **NPV** |
| --- | --- | --- | --- | --- |
| Discovery | 0.815 | 0.222 | 0.647 | 0.893 |
| Validation | 0.714 | 0.259 | 0.588 | 0.833 |
| Cutoff | 0.392 |  |  |  |

**Table 3:** Predictive accuracy of the two-cutoff tiered ML model.

| **Set** | **PPV** | **NPV** |
| --- | --- | --- |
| Discovery | 0.846 | 0.972 |
| Validation | 0.75 | 0.875 |
| Cutoffs | 0.653 | 0.247 |

### Severe COVID-19 ML features recapitulate injury mechanisms of non-COVID AKI

GOBP enrichment analysis showed that the top ML features identified were associated with pathophysiological mechanisms such as cell adhesion and cell component organization (**Figure S6**). ADGRL1, EPHB2, CILP2 and NECTIN1 have been previously reported to have functions in cell-cell adhesion and cell communication ^31–35^. Similarly, PLXDC2 and PTPRJ have been shown to be associated with angiogenesis-associated pathways in tumors ^36–38^. Some of these features such as PLS3, ^39^ NECTIN1, ^40^ PLXDC2, ^41^ and PTPRJ ^42^ have also been previously found to be differentially regulated in COVID-19. The publicly available Kidney Precision Medicine Project (KPMP) scRNA-seq database showed that many of these features are highly expressed in glomerular cells such as ECs, podocytes, and parietal epithelial cells (PECs) (**Figure S7**).

Previous studies have reported a decline in eGFR in severe COVID-19 and COVID-associated AKI within 30 days post-acute infection ^43, 44^. The eGFR of the severe acute COVID-19 cohort in our dataset was found to be significantly lower than that of the mild cohort (**Figure S8**). Since it was recently reported that AKI in severe COVID-19 shared mechanistic similarities with sepsis-associated AKI ^45, 46^, we sought to look at the mechanistic similarities between the severe COVID-19 and the non-COVID AKI urinary proteome. While the separation between non-COVID AKI and COVID-19 samples was not extensive, we observed a clear distinction between disease and healthy samples (**Figure S9A**). An unbiased unsupervised k-means clustering projected over the PCA plot clearly showed that a substantial proportion of severe COVID-19 and non-COVID AKI samples cluster together (**Figure S9B-C**). The top 150 DAPs ranked by absolute log2FC and p-value<0.05 in non-COVID AKI vs healthy participants showed a significant overlap of downregulated enrichments associated with cell adhesion and proteolysis pathways observed in severe COVID-19 (**Figure S10**). Urinary albumin abundance was significantly higher in severe and mild COVID-19 and non-COVID AKI patients compared to healthy participants, indicating altered kidney function in diseased patients (**Figure S11A**). Cystatin C, CST3, a commonly used kidney injury biomarker was also slightly, but not significantly elevated in severe COVID-19 and non-COVID AKI urine samples (**Figure S11B**) ^47, 48^. Abundance of ten out of twelve ML features were similarly downregulated in non-COVID AKI and severe COVID-19 urine samples (**Figure 3**). Taken together, these indicate that mechanisms associated with COVID-19 severity may have strong overlap with the kidney injury mechanisms involved in the progression of AKI. Further, validation of the features in an external AKI cohort from the KPMP database showed that 5 (NECTIN1, EPHB2, PLXDC2, PDCD1LG2, PTPRJ) out of the 9 features identified in both datasets were similarly recapitulated (**Figure S12**).

**Figure 3:**
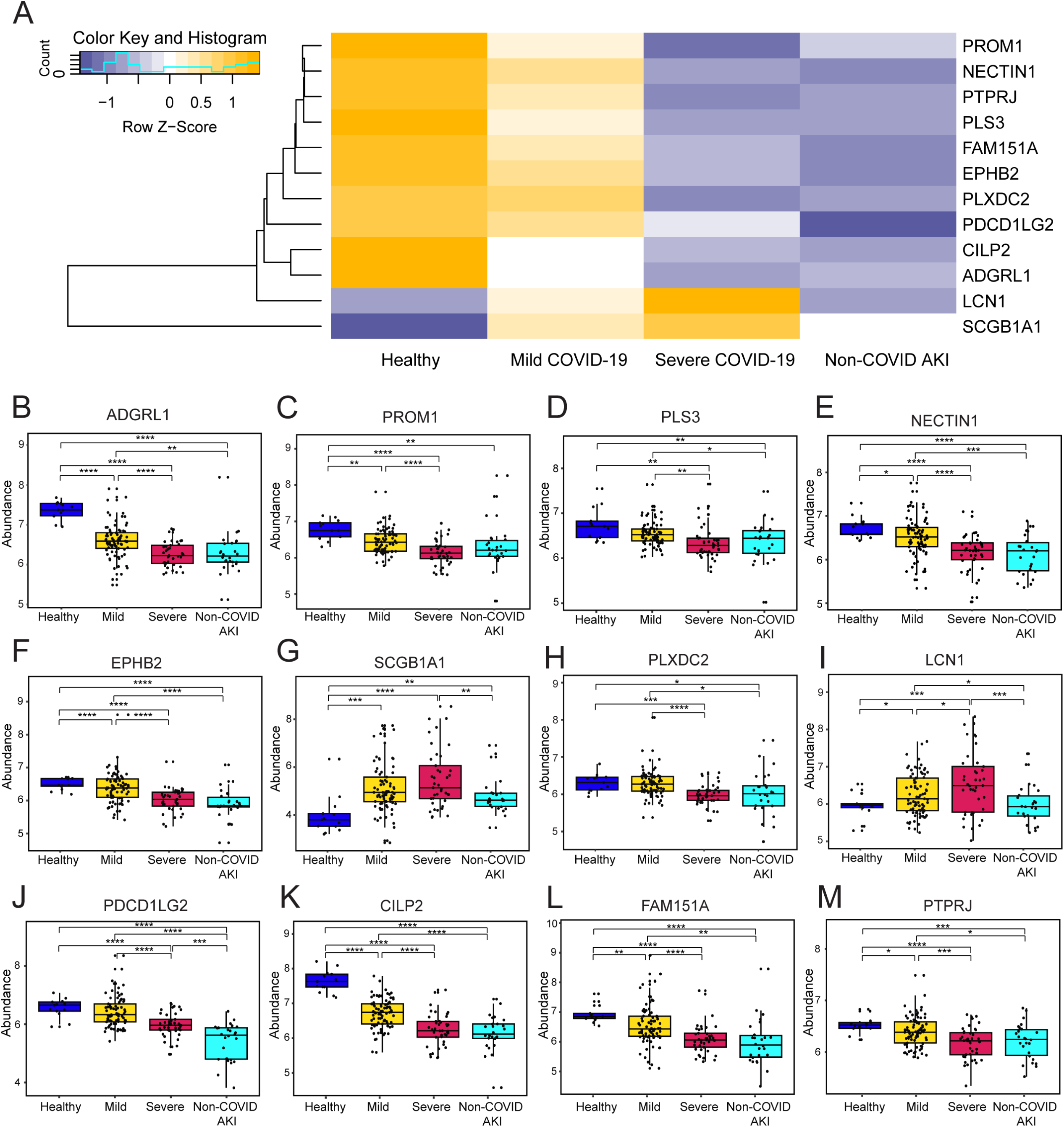
Abundances of top machine learning (ML) features are recapitulated in non-COVID AKI. **(A)** Unsupervised clustering heatmap and **(B-M)** individual box plots of abundance levels of the top-12 features in healthy, mild, severe COVID-19 and non-COVID AKI urine samples.

### Integrated multiomic analysis of clinical datasets show adhesion-associated processes highly impacted in severe COVID-19

In addition to predicting COVID-19 severity, urinary biomarkers identified here uncovered overlapping mechanisms in severe COVID-19 and non-COVID AKI. We recently showed that the plasma proteome can be used to characterize AKI and eGFR decline in COVID-19 hospitalized patients ^17^. We hypothesized that integrating DAPs from the plasma and the urine proteome will uncover cohesive mechanisms of kidney injury in these two independent datasets. The top 150 urine and 450 plasma proteomics DAPs ranked by absolute log2FC and p-value<0.05 were analyzed using the standard enrichment program in the Molecular Biology of the Cell Ontology (MBCO) database (**Figure 4A, Figure S13**). ECM homeostasis and coagulation-associated pathways were common between both proteomic datasets in addition to multiple pathways within the top 25% interactions. The Humanbase kidney-specific functional network discovery analysis resulted in five distinct gene clusters with more than 43 genes in each cluster (**Figure S14**). HumanBase integrates public genomic datasets into a tissue-specific functional module where individual gene clusters share a local network neighborhood. We found significant associations between genes within multiple clusters associated with terms such as autophagy, cell-matrix adhesion, regulation of proteolysis, and receptor-mediated endocytosis. We further used the publicly available KPMP scRNA-seq database to identify the kidney cell type specificity of the top 50 DAPs from the urine and plasma proteomics datasets. While the plasma proteins showed high specificity to immune and interstitial cells, urinary proteins were highly specific to kidney cell types such as ECs, podocytes, PT, and distal tubules (DCT) (**Figure 4B**).

**Figure 4:**
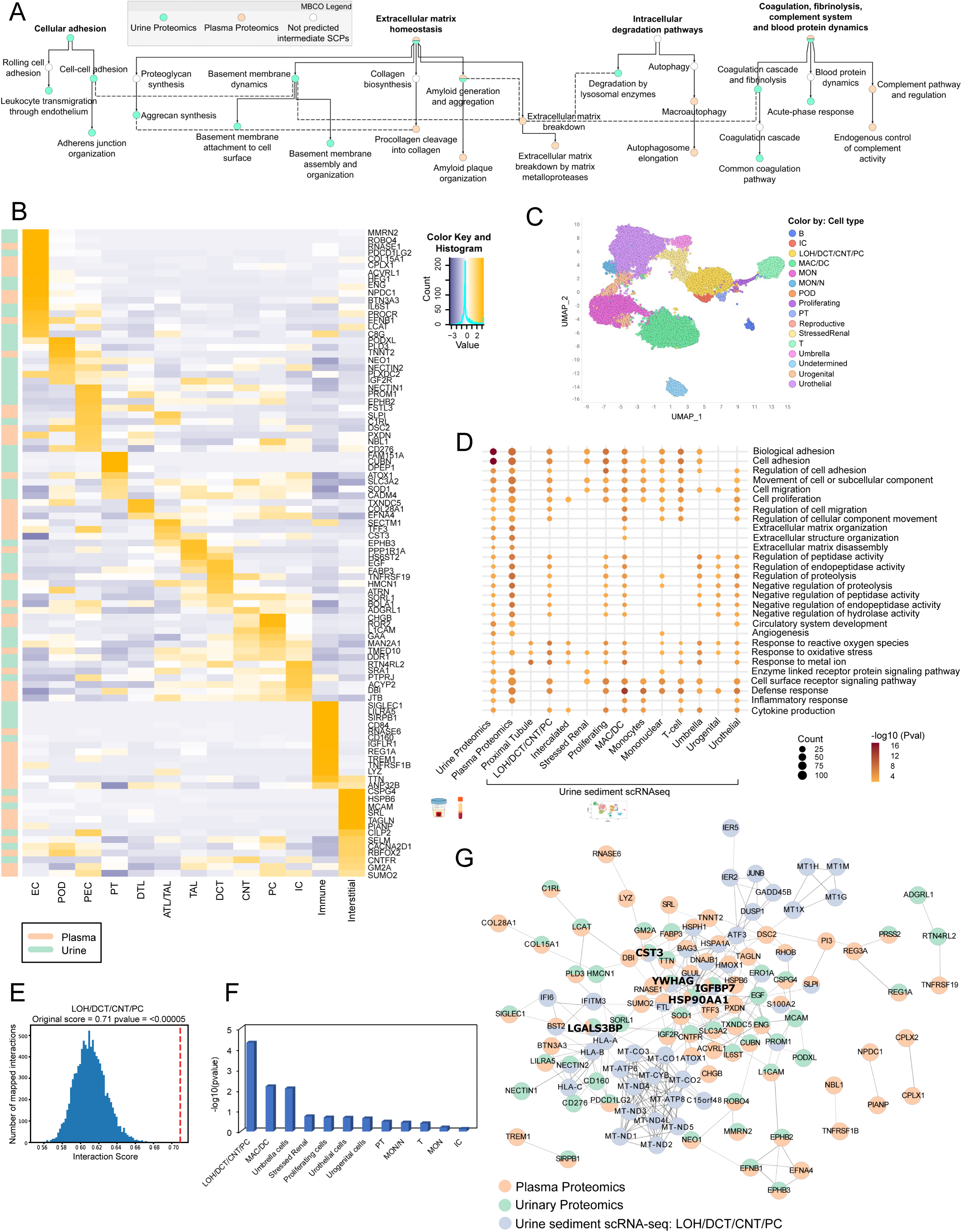
Integrative multiomic analysis reveals overlapping dysregulated pathways associated with COVID-19 severity. **(A)** Interaction of the top Urine proteomics differentially abundant proteins (DAPs) and the top-450 plasma proteomics DAPs analyzed using the Molecular-Biology-of-the-Cell-Ontology (MBCO) standard enrichment. Pathways among the top 25% interaction were connected by a dashed line. **(B)** Supervised clustering of Kidney Precision Medicine Project (KPMP) single cell RNA sequencing (scRNA-seq) data of the top 50 DAPs from urine and plasma proteomics datasets across different kidney cell types. **(C)** UMAP of urine sediment scRNA-seq showing diverse population of kidney, urinary and immune cell types. **(D)** Functional overlap analysis of top 150 DAPs from urine proteomics, top 450 DAPs from plasma proteomics and top-150 differentially expressed genes (DEGs) from individual cell type cluster from urine sediment scRNA-seq. **(E)** Histogram of the mean values of the permuted network scores along with original interaction score (red) of top 50 DEGs in LOH/DCT/CNT/PC cluster and top 50 DAPs from urine and plasma. **(F)** Bar plot of –log10(pvalue) showing the significance of interaction of top 50 DEGs from individual cell types in urine sediment scRNA-seq dataset with top 50 DAPs in urine and plasma proteomics datasets. **(G)** Protein-protein interaction (PPI) network of top 50 DEGs in urine scRNA-seq LOH/DCT/CNT/PC cluster and top 50 DAPs in urine and plasma proteomics. Proteins identified in all 3 datasets is highlighted in bold. Cell type abbreviations: Endothelial cell (EC), Podocytes (POD), Parietal epithelial cells (PEC), Proximal tubular cells (PT), loop of Henle (LOH), descending thin limb (DTL), ascending thin limb (ATL), thick ascending limb (TAL), distal convoluted tubule (DCT), connecting tubule (CNT), principal cell (PC), intercalated cells (IC), macrophages (MAC), dendritic cells (DC), monocytes (MON), mononuclear cells (MON_N).

Since obtaining kidney biopsy samples from these patients would have been invasive and challenging, we adopted urine sediment scRNA-seq to identify cell-type specific transcriptomic changes due to severe COVID-19. Urine sediment scRNA-seq has been shown to be a powerful tool for investigating the differential transcriptomic signature of the injured kidney cells in the context of AKI as well as COVID-AKI ^49–51^. We obtained urine sediment scRNA-seq data from a subset of urine samples (n=40) analyzed using proteomics. Sixteen distinct cell clusters consisting of kidney cells, immune cells and cells from the urinary tract were identified (**Figure 4C**). Clusters containing reproductive cells and unidentified cells were removed before further analysis. Podocytes were excluded from the differentially expressed genes (DEG) analysis due to the insufficient number of cells obtained. We identified five kidney cell-type clusters such as PT cells, intercalated cells (IC), proliferating cells, stressed renal cells and a hybrid cluster containing loop of Henle, DCT, connecting tubule and principal cells (LOH/DCT/CNT/PC). We also identified four immune cell clusters such as T, macrophages, and dendritic cells (MAC/DC), monocytes (MON), and mononuclear (MON/N cells) and three clusters of urinary tract cells such as umbrella, urothelial and urogenital cells. The top 150 DAPs from urine, 450 DAPs from plasma proteomics and top 150 DEGs from individual cell type clusters of the urine sediment scRNA-seq were analyzed using functional overlap analysis (**Figure 4D**). Functional enrichment showed significant overlap between several pathways such as cell adhesion, migration, proliferation, ECM organization, regulation of proteolysis, response to oxidative stress (ROS), and inflammatory response among others between urinary and plasma proteomics. The LOH/DCT/CNT/PC cluster and MAC/DC cluster from the urine sediment scRNA-seq showed significant overlap with urine and plasma proteome within these pathways. KPMP analysis showed that genes within the adhesion and migration pathways were highly specific to immune clusters followed by EC and podocytes (**Figure S15A-B**). Several genes associated with ROS pathway showed a highly specific expression in the PT cluster, suggesting that there is an increase in stress response in the PT due to severe SARS-CoV-2 infection (**Figure S15C**).

We further built a protein-protein interaction (PPI) network for the top 50 DEGs from individual cell types in the urine sediment scRNA-seq and the top 50 DAPs from the urine and plasma datasets PPI interaction information in STRING database. Permuted network scores based on interactions found in the STRING database showed a highly significant original network interaction score for LOH/DCT/CNT/PC cluster (**Figure 4E, S16**), followed by MAC/DC and umbrella cells (**Figure 4F**). Heatmap of enrichments associated with the top DEGs within the LOH/DCT/CNT/PC cluster showed associations with metal ion homeostasis, apoptotic processes, and ROS, among others (**Figure S17**). Humanbase kidney-specific functional module discovery and the MBCO standard enrichment analysis of top features in LOH/DCT/CNT/PC cluster and the urine and plasma proteomics datasets showed significant overlapping interactions between multiple pathways such as ECM homeostasis and intracellular degradation pathways (**Figure S18-S19**). The PPI network of the LOH/DCT/CNT/PC cluster and the urine and plasma proteomics datasets revealed highly interacting proteins among which we identified five proteins that were identified in all the three datasets: CST3, YWHAG, IGFBP7, LGALS3BP and HSP90AA1 (**Figure 4G**). Three out of these five proteins (CST3, LGALS3BP and IGFBP7), all cluster in the same module of the Humanbase functional module analysis demonstrating a shared function in ECM organization (**Figure S20**). Cystatin C (CST3) has been previously also shown to be a predictive biomarker for COVID-19 severity ^47, 48^. IGFBP7, a glycoprotein expressed in ECs and renal epithelial cells, has been extensively shown as a prognostic biomarker for early AKI along with TIMP2 ^52, 53^. LGALS3BP, an ECM associated glycoprotein, was shown to be differentially regulated in COVID-19 samples^19, 54^. The data from the multiomic integrative overlap and PPI network analysis show that ECM and degradation-associated pathways are highly impacted due to severe COVID-19, and that the tubules of the juxtamedullary nephron are likely the most susceptible cell types impacted due to these pathways leading to major adverse kidney events.

### iPSC-derived kidney organoids recapitulate mechanisms of injury observed in clinical datasets

Next, we sought to validate the proteomic and transcriptomic signature observed in the clinical datasets due to COVID-19 infection by adopting iPSC-derived kidney organoids as an *in vitro* disease model. While the urine sediment scRNA-seq data demonstrated the transcriptomic changes in the injured cell types shed in urine, a kidney organoid model would allow us to study transcriptional changes in a physiologically relevant context *in vitro*. Previous studies have established that SARS-CoV-2 directly infects kidney organoids and drives fibrosis within the organoids ^22, 23^. Kidney organoids expressed tubular and glomerular markers 18 days after differentiation (**Figure 5A-B**). After 25 days of differentiation, kidney organoids infected with SARS-CoV-2 showed sustained viral titer levels up to 6 days post infection (**Figure S21**). Immunofluorescence staining showed presence of SARS-CoV-2 nucleocapsid (SARS NP) protein in the LTL+ tubular cells, suggesting that they may be the main target of SARS-CoV-2 infection in the organoids (**Figure 5C**).

**Figure 5:**
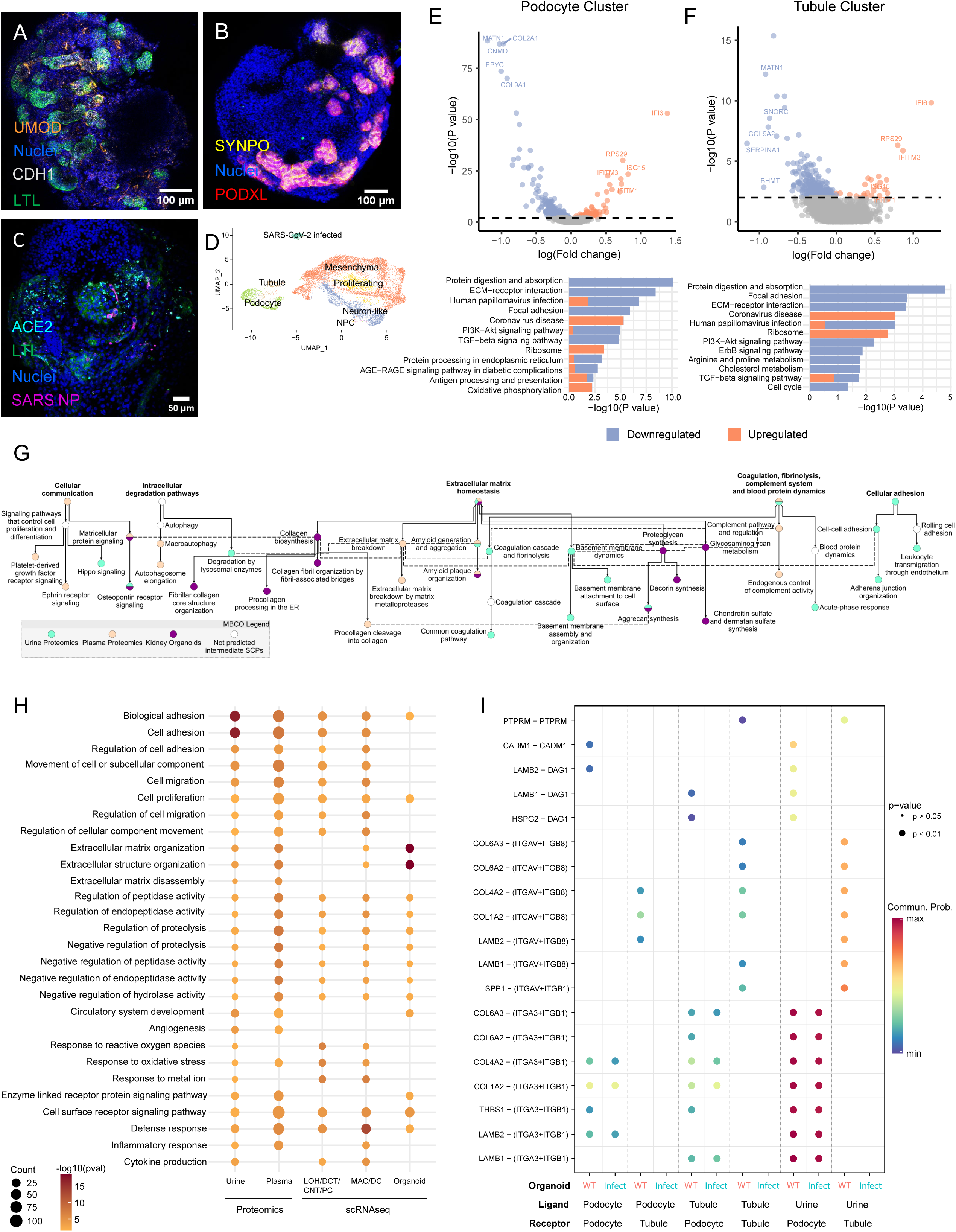
Kidney organoids infected with SARS-CoV-2 recapitulate mechanisms of kidney injury observed in clinical datasets. **(A)** Immunofluorescence confocal images of iPSC-derived kidney organoids expressing tubular markers such as lotus tetragonolobus lectin (LTL, proximal tubule, green), E-cadherin (CDH1, distal tubule, gray), uromodulin (UMOD, loop of Henle, orange) and **(B)** podocyte markers such as synaptopodin (SYNPO, yellow) and podocalyxin (PODXL, red). **(C)** SARS-CoV-2 infected LTL (green) positive cells express SARS-CoV-2 nucleocapsid protein (NP, magenta) and angiotensin converting enzyme 2 (ACE2, cyan). **(D)** tSNE plot of scRNA sequencing of kidney organoids. KEGG pathway analysis of top differentially expressed genes (DEGs) in **(E)** podocyte and **(F)** tubular clusters upon SARS-CoV-2 infection. **(G)** Interaction of top 150 kidney organoid DEGs, top 150 urine proteomics DAPs and top 450 plasma proteomics DAPs analyzed using MBCO standard enrichment. Shown here are SCPs among the top 25% interactions connected by a dashed line. **(H)** Functional overlap analysis of urine, plasma, organoid datasets along with LOH/DCT/CNT/PC and MAC/DC clusters from urine sediment scRNA-seq datasets. **(I)** Ligand-receptor interaction analysis within podocyte and tubule clusters in the organoids as well as in the urine proteomics dataset.

The scRNA-seq analysis identified 18 cell clusters consisting of kidney cell types such as *NPHS1*+ and *NPHS2*+ podocytes and *LRP2*+ tubules in addition to *PDGFRB*+ mesenchymal cells, *MKI67*+ proliferating cells, and off-target populations such as neuron-like and neuronal progenitor cells (NPC) (**Figure S22A**). Unsupervised clustering showed that SARS-CoV-2 infected cells cluster separately (**Figure S22B**). Upon removal of COVID genes, SARS-CoV-2 infected cells redistributed to PT, podocytes, and mesenchymal cell clusters (**Figure 5D**,). Heatmap of the top 150 DEGs ranked by absolute log2FC and p-value <0.05 in the infected vs WT kidney organoids showed ECM-associated processes were highly enriched (**Figure S23**). Differential transcriptomic signatures and KEGG pathway analysis of the podocyte and tubule clusters demonstrated a strong overlap in dysregulated pathways observed in the urine and plasma proteomic datasets (**Figure 5E-F**). We further performed MBCO standard enrichment analysis and Humanbase kidney-specific functional module discovery of top 150 DEGs in kidney organoids and the top 150 urine and 450 plasma DAPs (**Figure 5G, Figure S24-25**). Both analyses showed a significant overlap between ECM homeostasis and cellular communication pathways consistent with prior observations. Functional overlap analysis of the two proteomic (urine and plasma) and the two transcriptomic (urine sediment and kidney organoid) datasets showed overlap of ECM as well as proteolysis and peptidase-associated pathways indicating a potential maladaptive reorganization of the ECM through the activity of peptidases **(Figure 5H)**. These data corroborate the fibrotic response observed in previously published kidney organoid models of SARS-CoV-2 infection ^23^. Ligand-receptor interaction analysis of the tubule and podocyte clusters showed significant reduction and loss of crosstalk and signaling between ECM components such as collagen and laminin and their integrin receptors (**Figure 5I**). Within the podocyte cluster, loss of interaction between *LAMB2* and *DAG1* as well as *THBS1* and *ITGA3*/*ITGB1* indicated a potential impact on podocyte attachment to the basement membrane. Within the tubular cluster, we observed a greater impact on several integrin receptors and loss of communication between integrin and collagen or laminin. To identify whether these interactions are recapitulated in the clinical dataset, we investigated urine-derived ligands and organoid-derived receptors and found a significant loss of integrin and ECM interactions specifically within the tubular cluster of the kidney organoids.

## Discussion

In this multicenter study, we combined ML and multiomics datasets to predict COVID-19 severity and further identify kidney pathophysiological mechanisms underlying severe COVID-19. Our dataset from 130 COVID-19 patients represents one of the most extensive and deepest human urinary proteomic sets published to date. For severe outcome stratification, general clinical metrics such as ICU, MV, LOS, and death were used in addition to kidney specific outcome such as AKI after hospitalization. Renal replacement therapies such as dialysis were observed in a subset of COVID-AKI patients in our cohort and did not increase predictive power. Using a 10-fold cross validated random forest model, we demonstrated accurate prediction of severe outcomes within five days of hospitalization in over 80% of participants; we note that for the Mount Sinai cohort, majority of severe adverse events were observed 5-12 days after hospitalization. In addition to prediction of COVID-19 severity, 10 out of 12 biomarkers identified in our ML analysis revealed intersecting mechanisms of kidney injury in severe COVID-19 and non-COVID AKI. This was further validated in an independent external AKI dataset within the KPMP database where 5 out of 9 predictive urinary proteins were similarly recapitulated. Two of the 12 features, SCGB1A1 and LCN1, were upregulated in severe COVID-19 patients in the urine proteomics dataset. LCN1 was also significantly elevated in the plasma proteomics dataset. However, there was no significant difference in non-COVID AKI versus mild COVID-19 samples. SCGB1A1 and LCN1 are secreted proteins that have been shown to be dysregulated in infection related pathways, ^55–57^ and may not be relevant to mechanisms involved in other types of AKI.

Given the kidney-specific signature of the biomarkers, we investigated the presence of viral proteins in the urine. However, we only identified SARS-CoV-2 spike protein in 6 out of 130 COVID-19 samples. While we did not observe a significant difference in ACE2 expression in control, severe, and mild urine samples, TMPRSS2 was significantly downregulated in severe compared to mild COVID-19 and healthy urine samples indicating active SARS-CoV-2 infection ^58^ (**Figure S26**). Similarly, kidney injury marker-1 (KIM-1) expression was found in a significantly higher proportion of severe (27%) and non-COVID AKI (50%) samples compared to mild (16%) samples. The top DAPs between severe and mild COVID-19 showed significant enrichment of cell adhesion-associated pathways along with vascular function and angiogenesis. The enrichment of these pathways indicates potential endothelial dysfunction-mechanisms driving COVID-19 severity. From the KPMP cell type specificity analysis, we found that 10% of the top 100 DAPs from our urine and plasma datasets showed high expression in the ECs further confirming that vascular function may be highly impacted due to severe COVID-19.

To investigate the impact of COVID-19 infection on kidney specific cell targets, we integrated transcriptomic datasets from urine sediments and kidney organoids along with urine and plasma proteomic datasets. While previous studies have shown that PT cells are the main target of COVID-19 associated kidney injury ^59, 60^, in this study we further identify that the cells in the juxtamedullary nephron such as LOH, DCT, CNT and PC may also be significantly impacted early due to infection. Functional overlap analysis showed that ECM homeostasis, coagulation, and degradation pathways were highly dysregulated across all four datasets. Cohesive dysregulation of ECM-associated pathways suggests a profibrotic response due to severe COVID-19 infection as reported previously ^22, 23^, with a significant fibrotic response from the kidney organoid dataset. In addition to this, we also found an elevated expression of metallothionein genes in the hybrid LOH/DCT/CNT/PC cluster. In addition to maintaining metal ion homeostasis, metallothionein genes regulate autophagy and are upregulated in response to oxidative stress ^61^. Based on these data, it is possible that there is metallothionein-induced autophagy response from tubular cells in the medullary nephron in response to the infection.

This study has several limitations. The effect of comorbidities on severe outcomes in the sample population was not accounted for during the construction of the prediction model. It has been previously shown that patients with high-risk alleles of *APOL1* are more prone to COVID-associated AKI ^62, 63^. However, the *APOL1* status of participants in our cohort is unknown. Due to differences in sample collection times between the two centers, predictive power of the ML model was lower when the Sinai cohort was used as the discovery set and the Michigan cohort was used as the validation set (**Figure S27, Table S3**); however, our data showed that the time of sample collection after hospitalization did not impact prediction accuracy. Further, GOBP enrichments associated with ML features from this analysis also indicated similar mechanisms were dysregulated indicating that our conclusions would remain unchanged (**Figure S28**). While the proteomic datasets showed high expression of endothelial proteins, given the difficulty of capturing ECs in either specimen used in transcriptomic analysis, we were unable to validate the signature associated with COVID-induced vascular dysfunction. We acknowledge that kidney organoids in their current state do not represent mature compartments such as immune cells and vasculature and contain off-target cell populations. However, our results suggest that the SARS-CoV-2 induced dysfunction of adhesive processes is a general cell biological phenomenon and that organoids were able to sufficiently recapitulate the cohesive responses observed in the clinical datasets *in vitro*. Finally, we note that while our sample size may be limited compared to some of the larger targeted biomarker studies, we note that our mass spectrometry-based approach not only represents the most extensive urinary mass spectrometry data, but it also includes post-translational modification and peptide localization information, which could provide useful for future biomarker discovery and development.

In conclusion, we use multiple omics datasets and an integrative approach to uncover cohesive dysregulation of ECM and adhesion-associated pathways within multiple kidney cell types due to acute COVID-19 infection. Using orthogonal assays such as urinary proteomics and urine sediment scRNA-seq, we were able to filter out systemic response from the immune system and delineate kidney cell type-specific pathways that are impacted by severe COVID-19. While predictive urinary biomarkers identified in this study may not be extensively utilized for acute COVID-19 given the low hospitalization rates, as functional biomarkers, they may be useful for monitoring progression of long-COVID-associated kidney dysfunction. The validation of these biomarkers in an independent external AKI cohort also demonstrates their utility in kidney diseases beyond COVID-19 associated kidney injury.

## Data Availability Statement

All proteomics raw data will be made available on the ProteomeXchange PRIDE repository (https://www.ebi.ac.uk/pride/). The results here are in part based upon data generated by KPMP: the urinary SomaScan proteomics (6696930a-f707-430d-a964-110aefa93c62_Urine Biomarker Data-SomaScan-2022\Urine Biomarker Data-SomaScan-2022\Data\SS-2342467_2023-11-30_Urine.ANMLNormalized.xlsx) and the fully annotated scRNA-seq data (KidneyTissueAtlas/521c5b34-3dd0-4871-8064-61d3e3f1775a_PREMIERE_Alldatasets_08132021.h5Seurat) downloaded from https://atlas.kpmp.org/.

## Acknowledgements

We gratefully acknowledge the Mount Sinai Hospital and University of Michigan Emergency Department nurses and physicians for their assistance with biospecimen collections. We also thank the fellows in the Department of Medicine, particularly Meghana Eswarappa, and Chip Bowman-Zamora for their assistance with biospecimen collections. We thank Dr. Alecia Muwonge for assistance with consenting patients for the Pred-MAKER study. We thank Randy Albrecht for support with the BSL3 facility and procedures at the Icahn School of Medicine at Mount Sinai (ISMMS). We acknowledge the Genomics Core facility and the Microscopy and Advanced Bioimaging CoRE facility at the Icahn School of Medicine at Mount Sinai for assistance with QC analysis of cDNA and confocal microscopy and analytics, respectively. The scRNA-seq analysis of kidney organoids was performed at the Genomics Resources Core Facility (GRCF) at Weill Cornell Medicine.

We acknowledge funding from NIH R01DK118222 and DoD W81XWH −20-1-0837 (EUA). EUA is partially supported by Kidney Precision Medicine Project (KPMP), which maintains the Kidney Tissue Atlas of publicly available urine somascan and human transcriptomic data that were used in this study. The KPMP is funded by the following grants from the NIDDK: U01DK133081, U01DK133091, U01DK133092, U01DK133093, U01DK133095, U01DK133097, U01DK114866, U01DK114908, U01DK133090, U01DK133113, U01DK133766, U01DK133768, U01DK114907, U01DK114920, U01DK114923, U01DK114933, U24DK114886, UH3DK114926, UH3DK114861, UH3DK114915, UH3DK114937. LC is supported in part by a grant from the NIH/NIDDK (K23DK124645 and U01DK137259). The mass spectrometry data were obtained from an Orbitrap mass spectrometer funded in part by the NIH grant 1S10OD025047-01, for the support of proteomics research at Rutgers Newark campus. This work was partially supported by CRIPT (Center for Research on Influenza Pathogenesis and Transmission), a NIAID funded Center of Excellence for Influenza Research and Response (CEIRR, contract # 75N93021C00014), and by NIAID grants U19AI142733, U19AI135972 and U19AI168631 to A.G-S., by the JPB and OPP foundations and an anonymous philanthropic donor to A.G-S. J. Haydak was supported by NIH T32HD075735 NICHD-Interdisciplinary Training in Systems and Developmental Biology and Birth Defects; A. Mendoza was supported by NIH R01DK131047 Diversity Supplement.

## Author Contributions

EUA, LC, SGC, GNN and JCH designed the study; EUA, LC, GNN and AWC obtained IRB approvals; NA, AS, SD, GM, AM, SL, HHW and JAS enrolled participants and collected patient urine samples; NA, SD and AM processed the urine samples for proteomics; TL and QS performed the LC-MS/MS analysis; ZY performed the ML analysis; ZY and YD validated the ML analysis; NA and SD cultured the iPSC-derived kidney organoids and performed the IF and imaging experiments; LM and TK performed the organoid SARS-CoV-2 infection experiments; LM, TK, JF and NA processed the organoids for scRNA-seq; ZS analyzed the organoid scRNA-seq data; SE, RM, BG, and EAO analyzed the urine single-cell data; ZS performed the functional overlap and ligand-receptor interaction analysis; ZY and PJ performed the network analysis; JH performed the KPMP analysis; NA performed the HumanBase and MBCO analysis; SGC and LC provided clinical input; EUA and WZ provided input for scientific interpretation of results; NA compiled the figures and wrote the manuscript; NA, JH, ZY, ZS, MK, WZ and EUA revised and edited the manuscript. All the authors approved the final manuscript.

## Disclosures

The A.G.-S. laboratory has received research support from GSK, Pfizer, Senhwa Biosciences, Kenall Manufacturing, Blade Therapeutics, Avimex, Johnson & Johnson, Dynavax, 7Hills Pharma, Pharmamar, ImmunityBio, Accurius, Nanocomposix, Hexamer, N-fold LLC, Model Medicines, Atea Pharma, Applied Biological Laboratories and Merck, outside of the reported work. A.G.-S. has consulting agreements for the following companies involving cash and/or stock: Castlevax, Amovir, Vivaldi Biosciences, Contrafect, 7Hills Pharma, Avimex, Pagoda, Accurius, Esperovax, Applied Biological Laboratories, Pharmamar, CureLab Oncology, CureLab Veterinary, Synairgen, Paratus, Pfizer and Prosetta, outside of the reported work. A.G.-S. has been an invited speaker in meeting events organized by Seqirus, Janssen, Abbott, Astrazeneca and Novavax. A.G.-S. is inventor on patents and patent applications on the use of antivirals and vaccines for the treatment and prevention of virus infections and cancer, owned by the Icahn School of Medicine at Mount Sinai, New York, outside of the reported work.

## Notes

### Competing Interest Statement

E.U.A. has received research funding from Renalytix AI and Aurinia Pharmaceuticals; has consulting agreements with Ikena Oncology; has patents and applications owned by Icahn School of Medicine at Mount Sinai, all outside the scope of this manuscript. The A.G.-S. laboratory has received research support from GSK, Pfizer, Senhwa Biosciences, Kenall Manufacturing, Blade Therapeutics, Avimex, Johnson & Johnson, Dynavax, 7Hills Pharma, Pharmamar, ImmunityBio, Accurius, Nanocomposix, Hexamer, N-fold LLC, Model Medicines, Atea Pharma, Applied Biological Laboratories and Merck, outside of the reported work. A.G.-S. has consulting agreements for the following companies involving cash and/or stock: Castlevax, Amovir, Vivaldi Biosciences, Contrafect, 7Hills Pharma, Avimex, Pagoda, Accurius, Esperovax, Applied Biological Laboratories, Pharmamar, CureLab Oncology, CureLab Veterinary, Synairgen, Paratus, Pfizer and Prosetta, outside of the reported work. A.G.-S. has been an invited speaker in meeting events organized by Seqirus, Janssen, Abbott, Astrazeneca and Novavax. A.G.-S. is inventor on patents and patent applications on the use of antivirals and vaccines for the treatment and prevention of virus infections and cancer, owned by the Icahn School of Medicine at Mount Sinai, New York, outside of the reported work.

### Funding Statement

We acknowledge funding from NIH R01DK118222 and DoD W81XWH −20-1-0837 (E.U.A.). Azeloglu, Coca, Campbell, Schaub, and Kretzler are partially supported by Kidney Precision Medicine Project (KPMP), which maintains the Kidney Tissue Atlas of publicly available urine somascan and human transcriptomic data that were used in this study. The KPMP is funded by the following grants from the NIDDK: U01DK133081, U01DK133091, U01DK133092, U01DK133093, U01DK133095, U01DK133097, U01DK114866, U01DK114908, U01DK133090, U01DK133113, U01DK133766, U01DK133768, U01DK114907, U01DK114920, U01DK114923, U01DK114933, U24DK114886, UH3DK114926, UH3DK114861, UH3DK114915, UH3DK114937. L. Chan is supported in part by a grant from the NIH (K23DK124645 and U01DK137259). The mass spectrometry data were obtained from an Orbitrap mass spectrometer funded in part by the NIH grant S10OD025047, for the support of proteomics research at Rutgers Newark campus. This work was partially supported by CRIPT (Center for Research on Influenza Pathogenesis and Transmission), a NIAID funded Center of Excellence for Influenza Research and Response (CEIRR, contract # 75N93021C00014), and by NIAID grants U19AI142733, U19AI135972 and U19AI168631 to A.G-S., by the JPB and OPP foundations and an anonymous philanthropic donor to A.G-S. J. Haydak was supported by NIH T32HD075735 NICHD-Interdisciplinary Training in Systems and Developmental Biology and Birth Defects; A. Mendoza was supported by NIH R01DK131047 Diversity Supplement.

### Author Declarations

This study was approved by the Icahn School of Medicine at Mount Sinai Program Institutional Review Board (IRB) and the University of Michigan Medical School Institutional Review Board.

## References

1. WHO. WHO COVID-19 dashboard. https://data.who.int/dashboards/covid19/cases?m49=001&n=c

2. Nie X, Qian L, Sun R, et al. Multi-organ proteomic landscape of COVID-19 autopsies. Cell. Feb 4 2021;184(3):775–791 e14. doi:10.1016/j.cell.2021.01.004

3. Zaim S, Chong JH, Sankaranarayanan V, Harky A. COVID-19 and Multiorgan Response. Curr Probl Cardiol. Aug 2020;45(8):100618. doi:10.1016/j.cpcardiol.2020.100618

4. Puelles VG, Lutgehetmann M, Lindenmeyer MT, et al. Multiorgan and Renal Tropism of SARS-CoV-2. N Engl J Med. Aug 6 2020;383(6):590–592. doi:10.1056/NEJMc2011400

5. Hassler L, Reyes F, Sparks MA, Welling P, Batlle D. Evidence For and Against Direct Kidney Infection by SARS-CoV-2 in Patients with COVID-19. Clin J Am Soc Nephrol. Nov 2021;16(11):1755–1765. doi:10.2215/CJN.04560421

6. Nicosia RF. Kidney Disease and Viral Infection in COVID-19: Why Are Kidney Organoid and Biopsy Studies Not in Agreement? Nephron. 2023;147(8):458–464. doi:10.1159/000528460

7. Braun F, Lutgehetmann M, Pfefferle S, et al. SARS-CoV-2 renal tropism associates with acute kidney injury. Lancet. Aug 29 2020;396(10251):597–598. doi:10.1016/S0140-6736(20)31759-1

8. Chan L, Chaudhary K, Saha A, et al. AKI in Hospitalized Patients with COVID-19. J Am Soc Nephrol. Jan 2021;32(1):151–160. doi:10.1681/ASN.2020050615

9. Ye Y, Swensen AC, Wang Y, et al. A Pilot Study of Urine Proteomics in COVID-19-Associated Acute Kidney Injury. Kidney Int Rep. Dec 2021;6(12):3064–3069. doi:10.1016/j.ekir.2021.09.010

10. Schiffl H, Lang SM. Long-term interplay between COVID-19 and chronic kidney disease. Int Urol Nephrol. Feb 24 2023:1–8. doi:10.1007/s11255-023-03528-x

11. Ramamoorthy R, Hussain H, Ravelo N, et al. Kidney Damage in Long COVID: Studies in Experimental Mice. Biology (Basel). Jul 30 2023;12(8)doi:10.3390/biology12081070

12. Chavan S, Mangalaparthi KK, Singh S, et al. Mass Spectrometric Analysis of Urine from COVID-19 Patients for Detection of SARS-CoV-2 Viral Antigen and to Study Host Response. J Proteome Res. Jul 2 2021;20(7):3404–3413. doi:10.1021/acs.jproteome.1c00391

13. Bi X, Liu W, Ding X, et al. Proteomic and metabolomic profiling of urine uncovers immune responses in patients with COVID-19. Cell Rep. Jan 18 2022;38(3):110271. doi:10.1016/j.celrep.2021.110271

14. Li Y, Wang Y, Liu H, et al. Urine proteome of COVID-19 patients. Urine (Amst). 2020;2:1–8. doi:10.1016/j.urine.2021.02.001

15. Batra R, Uni R, Akchurin OM, et al. Urine-based multi-omic comparative analysis of COVID-19 and bacterial sepsis-induced ARDS. Mol Med. Jan 26 2023;29(1):13. doi:10.1186/s10020-023-00609-6

16. Richard VR, Gaither C, Popp R, et al. Early Prediction of COVID-19 Patient Survival by Targeted Plasma Multi-Omics and Machine Learning. Mol Cell Proteomics. Oct 2022;21(10):100277. doi:10.1016/j.mcpro.2022.100277

17. Paranjpe I, Jayaraman P, Su CY, et al. Proteomic characterization of acute kidney injury in patients hospitalized with SARS-CoV2 infection. Commun Med (Lond). Jun 12 2023;3(1):81. doi:10.1038/s43856-023-00307-8

18. Demichev V, Tober-Lau P, Lemke O, et al. A time-resolved proteomic and prognostic map of COVID-19. Cell Syst. Aug 18 2021;12(8):780–794 e7. doi:10.1016/j.cels.2021.05.005

19. Geyer PE, Arend FM, Doll S, et al. High-resolution serum proteome trajectories in COVID-19 reveal patient-specific seroconversion. EMBO Mol Med. Aug 9 2021;13(8):e14167. doi:10.15252/emmm.202114167

20. Vollmy F, van den Toorn H, Zenezini Chiozzi R, et al. A serum proteome signature to predict mortality in severe COVID-19 patients. Life Sci Alliance. Sep 2021;4(9)doi:10.26508/lsa.202101099

21. He Q, Mok TN, Yun L, He C, Li J, Pan J. Single-cell RNA sequencing analysis of human kidney reveals the presence of ACE2 receptor: A potential pathway of COVID-19 infection. Mol Genet Genomic Med. Oct 2020;8(10):e1442. doi:10.1002/mgg3.1442

22. Helms L, Marchiano S, Stanaway IB, et al. Cross-validation of SARS-CoV-2 responses in kidney organoids and clinical populations. JCI Insight. Dec 22 2021;6(24)doi:10.1172/jci.insight.154882

23. Jansen J, Reimer KC, Nagai JS, et al. SARS-CoV-2 infects the human kidney and drives fibrosis in kidney organoids. Cell Stem Cell. Feb 3 2022;29(2):217–231 e8. doi:10.1016/j.stem.2021.12.010

24. Siegerist F, Hay E, Dikou JS, et al. ScoMorphoFISH: A deep learning enabled toolbox for single-cell single-mRNA quantification and correlative (ultra-)morphometry. J Cell Mol Med. Jun 2022;26(12):3513–3526. doi:10.1111/jcmm.17392

25. Ritchie ME, Phipson B, Wu D, et al. limma powers differential expression analyses for RNA-sequencing and microarray studies. Nucleic Acids Res. Apr 20 2015;43(7):e47. doi:10.1093/nar/gkv007

26. Huang da W, Sherman BT, Lempicki RA. Systematic and integrative analysis of large gene lists using DAVID bioinformatics resources. Nat Protoc. 2009;4(1):44–57. doi:10.1038/nprot.2008.211

27. Chen EY, Tan CM, Kou Y, et al. Enrichr: interactive and collaborative HTML5 gene list enrichment analysis tool. BMC Bioinformatics. Apr 15 2013;14:128. doi:10.1186/1471-2105-14-128

28. Kuleshov MV, Jones MR, Rouillard AD, et al. Enrichr: a comprehensive gene set enrichment analysis web server 2016 update. Nucleic Acids Res. Jul 8 2016;44(W1):W90–7. doi:10.1093/nar/gkw377

29. Xie Z, Bailey A, Kuleshov MV, et al. Gene Set Knowledge Discovery with Enrichr. Curr Protoc. Mar 2021;1(3):e90. doi:10.1002/cpz1.90

30. Kursa MB, Rudnicki WR. Feature Selection with the Boruta Package. Journal of Statistical Software. 09/16 2010;36(11):1 - 13. doi:10.18637/jss.v036.i11

31. Knapp B, Wolfrum U. Adhesion GPCR-Related Protein Networks. Handb Exp Pharmacol. 2016;234:147–178. doi:10.1007/978-3-319-41523-9_8

32. Singh A, Winterbottom E, Daar IO. Eph/ephrin signaling in cell-cell and cell-substrate adhesion. Front Biosci (Landmark Ed). Jan 1 2012;17(2):473–97. doi:10.2741/3939

33. Bojesen KB, Clausen O, Rohde K, et al. Nectin-1 binds and signals through the fibroblast growth factor receptor. J Biol Chem. Oct 26 2012;287(44):37420–33. doi:10.1074/jbc.M112.345215

34. Irie K, Shimizu K, Sakisaka T, Ikeda W, Takai Y. Roles and modes of action of nectins in cell-cell adhesion. Semin Cell Dev Biol. Dec 2004;15(6):643–56. doi:10.1016/j.semcdb.2004.09.002

35. Bernardo BC, Belluoccio D, Rowley L, Little CB, Hansen U, Bateman JF. Cartilage intermediate layer protein 2 (CILP-2) is expressed in articular and meniscal cartilage and down-regulated in experimental osteoarthritis. J Biol Chem. Oct 28 2011;286(43):37758–67. doi:10.1074/jbc.M111.248039

36. Cheng G, Zhong M, Kawaguchi R, et al. Identification of PLXDC1 and PLXDC2 as the transmembrane receptors for the multifunctional factor PEDF. Elife. Dec 23 2014;3:e05401. doi:10.7554/eLife.05401

37. Guan Y, Du Y, Wang G, et al. Overexpression of PLXDC2 in Stromal Cell-Associated M2 Macrophages Is Related to EMT and the Progression of Gastric Cancer. Front Cell Dev Biol. 2021;9:673295. doi:10.3389/fcell.2021.673295

38. Fournier P, Dussault S, Fusco A, Rivard A, Royal I. Tyrosine Phosphatase PTPRJ/DEP-1 Is an Essential Promoter of Vascular Permeability, Angiogenesis, and Tumor Progression. Cancer Res. Sep 1 2016;76(17):5080–91. doi:10.1158/0008-5472.CAN-16-1071

39. Zacharias M, Kashofer K, Wurm P, et al. Host and microbiome features of secondary infections in lethal covid-19. iScience. Sep 16 2022;25(9):104926. doi:10.1016/j.isci.2022.104926

40. Luo YS, Li W, Cai Y, et al. Genome-wide screening of sex-biased genetic variants potentially associated with COVID-19 hospitalization. Front Genet. 2022;13:1014191. doi:10.3389/fgene.2022.1014191

41. Leng L, Cao R, Ma J, et al. Pathological features of COVID-19-associated lung injury: a preliminary proteomics report based on clinical samples. Signal Transduct Target Ther. Oct 15 2020;5(1):240. doi:10.1038/s41392-020-00355-9

42. Tan Y, Zhang W, Zhu Z, et al. Integrating longitudinal clinical laboratory tests with targeted proteomic and transcriptomic analyses reveal the landscape of host responses in COVID-19. Cell Discov. Jun 8 2021;7(1):42. doi:10.1038/s41421-021-00274-1

43. Bowe B, Xie Y, Xu E, Al-Aly Z. Kidney Outcomes in Long COVID. J Am Soc Nephrol. Nov 2021;32(11):2851–2862. doi:10.1681/ASN.2021060734

44. Xia T, Zhang W, Xu Y, et al. Early kidney injury predicts disease progression in patients with COVID-19: a cohort study. BMC Infect Dis. Sep 27 2021;21(1):1012. doi:10.1186/s12879-021-06576-9

45. Alexander MP, Mangalaparthi KK, Madugundu AK, et al. Acute Kidney Injury in Severe COVID-19 Has Similarities to Sepsis-Associated Kidney Injury: A Multi-Omics Study. Mayo Clin Proc. Oct 2021;96(10):2561–2575. doi:10.1016/j.mayocp.2021.07.001

46. Jayaraman P, Rajagopal M, Paranjpe I, et al. Peripheral Transcriptomics in Acute and Long-Term Kidney Dysfunction in SARS-CoV2 Infection. medRxiv. Oct 27 2023;doi:10.1101/2023.10.25.23297469

47. Gupta A, Al-Tamimi AO, Halwani R, Alsaidi H, Kannan M, Ahmad F. Lipocalin-2, S100A8/A9, and cystatin C: Potential predictive biomarkers of cardiovascular complications in COVID-19. Exp Biol Med (Maywood). Jul 2022;247(14):1205–1213. doi:10.1177/15353702221091990

48. Matuszewski M, Reznikov Y, Pruc M, et al. Prognostic Performance of Cystatin C in COVID-19: A Systematic Review and Meta-Analysis. Int J Environ Res Public Health. Nov 7 2022;19(21)doi:10.3390/ijerph192114607

49. Cheung MD, Erman EN, Liu S, et al. Single-Cell RNA Sequencing of Urinary Cells Reveals Distinct Cellular Diversity in COVID-19-Associated AKI. Kidney360. Jan 27 2022;3(1):28–36. doi:10.34067/KID.0005522021

50. Hinze C, Kocks C, Leiz J, et al. Single-cell transcriptomics reveals common epithelial response patterns in human acute kidney injury. Genome Med. Sep 9 2022;14(1):103. doi:10.1186/s13073-022-01108-9

51. Klocke J, Kim SJ, Skopnik CM, et al. Urinary single-cell sequencing captures kidney injury and repair processes in human acute kidney injury. Kidney Int. Dec 2022;102(6):1359–1370. doi:10.1016/j.kint.2022.07.032

52. Aregger F, Uehlinger DE, Witowski J, et al. Identification of IGFBP-7 by urinary proteomics as a novel prognostic marker in early acute kidney injury. Kidney Int. Apr 2014;85(4):909–19. doi:10.1038/ki.2013.363

53. Su L, Zhang J, Peng Z. The role of kidney injury biomarkers in COVID-19. Ren Fail. Dec 2022;44(1):1280–1288. doi:10.1080/0886022X.2022.2107544

54. Suvarna K, Salkar A, Palanivel V, et al. A Multi-omics Longitudinal Study Reveals Alteration of the Leukocyte Activation Pathway in COVID-19 Patients. J Proteome Res. Oct 1 2021;20(10):4667–4680. doi:10.1021/acs.jproteome.1c00215

55. Fang Y, Liu H, Huang H, et al. Distinct stem/progenitor cells proliferate to regenerate the trachea, intrapulmonary airways and alveoli in COVID-19 patients. Cell Res. Aug 2020;30(8):705–707. doi:10.1038/s41422-020-0367-9

56. Lechner M, Wojnar P, Redl B. Human tear lipocalin acts as an oxidative-stress-induced scavenger of potentially harmful lipid peroxidation products in a cell culture system. Biochem J. May 15 2001;356(Pt 1):129–35. doi:10.1042/0264-6021:3560129

57. Xu M, Yang W, Wang X, Nayak DK. Lung Secretoglobin Scgb1a1 Influences Alveolar Macrophage-Mediated Inflammation and Immunity. Front Immunol. 2020;11:584310. doi:10.3389/fimmu.2020.584310

58. Cao W, Feng Q, Wang X. Computational analysis of TMPRSS2 expression in normal and SARS-CoV-2-infected human tissues. Chem Biol Interact. Sep 1 2021;346:109583. doi:10.1016/j.cbi.2021.109583

59. Aroca-Martinez G, Avendano-Echavez L, Garcia C, et al. Renal tubular dysfunction in COVID-19 patients. Ir J Med Sci. Apr 2023;192(2):923–927. doi:10.1007/s11845-022-02993-0

60. Werion A, Belkhir L, Perrot M, et al. SARS-CoV-2 causes a specific dysfunction of the kidney proximal tubule. Kidney Int. Nov 2020;98(5):1296–1307. doi:10.1016/j.kint.2020.07.019

61. Baird SK, Kurz T, Brunk UT. Metallothionein protects against oxidative stress-induced lysosomal destabilization. Biochem J. Feb 15 2006;394(Pt 1):275–83. doi:10.1042/BJ20051143

62. Larsen CP, Wickman TJ, Braga JR, et al. APOL1 Risk Variants and Acute Kidney Injury in Black Americans with COVID-19. Clin J Am Soc Nephrol. Dec 2021;16(12):1790–1796. doi:10.2215/CJN.01070121

63. Nystrom SE, Li G, Datta S, et al. JAK inhibitor blocks COVID-19 cytokine-induced JAK/STAT/APOL1 signaling in glomerular cells and podocytopathy in human kidney organoids. JCI Insight. Jun 8 2022;7(11)doi:10.1172/jci.insight.157432

