## Supplementary materials for "Integrated multiomics implicates dysregulation of ECM and cell adhesion pathways as drivers of severe COVID-associated kidney injury"

#### Supplementary methods

All acute COVID-19 samples from hospitalized participants were collected between April 2020 and April 2021. All non-COVID AKI samples from hospitalized participants were collected between May 2021 and August 2021. AKI was defined by KDIGO creatinine criteria. Urine specimen was collected in sterile midstream specimen collection container (ThermoScientific, Cat. No. 070020). For the instrument controls, we pooled urine from 400 (100 healthy, 100 AKI, 100 CKD and 100 AKI+CKD) samples from the ASSESS AKI cohort [1]. Samples were processed according to the workflow shown in **Figure S29**. Since samples were assessed over the course of several months during the pandemic from two different medical centers, triplicates of a singular instrument control were used in each run to account for LC-MS instrument variability and batch to batch variation. Each of the TMT-16 plex contained thirteen samples in addition to three instrument controls. Estimated glomerular filtration rate (eGFR) of the COVID-19 samples was calculated using the peak serum creatinine values as per the recommendations published previously [2].

*Urine processing at Mount Sinai Hospital*

Urine was processed in a BSL2+ facility according to guidelines provided by the Institutional Biosafety committee (IBC) at the Mount Sinai Hospital. Urine was spun down at 2000 rpm for 15 minutes at 4°C and the supernatant was aliquoted and stored at -80°C until use or for long term storage. For samples that were not immediately processed, urine was snap frozen in dry ice and stored at -80°C until ready to use.

*Urine processing at University of Michigan*

Urine was spun down at 200xg for 15 minutes at 4°C and the supernatant was filtered using a 30-micron mesh filter. Urine supernatants were then aliquoted and stored at -80°C until use or for long term storage. 8 samples that were collected more than 18 days after hospitalization were excluded from the analysis.

*Urine processing for proteomics*

Urine protein was precipitated in a BSL2+ facility using a previously published protocol [3]. Briefly, urine supernatant is mixed with acetone (Sigma, Cat. No. 179124) and trichloroacetic acid (TCA, Sigma, Cat. No. 91228) at a ratio of 1:8:1 (Urine:Acetone:TCA). After incubation for 1h at -20°C, the samples were spun down at 4000xg for 1 hour at 4°C. The supernatant was disinfected according to biosafety guidelines and discarded, and the urine protein pellet was washed twice with ice-cold acetone to remove TCA and centrifuged at 12,000xg for 15 minutes. The pellet was air dried for 15 minutes, resuspended in rehydration buffer containing 7M Urea (Sigma, Cat. No. U5378), 2M Thiourea (Sigma Cat. No. T8656), 50 mM Dithiothreitol (DTT, Thermofisher, Cat. No. R0861) and 1% CHAPS (Thermofisher, Cat. No. 28300). Protein content was quantified using Bradford assay (Sigma, Cat. No. B6916) against BSA standards, and aliquots were stored at -80°C until further use.

For proteomic analysis, urine protein samples boiled with LDS sample buffer (Life Technologies, Cat. No. B0007) and then loaded on SDS-PAGE gels (Life Technologies, Cat. No. NW4122BOX) at 30 µg protein per lane. After protein separation, the protein gels were fixed in 50% methanol (Fisher Scientific, Cat. No. A412-4) and 10% acetic acid (Fisher Scientific, Cat. No. A38-212) for 30 minutes and then stained with Coomassie R-250 (BioRad, Cat. No. 1610436) for an hour. The gels were further processed for isobaric tagged TMT-16 plex proteomics at the Center for Advanced Proteomics research at Rutgers University.

*Isobaric TMT-16 Plex LC-MS/MS analysis*

The gels were cut into small blocks, washed with a solution of 30% acetonitrile (ACN) and 70 % of 100 mM triethylammonium bicarbonate (TEAB) to remove contaminants, and treated with 25 MM dithiothreitol (DTT, at 55 °C for 30 min) and then with 50 mM iodoacetamide (IAM, at room temperature in the dark for 30 min) to reduce disulfides and alkylate thiols, respectively. The gel blocks were then dehydrated with ACN (9 mL) to remove DTT and IAM, and trypsin was added for in-gel digestion, which was carried out at 37 °C for 16 hours. To extract the ensuing peptides, 1 mL of 0.1% trifluoroacetic acid was added, followed by 1 mL of ACN three times. The peptide solution was dried using a Speedvac. The peptides from each sample were then labeled with one of the TMT-16 tags (ThermoScientific, Cat. No. A44520) as per the manufacturer's instructions for 2 hours at room temperature. After neutralizing the remaining TMT reagents with hydroxylamine, the TMT-labeled peptides were mixed and separated using high pH RPLC chromatography (Waters XBridge BEH $C_{18}$ column, 5 µm, 4.6x250 mm). Sixty 1-min fractions were collected and combined into 12 fractions. Each fraction was dried using a Speedvac and purified using $C_{18}$ cartridges.

LC-MS/MS analysis of the peptides was performed using a Tribrid Fusion Lumos Orbitrap Mass Spectrometer with an UltiMate 3000 RSLC nano system from Thermo Scientific. Peptides were separated on an Acclaim PepMap $C_{18}$ column (75 µm × 50 cm, 2 µm, 100 Å) using a 3H binary gradient, from 2 to 95% of Solvent B (85% ACN in 0.1% FA), at a flow rate of 300 nL/min. The eluted peptides were then introduced into the MS system for data-dependent analysis in positive ion mode. Full scans of the peptides were got in an m/z range of 350 to 1600 with a resolution of 120,000, using a maximal injection time of 50 ms. The MS/MS scan used a quadrupole isolation window of 0.7 m/z, with HCD being used for peptide fragmentation and a normalized HCD collision energy of 34%. The peptide fragments were detected in the Orbitrap analyzer at a resolution of 50,000, with the AGC target set to a normalized target of 150% and the dynamic maximum injection time selected. The dynamic exclusion was set to 60 seconds.

The tandem spectra were analyzed using the SEQUEST search engine through the Proteome Discoverer platform (version 2.4, Thermo Scientific) and matched to the Uniprot human database (which was updated on 11/12/2020 and contained 74,848 entries). TMT $MS^2$ reporter-based quantitation method was used for protein identification and measurement, with methionine oxidation set as a dynamic modification and cysteine carbamidomethylation, TMT labeling of peptide N-termini, and lysine side chains set as static modifications. The Percolator module was used to validate peptide identification with a false discovery rate of less than 1%. Protein quantification was normalized based on the total amounts of peptides and included both unique and razor peptides.

*ML training and validation parameters*

Eight COVID-19 samples collected more than 18 days after hospitalization were excluded from ML analysis. Participants with reported clinical outcomes of admission to the intensive care unit (ICU), mechanical ventilation (MV), acute kidney injury (AKI), incidence after hospitalization, length of stay over 21 days (LOS>21) and/or death were considered severe outcome. The samples were first randomly divided into the discovery set and the validation set in a 2:1 ratio while maintaining a similar proportion of severe outcomes cases. The discovery set was used to derive prediction set of genes and the validation set was untouched during training process and was only used for validation of composite outcome prediction. Limma test, as described previously [4], was first performed in the discovery set to obtain differential abundant proteins between severe and mild outcomes cases. Then we selected important proteins (features) with respect to the composite outcome (severe vs mild) using Boruta feature selection method [5]. The selected proteins (features) were used for random forest model construction within the discovery set. Parameter "mtry" was tuned using grid search and "ntree" were tuned within range [500,1000,1500,2000,2500] via 10-fold cross validation, and were determined by the highest accuracy (mtry=1, ntree=500). Final random forest model was applied to both the discovery set and the validation set for receiver operating characteristic curves (ROC)/Area under the ROC curve (AUC) calculation. The cut off was determined in the discovery set by the nearest distance between ROC curve and point (0.1) and applied to the validation set to obtain True Positive Rate (TPR), False Positive Rate (FPR), Positive Predictive Value (PPV), and Negative Predictive Value (NPV).

*Plasma Proteomics analysis*

As defined in a previously published manuscript [6, 7], blood collected in CPT tubes were centrifuged within two hours of blood collection at room temperature for 15 minutes at 1500-1800 RCF (Relative Centrifugal Force). The plasma layer (under the top whitish layer of mononuclear cells and platelets) was aspirated without disturbing the cell layer, aliquoted and stored at −80 °C. AKI was defined according to Kidney Disease Improving Global Outcomes (KDIGO) criteria, with baseline creatinine determined based on previous measurements or eGFR. Clinical data, including demographic and laboratory information, were obtained from institutional electronic health records (EHR). Disease severity was assessed by supplemental oxygen requirement. The SomaScan platform was employed for proteomic analysis, quantifying protein expression levels [8]. Standard preprocessing protocols, including normalization and calibration, were applied to the data, ensuring quality control and removal of samples with intrinsic issues. The relative fluorescence unit (RFU) values were log2 transformed for analysis.

For prevalent acute kidney injury (AKI) analysis, log2 transformed normalized protein values were subjected to multivariable linear regression using the Limma framework. Models were adjusted for age, sex, history of chronic kidney disease (CKD), and supplemental oxygen requirement at the time of specimen collection, as defined previously [6]. P-values were adjusted using the Benjamin-Hochberg procedure to control the false discovery rate (FDR) at 5%. This research was reviewed and approved by the Icahn School of Medicine at Mount Sinai Program for the Protection of Human Subjects (PPHS) under study number 20-00341.

*Urine sediment scRNA sequencing*

Cells isolated from urine sediments were further processed using single-cell sequencing analysis, as reported previously [9]. Briefly, cell pellet after removal of urine supernatant was washed with 1 ml cold X-VIVO™10 medium once (Cat#: 04-743Q, Lonza) and then centrifuged for 5 minutes at 200 x g at 4°C. The cell pellet was then resuspended in 50 µl DMEM/F12 medium supplemented with HEPES and 10% (v/v) FBS. About 50,000 cells were loaded onto the single-cell droplet based RNAseq platform and next generation sequencing was carried out on a NovaSeq6000 (Illumina) machine generating a 200 million reads per sample.

*Kidney Organoid Generation*

Induced pluripotent stem cell (iPSC) line ISMMS 102i used in this study is publicly available from WiCell research institute. Kidney organoids were generated from 102i using a modified protocol adapted from Takasato et al [10] (**Figure S30**). Briefly, iPSCs were first induced towards intermediate mesoderm (IM) cells in 2D culture by activating the Wnt signaling pathway using 7μM CHIR (Stemgent, Cat No. 04–0004) in STEMdiff APEL2 Medium (Stem Cell Technologies, Cat. No. 05275) medium for 4 days, followed by switching to APEL2 with 200 ng/mL FGF-9 (R&D, Cat No. 273-F9-025), 1μM CHIR and 1 μg/mL Heparin (Sigma, Cat No. H4784). Deviant from the original protocol, after 7 days in culture, the cells were seeded in a U-bottom 96-well plate (Life Technologies, Cat No. 174925) at a density of 25,000 cells per well. The spheroids were cultured in APEL2 with 200 ng/mL FGF-9, 1μM CHIR and 1 μg/mL Heparin until day 5. After 5 days, growth factors were withdrawn, and the organoids were grown in APEL2 medium with 1 μg/mL Heparin for 13 days at which point they are ready to be used for further experiments.

*SARS-CoV-2 infection of Kidney organoids*

Kidney organoids differentiated for 25 days were infected with SARS-CoV-2, isolate USA-WA1/2020 (BEI Resources NR-52281) under BSL3 containment in accordance with the biosafety protocols developed by the Icahn School of Medicine at Mount Sinai. Virus particles at $10^4$ PFU were used for infection and media supernatants were collected at days 2, 4 and 6 post infection for viral titer quantification using TCID50 assay, as previously described [11, 12].

*Whole mount Immunofluorescent staining*

Organoids were moved to an optical bottom 96-well plate, fixed at RT for 1 hour with 4% paraformaldehyde (PFA) (Electron Microscopy Sciences, Cat No. 15710), and blocked at RT for 3h using blocking buffer consisting of 10% bovine serum albumin (Sigma, Cat No. A8806), 10% donkey serum (Sigma, Cat No. S30) and 0.3% Triton-X (Sigma, Cat No. X100). After 3h, blocking buffer was replaced with primary antibodies prepared at dilutions listed in **Table S4** and incubated overnight at 4°C. The organoids were then washed two times with PBS for 1h each and a third wash for 3h. The corresponding secondary antibodies (dilution 1:250) and Lotus Tetragonolobus Lectin (LTL, 1:200) (Vector Laboratories Cat. No. FL-1321-2) were added and incubated at RT for 2 hours. Samples were washed for 30 minutes three times with PBS and then Hoechst 33342 (ThermoScientific, Cat No. 62249) was added at 1:5,000 dilution at RT for 30 minutes. The stain was again replaced thrice with PBS and imaged in an optical bottom plate or a glass bottom dish (Cellvis Cat No. D35-14-1.5N) using Zeiss LSM780 laser scanning confocal microscope on 20X water immersion objective.

*Kidney organoid scRNA-seq*

Kidney organoids after day 4 of infection were dissociated using Accutase (Stem Cell Technologies, Cat No. 07920). The single-cell suspension obtained from 7 organoids were pooled into one sample and 6000 cells were loaded onto the 10X Chromium Next GEM Chip (10X Genomics, Cat No. 1000127) per manufacturer's instructions. After GEM preparation in a Chromium controller, library construction was carried out using 10X Chromium Next GEM single-cell library kit (10X Genomics, Cat. No. 1000165). scRNA-seq was performed on a NovaSeq6000 (Illumina) machine generating a 200 million reads per sample.

*scRNA-seq data analysis*

The raw sequencing data was processed with Cellranger count (version 5.0.0) [13]. To quantify COVID gene expression, a reference genome was built with Cellranger mkref combining human hg38 and SARS-CoV-2 from the Ensembl database. The gene-cell count matrix was subject to Seurat package for quality control (QC), normalization, clustering, and visualization [14]. Genes expressed in less than 3 cells were considered as noise and removed from further analysis. Cells with < 200 genes expressed or with > 50 precent transcripts from mitochondrial genes were considered as low quality and removed, while cells with > 5000 genes expressed were considered as doublets and filtered out. The expression profile after QC was normalized with the read depth for each cell and multiplied by the scale factor 10,000, then log transformed. To investigate the organoid tissue similarities, COVID genes were excluded for clustering analysis. The KNN graph was built with the first 15 PCAs with FindNeighbors and unsupervised clustering was conducted by FindClusters using Louvain algorithm with resolution 0.8. The clusters were further annotated with the markers in previous organoid single cell studies [15]. The differentially expressed genes (DEG) between infected and non-infected sample of each cell was identified by FindMarker implemented in Seurat package and genes with p-value <= 0.01 were considered as significantly dysregulated.

*KPMP analysis*

Fully annotated scRNA-seq data (file KidneyTissueAtlas/521c5b34-3dd0-4871-8064-61d3e3f1775a_PREMIERE_Alldatasets_08132021.h5Seurat) was downloaded from the KPMP Kidney Tissue Atlas (https://atlas.kpmp.org/). Cell identity was assigned based on the "subclass.l1" classifications. To determine the specificity of each gene within kidney cell types, the average expression for each cell type was calculated and divided by the sum of average expression for all cell types.

The validation of the urinary features used in this study in the KPMP AKI cohort was determined from publicly available and normalized urinary SomaScan proteomics downloaded from https://atlas.kpmp.org/ (6696930a-f707-430d-a964-

110aefa93c62_Urine Biomarker Data-SomaScan-2022\Urine Biomarker Data-SomaScan-2022\Data\SS-2342467_2023-11-30_Urine.ANMLNormalized.xlsx). Patient identifiers in the SomaScan data was matched to the corresponding clinical data (also publicly available from https://atlas.kpmp.org/; 1d64f325-eca1-4a9f-a89a-6b62b96c4d28_20231129_OpenAccessClinicalData.csv) to identify patients as belonging to the healthy reference or AKI group. For each feature, the fold change was calculated by taking the average abundance value in all AKI patients divided by the average abundance in all healthy reference patients.

*Integrated multiomic data analysis*

Functional enrichment analysis of DAPs/DEGs were performed by Enrichr package [16-18]. DAPs/DEGs were first ranked by the absolute value of log (fold change) and top features with a maximum p-value of 0.05 were used for subsequent analysis. Enriched biological functions were determined by fisher-exact test using the information of biological process category in Gene Ontology (GO) [19]. Top DAPs and DEGs from multiple datasets employed in this study were analyzed using the standard enrichment module in MBCO to visualize functionally overlapping pathways [20]. The top 25% SCPs were considered for all analyses and functionally related SCPs were connected by a dashed line. All MBCO networks were visualized using hierarchical clustering in yED (https://www.yworks.com/products/yed). The kidney specific functional network in the Humanbase interface (https://hb.flatironinstitute.org/) was used to identify tissue specific functional neighborhood networks [21].

The top 150 DAPs/DEGs from the urine proteomics, urine sediment and kidney organoid scRNA-seq datasets and the top 450 DAPs from plasma proteomics dataset were used to ensure a similar proportion of the proteome is employed for the functional overlap analysis. The plots were generated using ggplot in R. Protein–protein interaction (PPI) network was constructed using the Network X package in Python v3.4.10 to display a Minimum Spanning Tree (MST) using Prim's algorithm. Top 50 urine sediment scRNA-seq DEGs and plasma and urinary proteomics DAPs were ranked by log2 fold-change and at 5% FDR to create the interaction network. Protein interactions were annotated from the STRING database to build a functional network [22]. We called this network the 'truth set'. Permutation testing of this set against randomized network cluster of 50 DEGs from urine sediment scRNA-seq, 50 plasma and 50 urine DAPs and was conducted along with hypothesis testing at a 5% FDR threshold. A histogram of the mean values of the permuted network scores was plotted along with the mean value of the truth set and two-tailed cutoff to identify significantly enriched network clusters from the truth set.

**List Supplementary Figures**

**Figure S1:** Levey-Jennings plot of number of proteins identified in each TMT-16 Plex set of the urine proteomics

**Figure S2**: Principal component analysis (PCA) plots of instrument controls and COVID-19 samples across (A) sample collection site and (B) TMT-16 plex set

**Figure S3**: Gene Ontology biological processes (GOBP) enrichment analysis heatmap of top 150 (A) upregulated and (B) downregulated differentially abundant proteins (DAPs) in COVID-19 vs healthy urine samples

**Figure S4**: GOBP enrichment heatmap of top 150 (A) total and (B) downregulated DAPs in severe vs mild COVID-19 urine samples

**Figure S5**: Correlation of prediction accuracy and (A) number of days hospitalized at urine collection; (B) outcome

**Figure S6**: GOBP enrichment analysis of the 12 features from urine proteomics

**Figure S7**: Kidney cell type specificity heatmap of ML features from the Kidney Precision Medicine Project (KPMP) database scRNA-seq dataset

**Figure S8:** Estimated glomerular filtration rate (eGFR) in severe versus mild

**Figure S9:** (A) Uniform Manifold Approximation and Projection (UMAP) on cosine distance metric; (B) K-means clustering of unscaled principal component analysis (PCA) plot; (C) Stacked bar graph of distribution of outcomes in each cluster

**Figure S10**: GOBP enrichment analysis heatmap of top 150 (A) upregulated and (B) downregulated DAPs in non-COVID AKI vs healthy urine samples

**Figure S11**: Abundance of (A) Albumin and (B) Cystatin C in healthy, mild and severe COVID-19 and non-COVID AKI urine samples

**Figure S12:** Log2 fold-change urinary abundance of ML features in AKI samples from an external dataset from the KPMP database

**Figure S13**: Interaction of top 150 urine and top 450 plasma proteomics DAPs analyzed using Molecular-Biology-of-the-Cell-Ontology (MBCO) standard enrichment. Pathways among top 25% interaction were connected by a dashed line

**Figure S14**: HumanBase kidney-specific functional module discovery analysis of top 150 DAPs from urine and 450 DAPs from plasma proteomics datasets

**Figure S15**: Kidney cell type specificity heatmap from scRNA-seq dataset in the KPMP database of (A) cell adhesion (B) cell migration and (C) response to oxidative stress associated genes from the functional overlap analysis

**Figure S16**: Histogram of the mean values of the permuted network scores along with original interaction score (red) of top 50 differentially expressed genes (DEGs) from individual cell clusters in the urine sediment single-cell RNA sequencing (scRNA-seq) and top 50 DAPs from urine and plasma

**Figure S17**: GOBP enrichment heatmap of DEGs in severe vs mild urine sediment scRNA-seq dataset LOH/DCT/CNT/PC cluster (hybrid cluster of loop of Henle, distal tubule, connecting tubule and principal cells)

**Figure S18**: Interaction of top 150 DAPs from Urine and top 450 DAPs from plasma proteomics with top 150 DEGs from the Urine sediment scRNA-seq LOH cluster analyzed using MBCO standard enrichment. Pathways among top 25% interaction were connected by a dashed line

**Figure S19**: HumanBase kidney-specific functional module discovery analysis of top 150 DAPs from urine and top 450 from plasma proteomics and top 150 DEGs from the urine sediment scRNA-seq LOH/DCT/CNT/PC cluster

**Figure S20**: HumanBase kidney-specific functional module discovery analysis of overlapping proteins from the protein-protein interaction (PPI) network of urine and plasma proteomics datasets and urine scRNAseq LOH/DCT/CNT/PC cluster

**Figure S21:** Viral titer quantification using supernatants from infected organoids on 2-, 4- and 6-days post infection (dpi)

**Figure S22**: Cell type clusters identified from the kidney organoid scRNA-seq

**Figure S23**: GOBP enrichment heatmap of top 150 DEGs in infected vs mock kidney organoids

**Figure S24**: Interaction of top 150 DEGs from kidney organoid scRNAseq and the top 150 DAPs from urine and the top 450 DAPs from plasma proteomics datasets analyzed using MBCO standard enrichment. Pathways among the top 25% interaction were connected by a dashed line

**Figure S25**: HumanBase kidney-specific functional module discovery analysis of top 150 DAPs from urine and plasma proteomics and top 150 DEGs from the kidney organoid scRNAseq dataset

**Figure S26**: Abundance of TMPRSS2 in mild and severe COVID-19 samples

**Figure S27:** Receiver operating characteristics (ROC) curves of COVID-19 severity prediction using Mount Sinai Hospital cohort as (A) discovery set and University of Michigan cohort as (B) validation set

**Figure S28:** (A) List of ML features identified and (B) GOBP enrichment analysis of biomarkers from ML model using Sinai cohort as discovery set and Michigan cohort as validation set

**Figure S29:** Schematic showing urine sample processing workflow for urinary proteomics

**Figure S30:** Schematic of iPSC-derived kidney organoid generation

#### Supplementary Tables

**Table S1:** Demographics of severe and mild COVID-19 samples

| **Demographics** | **Total** | **Mild** | **Severe** |
|---|---|---|---|
| Age, year, mean ± SD | 58 ± 15 | 57 ± 14 | 58 ± 17 |
| Sex, female/total (%) | 39.3 | 35.8 | 46 |
| Race, n/total (%) | | | |
| Asian | 5.73 | 6.17 | 4.87 |
| Black | 27.87 | 30.86 | 21.95 |
| White | 40.16 | 35.8 | 48.78 |
| Hispanic | 0.8 | 1.23 | 0 |
| Not reported | 25.4 | 24.69 | 26.82 |
| Baseline eGFR (ml/min/1.73 $m^2$), mean ± SD | 71.67 ± 33.68 | 79.1 ± 29.1 | 57 ± 37.9 |

**Table S2:** Features identified using the Boruta feature selection method

| **Accession** | **Importance** | **Gene Symbol** | **Description** | **p** | **p.adj** | **Log2FC** | **Odds Ratio** |
|---|---|---|---|---|---|---|---|
| O94910 | 7.345229152 | ADGRL1 | Adhesion G protein-coupled receptor L1 | 2.83E-05 | 0.013060871 | -0.417566734 | 0.08690704 |
| O43490 | 6.823453492 | PROM1 | Prominin-1 | 3.06E-05 | 0.013060871 | -0.389791982 | 0.040413 |
| A0A0A0MSQ0 | 6.516856314 | PLS3 | Plastin-3 | 0.024108612 | 0.157166064 | -0.234264162 | 0.0986646 |
| Q15223 | 4.85202059 | NECTIN1 | Nectin-1 | 0.000294101 | 0.020858332 | -0.35681949 | 0.14985783 |
| B1AKC9 | 4.810676631 | EPHB2 | Receptor protein-tyrosine kinase | 0.000877962 | 0.025106991 | -0.313390911 | 0.08481353 |
| P11684 | 4.250028861 | SCGB1A1 | Uteroglobin | 0.017870519 | 0.141167197 | 0.674266899 | 1.36289063 |
| Q6UX71 | 4.111293195 | PLXDC2 | Plexin domain-containing protein 2 | 0.000569995 | 0.022126152 | -0.321354845 | 0.02553036 |
| P31025 | 3.740521181 | LCN1 | Lipocalin-1 | 0.005569905 | 0.07161001 | 0.543814933 | 2.00956486 |
| Q9BQ51 | 3.533051227 | PDCD1LG2 | Programmed cell death 1 ligand 2 | 0.000451324 | 0.020858332 | -0.32479881 | 0.09988319 |
| K7EPJ4 | 3.388225058 | CILP2 | Cartilage intermediate layer protein 2 | 6.81E-05 | 0.0145382 | -0.504393948 | 0.08570248 |
| Q8WW52 | 3.259612754 | FAM151A | Protein FAM151A | 0.002159912 | 0.038554912 | -0.378602379 | 0.22040379 |

| | | | | | | | |
|---|---|---|---|---|---|---|---|
| A0A087WVC6 | 3.154126434 | PTPRJ | Protein-tyrosine-phosphatase | 0.000456222 | 0.020858332 | -0.192464054 | 0.06666445 |

**Table S3:** Prediction accuracy of ML algorithm using Mount Sinai Hospital as discovery cohort and University of Michigan as validation cohort

| Site | AUC | Sensitivity | Specificity | PPV | NPV | TP | TN | FP | FN |
|---|---|---|---|---|---|---|---|---|---|
| Mount Sinai Hospital - Discovery set | 0.874 | 0.5 | 0.949 | 0.733 | 0.872 | 11 | 75 | 4 | 11 |
| University of Michigan - Validation set | 0.599 | 0.154 | 0.929 | 0.8 | 0.371 | 4 | 13 | 1 | 22 |

**Table S4**: List of Antibodies used for immunofluorescence staining of kidney organoids

| Antibodies | Species | Vendor | Catalog # | Dilution |
|---|---|---|---|---|
| E-cadherin (CDH1) | Mouse | BD Biosciences | 610181 | 1:200 |
| Uromodulin (UMOD) | Sheep | R&D Systems | AF5144 | 1:250 |
| Synaptopodin (SYNPO) | Mouse | Progen | 65294 | 1:400 |
| Podocalyxin (PODXL) | Goat | Novus Biologicals | AF1658 | 1:200 |
| Angiotensin converting enzyme 2 (ACE2) | Goat | R&D Systems | AF933 | 1:400 |
| SARS-CoV-2 Nucleocapsid (N) | Mouse | Center for Therapeutic Antibody Discovery, ISMMS | 1C7C7 | 1:500 |

### References


1. Go AS, Parikh CR, Ikizler TA, et al. The assessment, serial evaluation, and subsequent sequelae of acute kidney injury (ASSESS-AKI) study: design and methods. *BMC Nephrol*. Aug 27 2010;11:22. doi:10.1186/1471-2369-11-22
2. Delgado C, Baweja M, Crews DC, et al. A Unifying Approach for GFR Estimation: Recommendations of the NKF-ASN Task Force on Reassessing the Inclusion of Race in Diagnosing Kidney Disease. *Am J Kidney Dis*. Feb 2022;79(2):268-288 e1. doi:10.1053/j.ajkd.2021.08.003
3. Beretov J, Wasinger VC, Schwartz P, Graham PH, Li Y. A standardized and reproducible urine preparation protocol for cancer biomarkers discovery. *Biomark Cancer*. 2014;6:21-7. doi:10.4137/BIC.S17991
4. Ritchie ME, Phipson B, Wu D, et al. limma powers differential expression analyses for RNA-sequencing and microarray studies. *Nucleic Acids Res*. Apr 20 2015;43(7):e47. doi:10.1093/nar/gkv007

5. Kursa MB, Rudnicki WR. Feature Selection with the Boruta Package. *Journal of Statistical Software*. 09/16 2010;36(11):1 - 13. doi:10.18637/jss.v036.i11
6. Del Valle DM, Kim-Schulze S, Huang HH, et al. An inflammatory cytokine signature predicts COVID-19 severity and survival. *Nat Med*. Oct 2020;26(10):1636-1643. doi:10.1038/s41591-020-1051-9
7. Paranjpe I, Jayaraman P, Su CY, et al. Proteomic characterization of acute kidney injury in patients hospitalized with SARS-CoV2 infection. *Commun Med (Lond)*. Jun 12 2023;3(1):81. doi:10.1038/s43856-023-00307-8
8. Williams SA, Kivimaki M, Langenberg C, et al. Plasma protein patterns as comprehensive indicators of health. *Nat Med*. Dec 2019;25(12):1851-1857. doi:10.1038/s41591-019-0665-2
9. Menon R, Otto EA, Sealfon R, et al. SARS-CoV-2 receptor networks in diabetic and COVID-19-associated kidney disease. *Kidney Int*. Dec 2020;98(6):1502-1518. doi:10.1016/j.kint.2020.09.015
10. Takasato M, Er PX, Chiu HS, Little MH. Generation of kidney organoids from human pluripotent stem cells. *Nat Protoc*. Sep 2016;11(9):1681-92. doi:10.1038/nprot.2016.098
11. Amanat F, White KM, Miorin L, et al. An In Vitro Microneutralization Assay for SARS-CoV-2 Serology and Drug Screening. *Curr Protoc Microbiol*. Sep 2020;58(1):e108. doi:10.1002/cpmc.108
12. Miorin L, Kehrer T, Sanchez-Aparicio MT, et al. SARS-CoV-2 Orf6 hijacks Nup98 to block STAT nuclear import and antagonize interferon signaling. *Proc Natl Acad Sci U S A*. Nov 10 2020;117(45):28344-28354. doi:10.1073/pnas.2016650117
13. Zheng GX, Terry JM, Belgrader P, et al. Massively parallel digital transcriptional profiling of single cells. *Nat Commun*. Jan 16 2017;8:14049. doi:10.1038/ncomms14049
14. Hao Y, Hao S, Andersen-Nissen E, et al. Integrated analysis of multimodal single-cell data. *Cell*. Jun 24 2021;184(13):3573-3587 e29. doi:10.1016/j.cell.2021.04.048
15. Subramanian A, Sidhom EH, Emani M, et al. Single cell census of human kidney organoids shows reproducibility and diminished off-target cells after transplantation. *Nat Commun*. Nov 29 2019;10(1):5462. doi:10.1038/s41467-019-13382-0
16. Kuleshov MV, Jones MR, Rouillard AD, et al. Enrichr: a comprehensive gene set enrichment analysis web server 2016 update. *Nucleic Acids Res*. Jul 8 2016;44(W1):W90-7. doi:10.1093/nar/gkw377
17. Chen EY, Tan CM, Kou Y, et al. Enrichr: interactive and collaborative HTML5 gene list enrichment analysis tool. *BMC Bioinformatics*. Apr 15 2013;14:128. doi:10.1186/1471-2105-14-128
18. Xie Z, Bailey A, Kuleshov MV, et al. Gene Set Knowledge Discovery with Enrichr. *Curr Protoc*. Mar 2021;1(3):e90. doi:10.1002/cpz1.90
19. Huang da W, Sherman BT, Lempicki RA. Systematic and integrative analysis of large gene lists using DAVID bioinformatics resources. *Nat Protoc*. 2009;4(1):44-57. doi:10.1038/nprot.2008.211
20. Hansen J, Meretzky D, Woldesenbet S, Stolovitzky G, Iyengar R. A flexible ontology for inference of emergent whole cell function from relationships between subcellular processes. *Sci Rep*. Dec 18 2017;7(1):17689. doi:10.1038/s41598-017-16627-4

21. Greene CS, Krishnan A, Wong AK, et al. Understanding multicellular function and disease with human tissue-specific networks. *Nat Genet*. Jun 2015;47(6):569-76. doi:10.1038/ng.3259
22. Szklarczyk D, Kirsch R, Koutrouli M, et al. The STRING database in 2023: protein-protein association networks and functional enrichment analyses for any sequenced genome of interest. *Nucleic Acids Res*. Jan 6 2023;51(D1):D638-D646. doi:10.1093/nar/gkac1000

**Figure S1:** Levey-Jennings plot of number of proteins identified in each TMT-16 Plex set of the urine proteomics

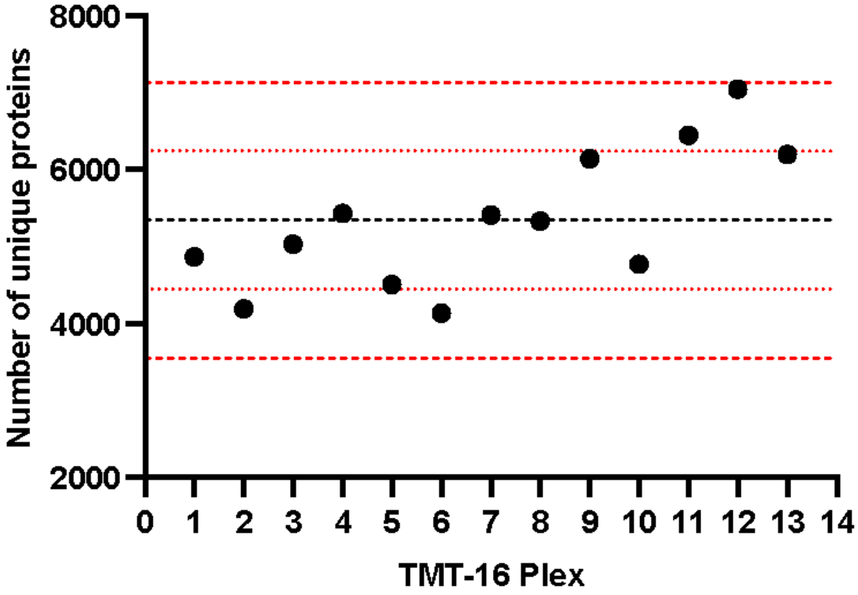

**Figure S2**: Principal component analysis (PCA) plots of instrument controls and COVID-19 samples across (A) sample collection site and (B) TMT-16 plex set

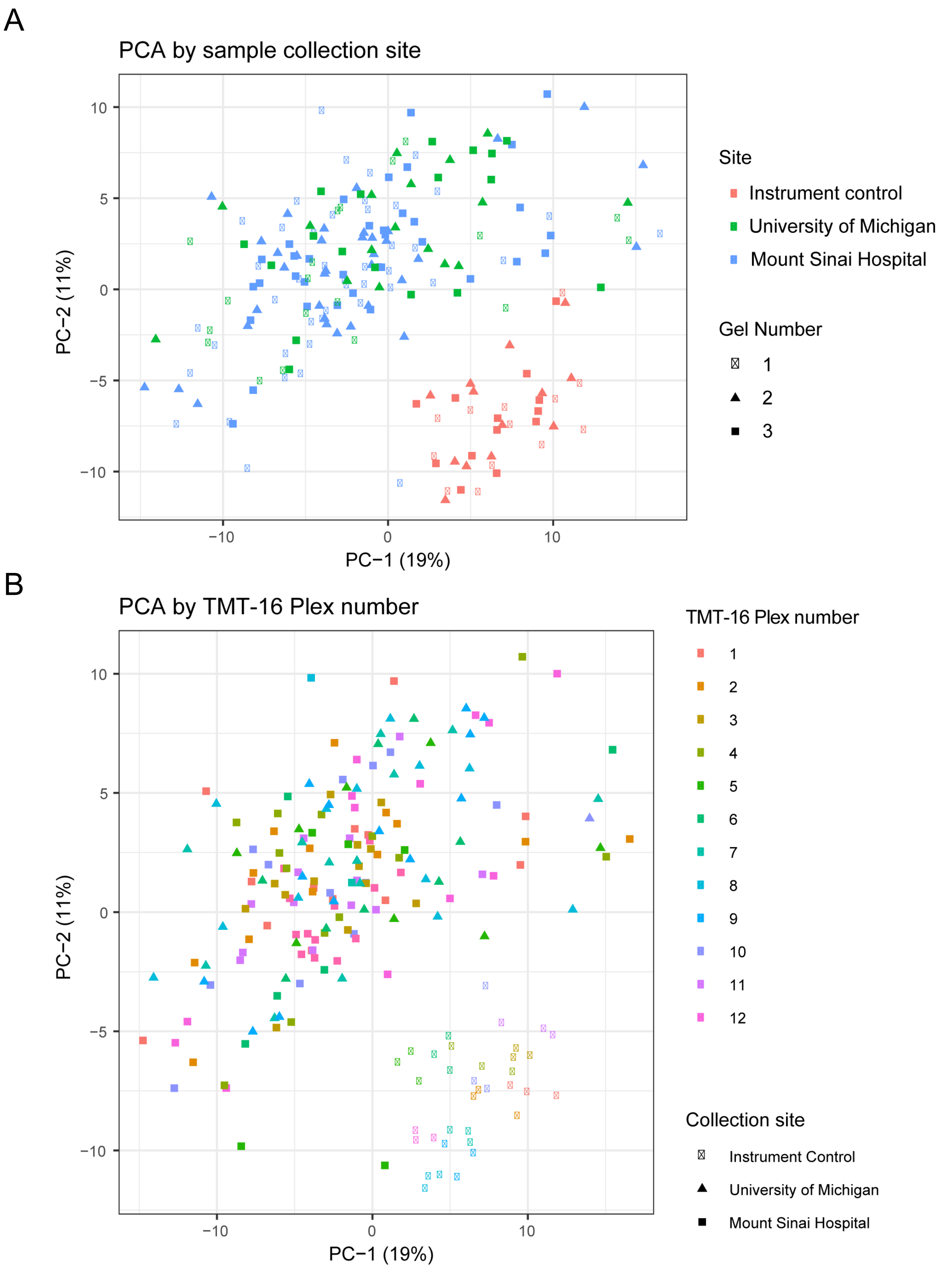

**Figure S3:** Gene Ontology biological processes (GOBP) enrichment analysis heatmap of top 150 (A) upregulated and (B) downregulated differentially abundant proteins (DAPs) in COVID-19 vs healthy urine samples

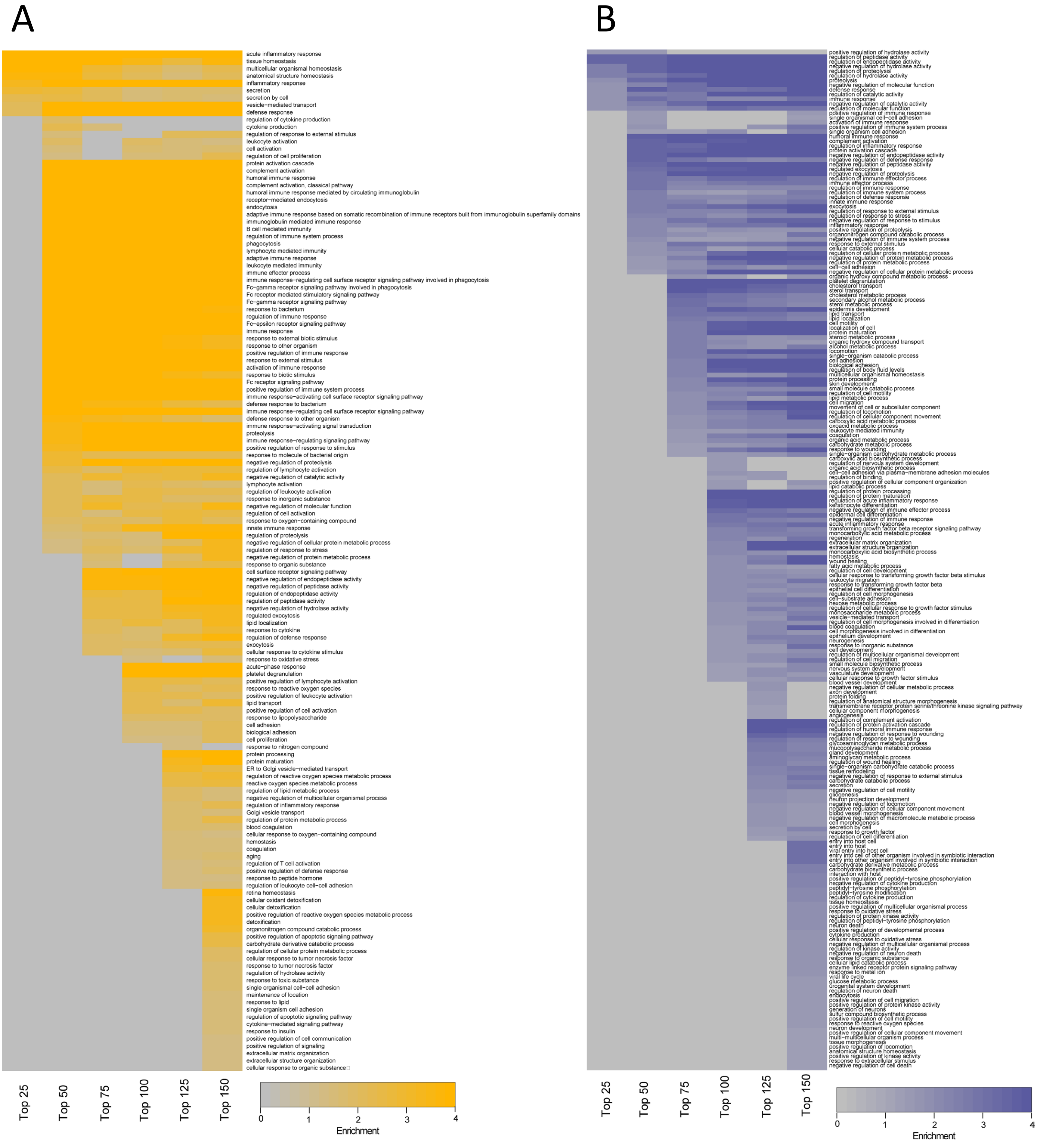

**Figure S4**: GOBP enrichment heatmap of top 150 (A) total and (B) downregulated DAPs in severe vs mild COVID-19 urine samples

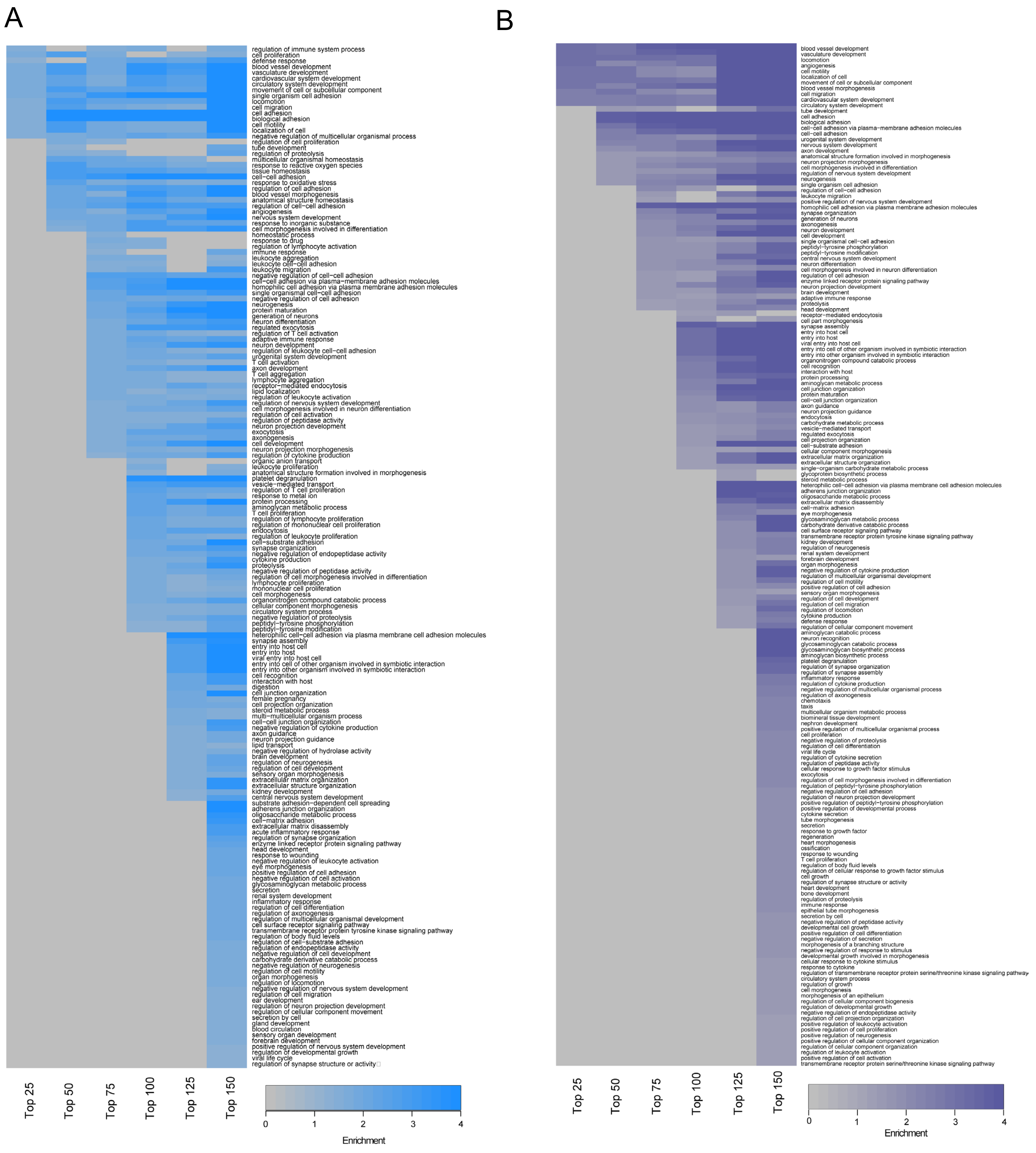

**Figure S5**: Correlation of prediction accuracy and (A) number of days hospitalized at urine collection; (B) outcome

A

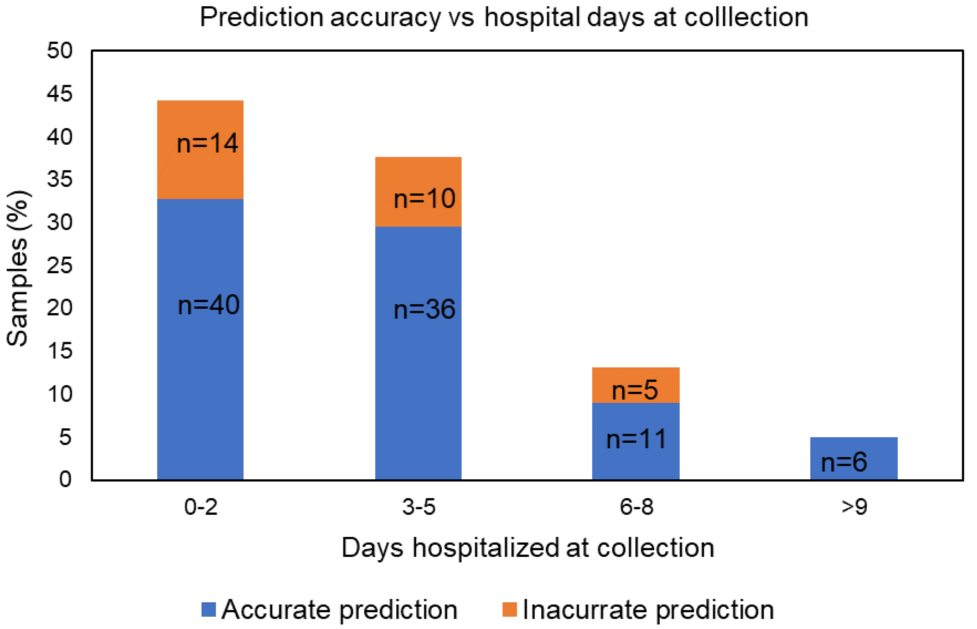


B

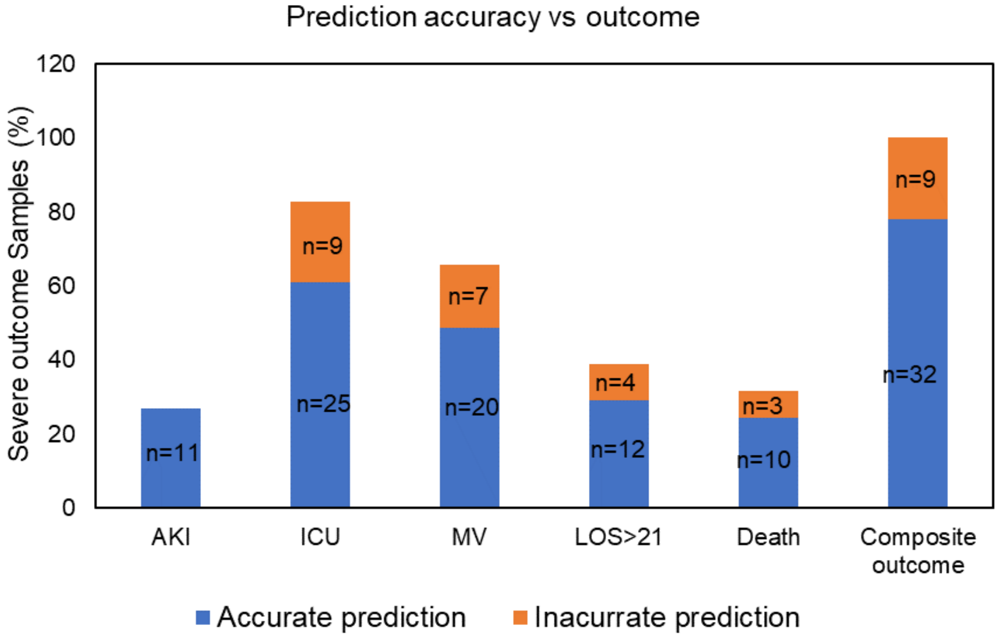

**Figure S6**: GOBP enrichment analysis of the 12 features from urine proteomics

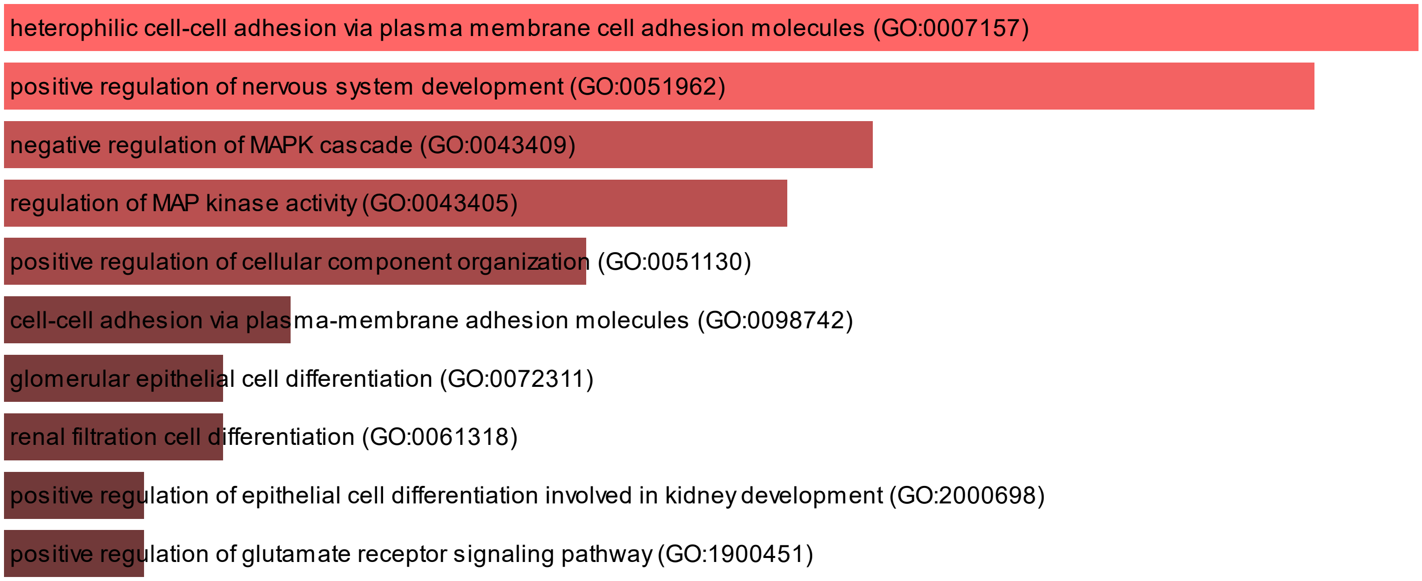

**Figure S7**: Kidney cell type specificity heatmap of ML features from the Kidney Precision Medicine Project (KPMP) database scRNA-seq dataset

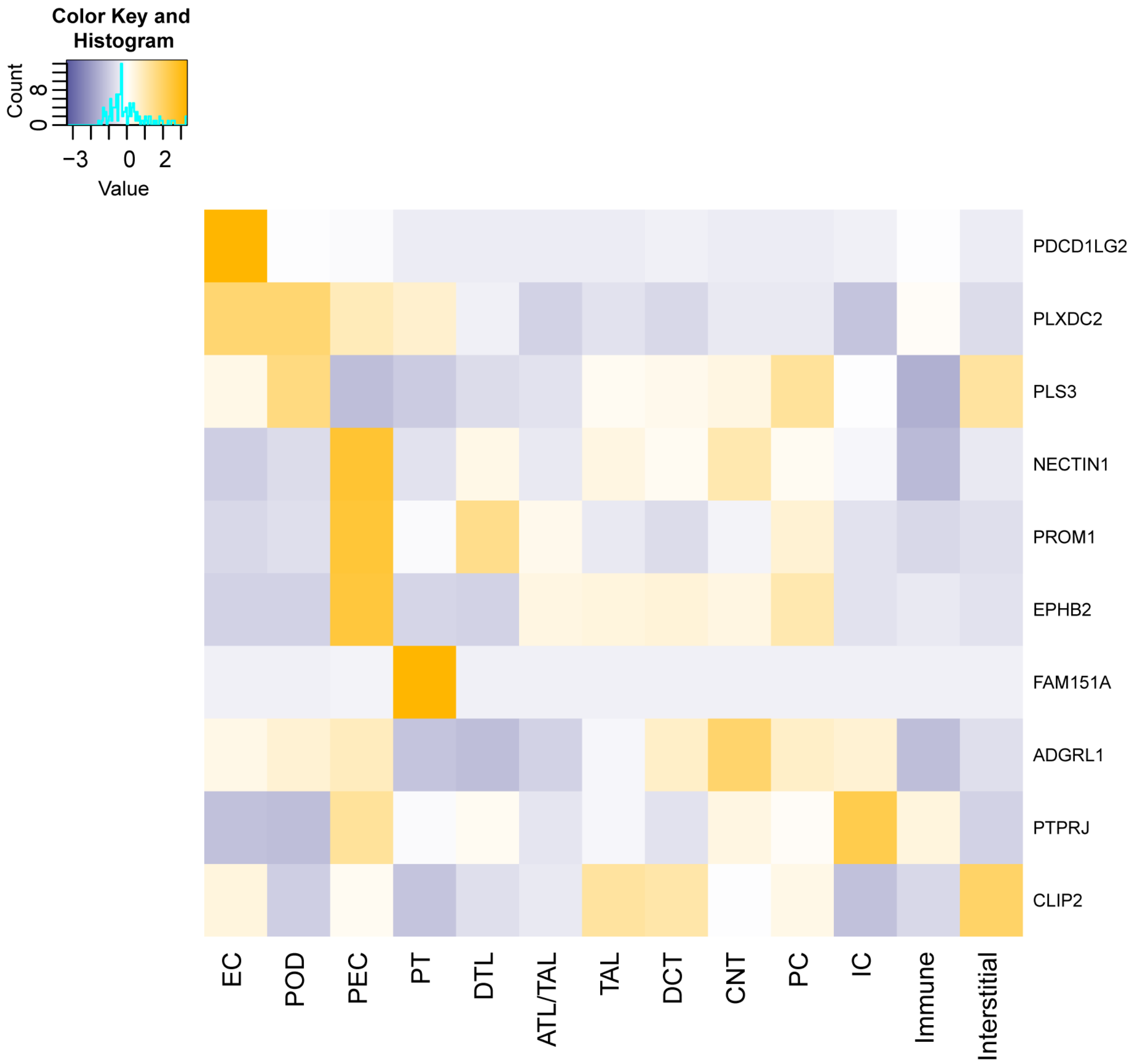

**Figure S8:** Estimated glomerular filtration rate (eGFR) in severe versus mild

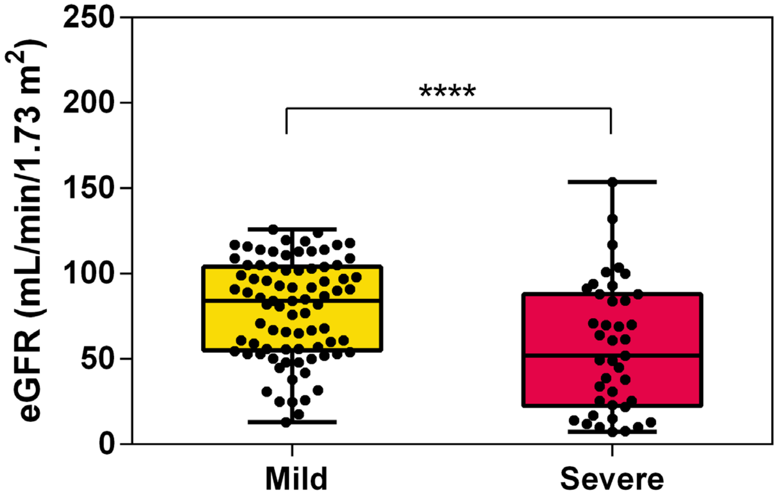

**Figure S9:** (A) Uniform Manifold Approximation and Projection (UMAP) on cosine distance metric; (B) K-means clustering of unscaled principal component analysis (PCA) plot; (C) Stacked bar graph of distribution of outcomes in each cluster

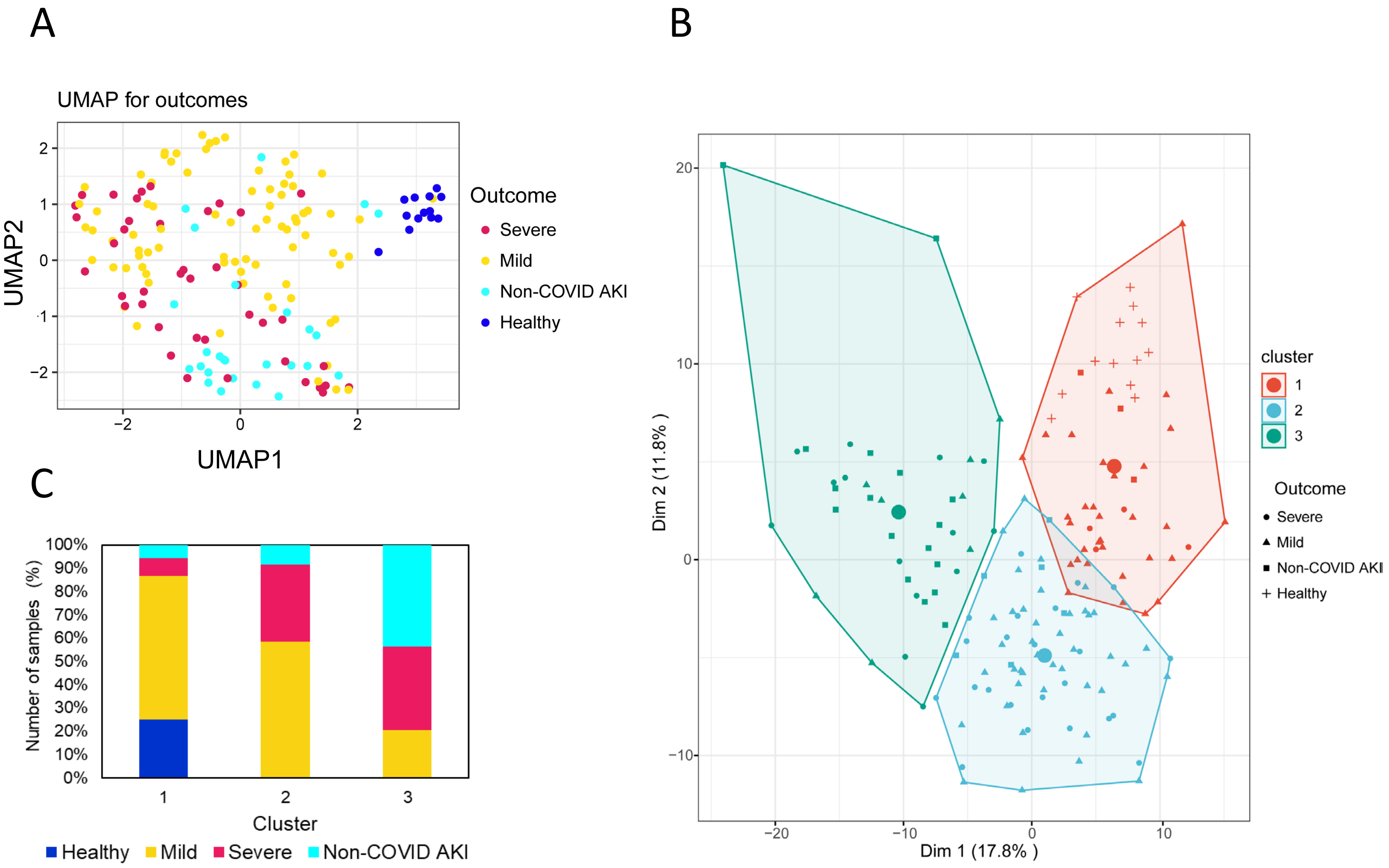

**Figure S10**: GOBP enrichment analysis heatmap of top 150 (A) upregulated and (B) downregulated DAPs in non-COVID AKI vs healthy urine samples

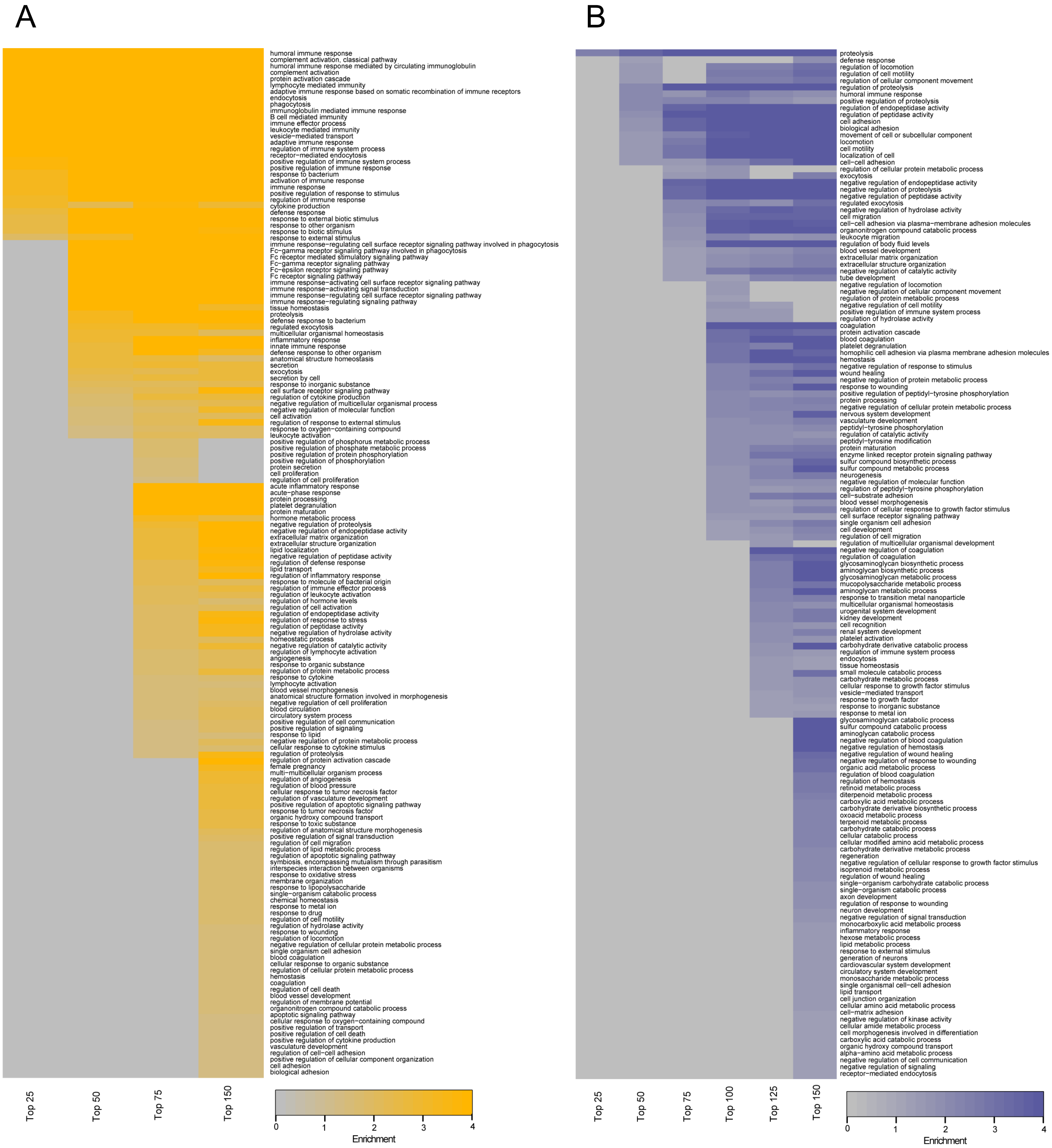

**Figure S11**: Abundance of (A) Albumin and (B) Cystatin C in healthy, mild and severe COVID-19 and non-COVID AKI urine samples

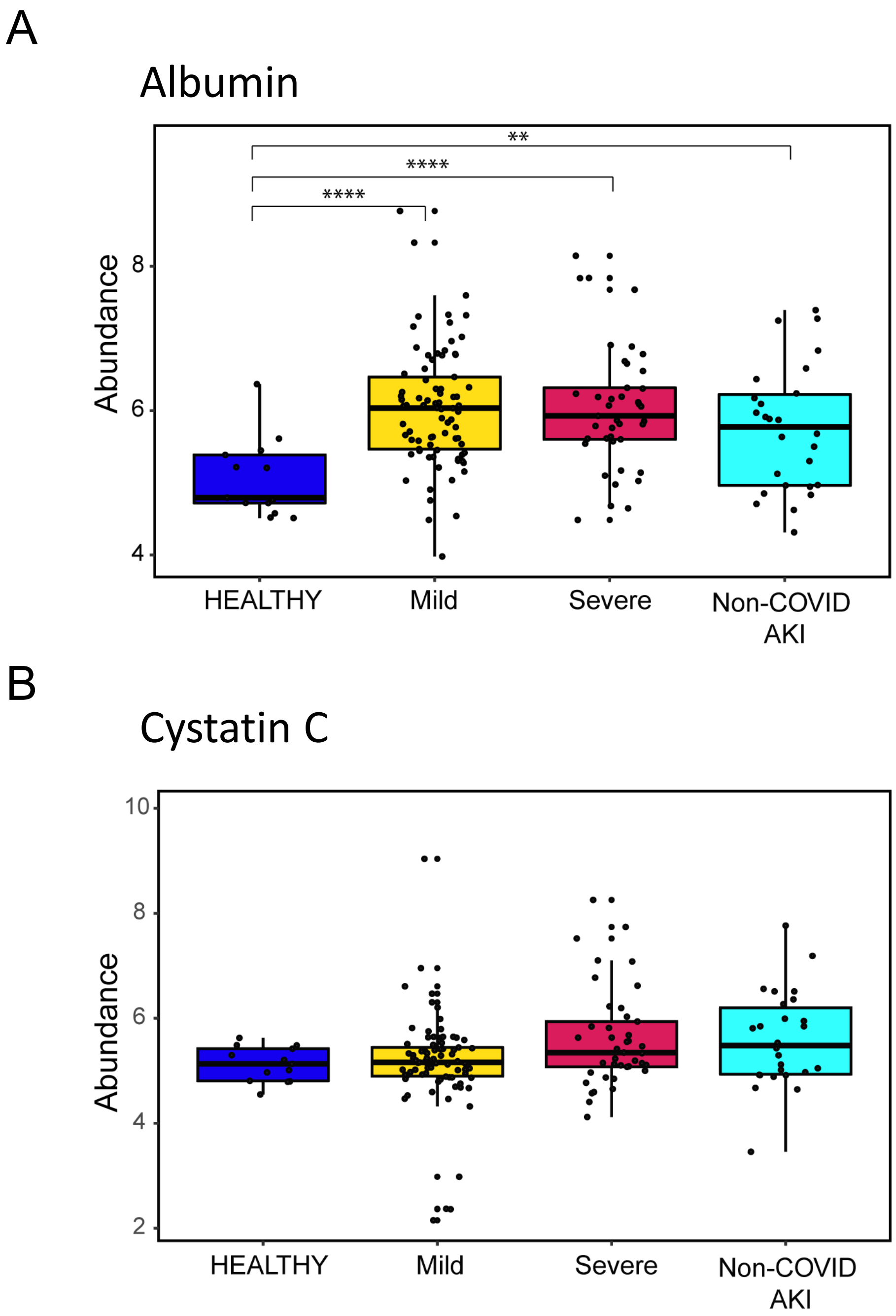

**Figure S12:** Log2 fold-change urinary abundance of ML features in AKI samples from an external dataset from the KPMP database

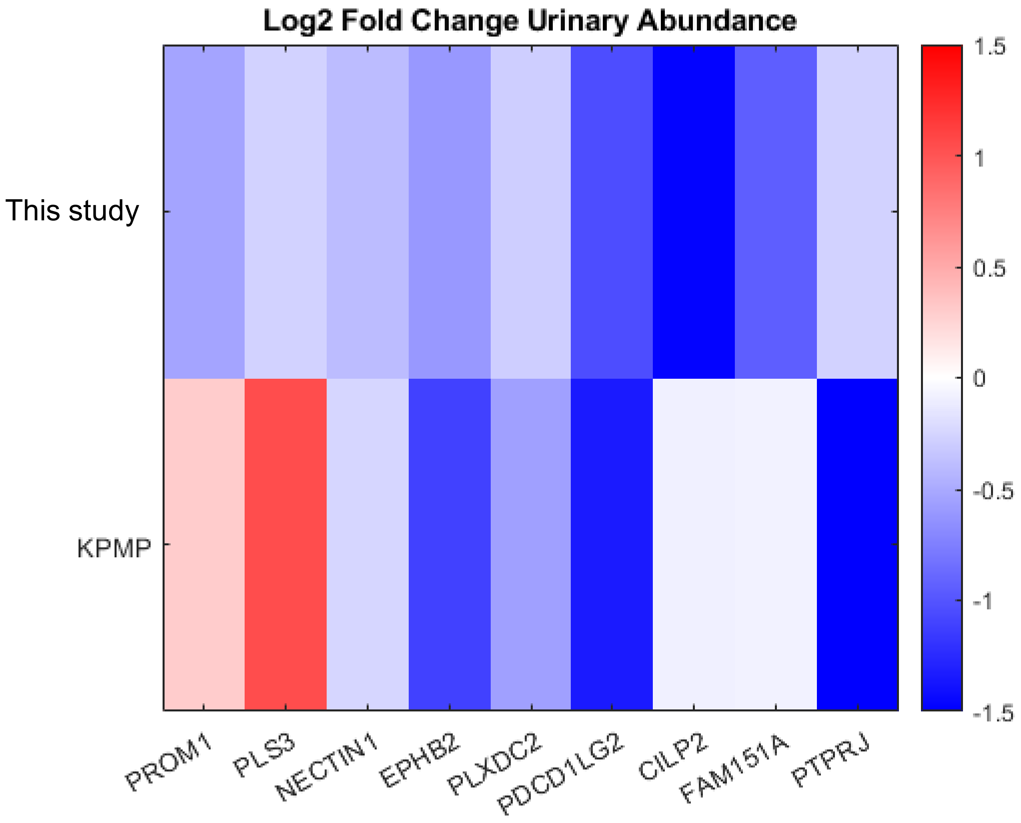

**Figure S13**: Interaction of top 150 urine and top 450 plasma proteomics DAPs analyzed using Molecular-Biology-of-the-Cell-Ontology (MBCO) standard enrichment. Pathways among top 25% interaction were connected by a dashed line

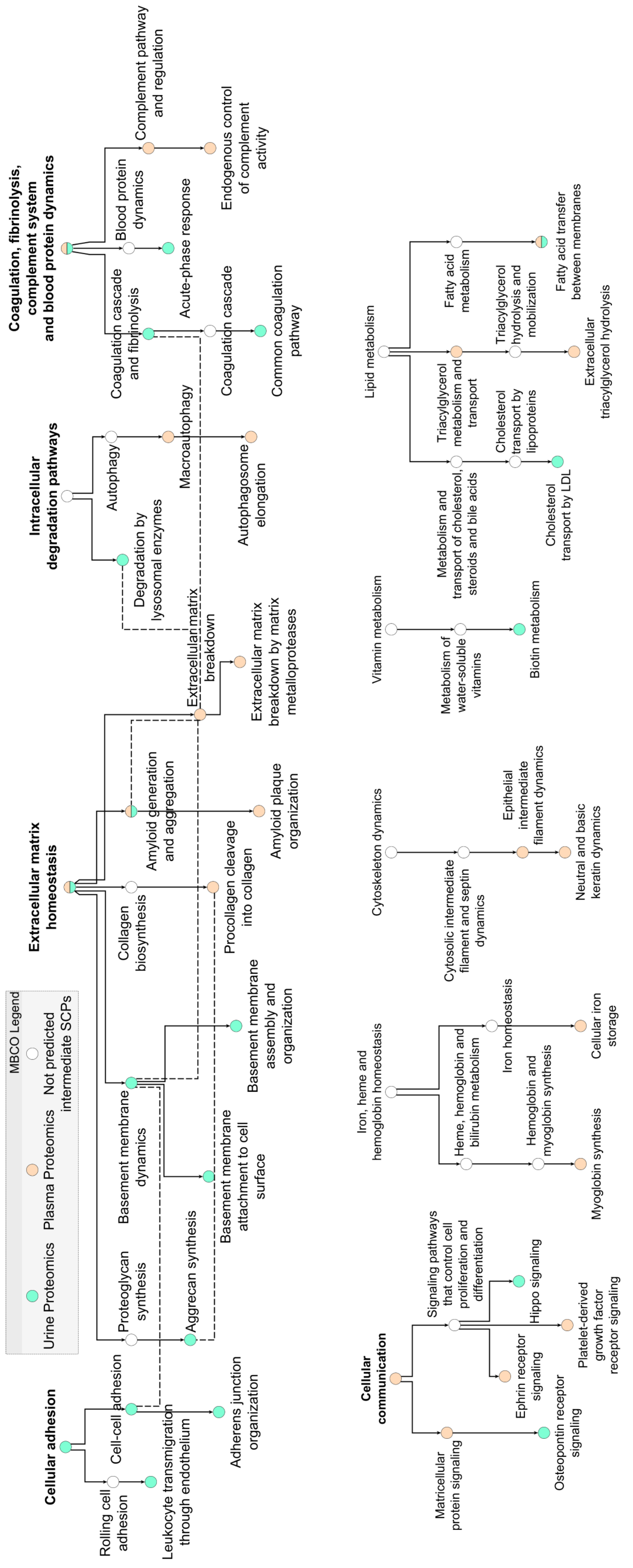

**Figure S14**: HumanBase kidney-specific functional module discovery analysis of top 150 DAPs from urine and 450 DAPs from plasma proteomics datasets

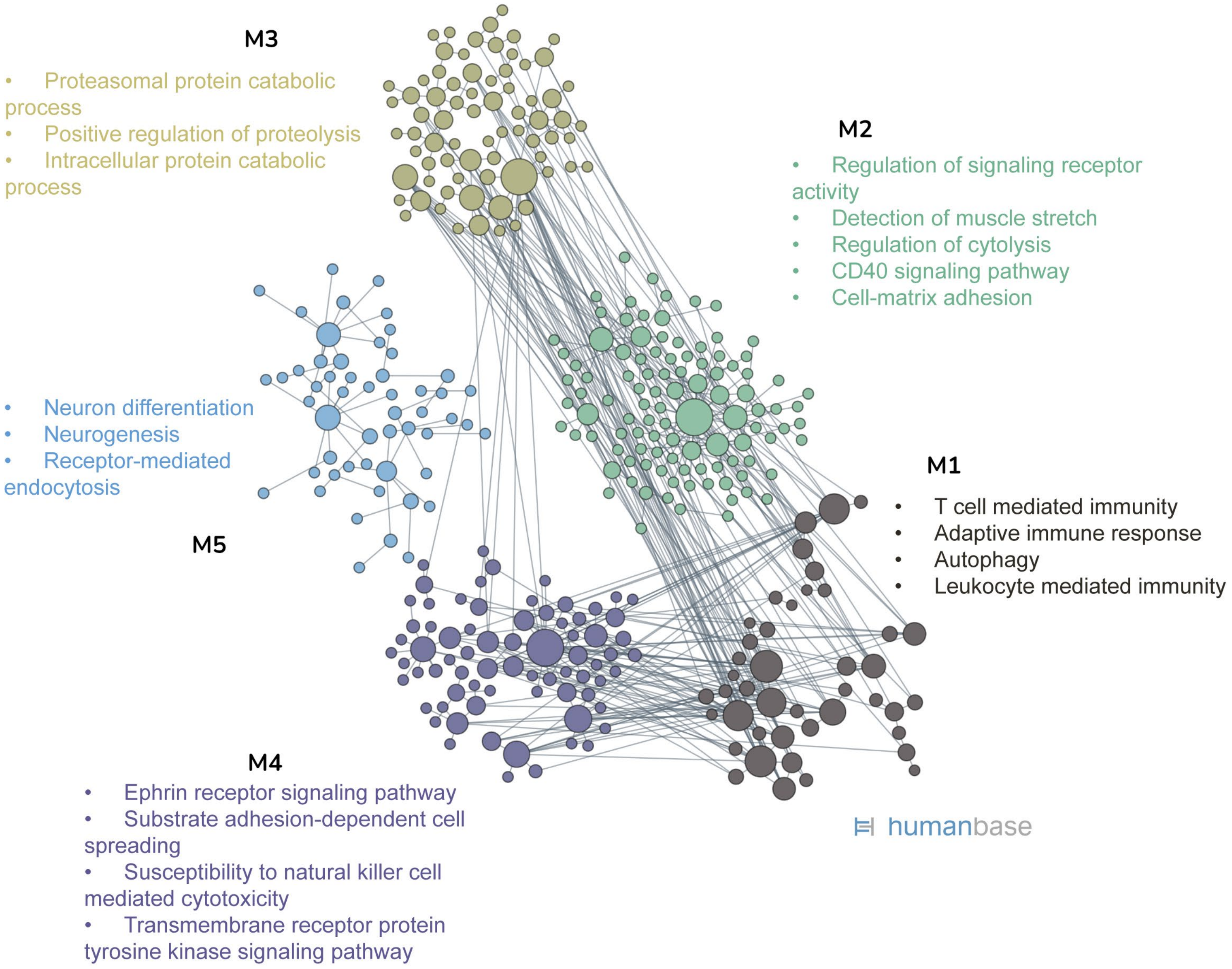

**Figure S15**: Kidney cell type specificity heatmap from scRNA-seq dataset in the KPMP database of (A) cell adhesion (B) cell migration and (C) response to oxidative stress associated genes from the functional overlap analysis

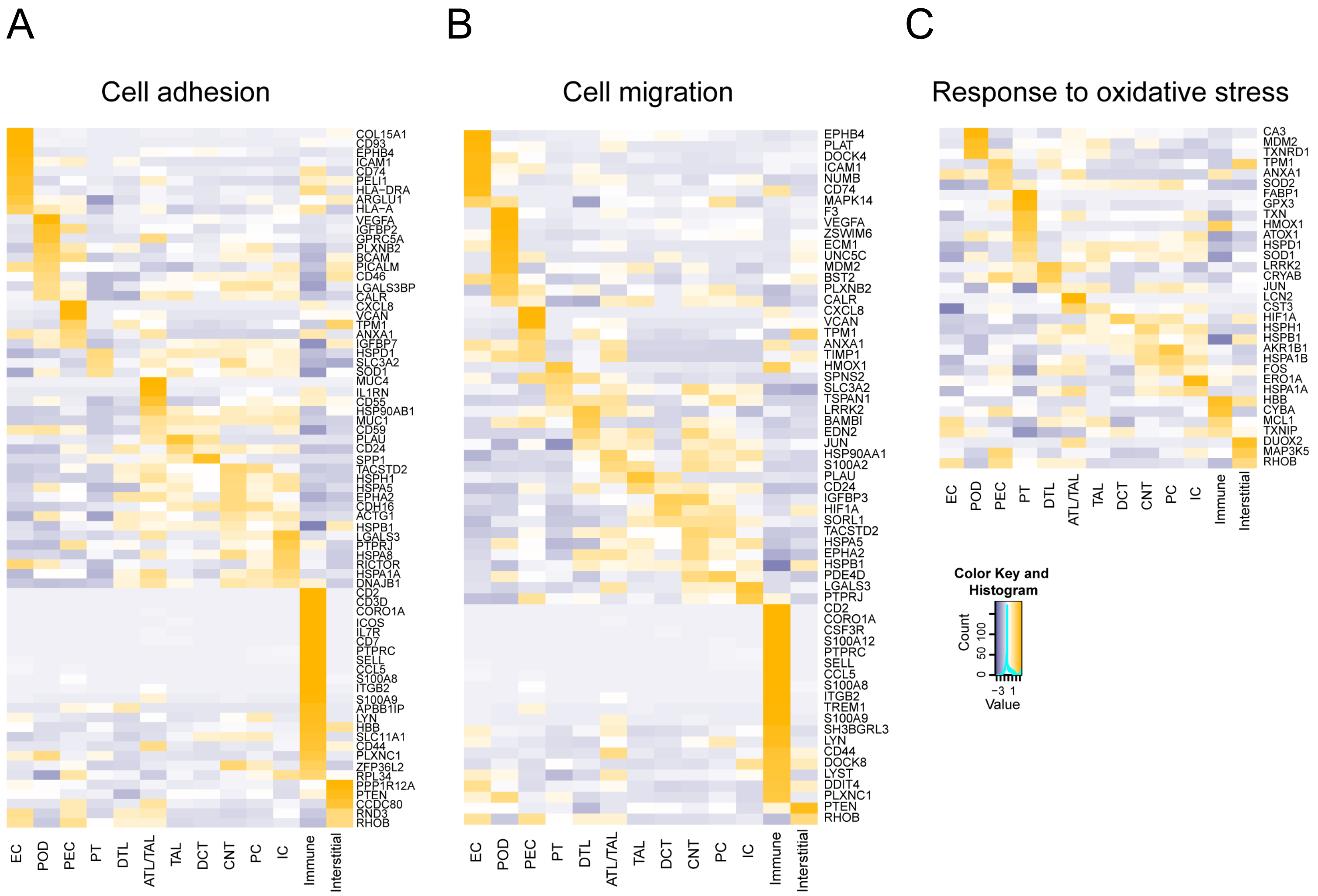

**Figure S16**: Histogram of the mean values of the permuted network scores along with original interaction score (red) of top 50 differentially expressed genes (DEGs) from individual cell clusters in the urine sediment single-cell RNA sequencing (scRNA-seq) and top 50 DAPs from urine and plasma

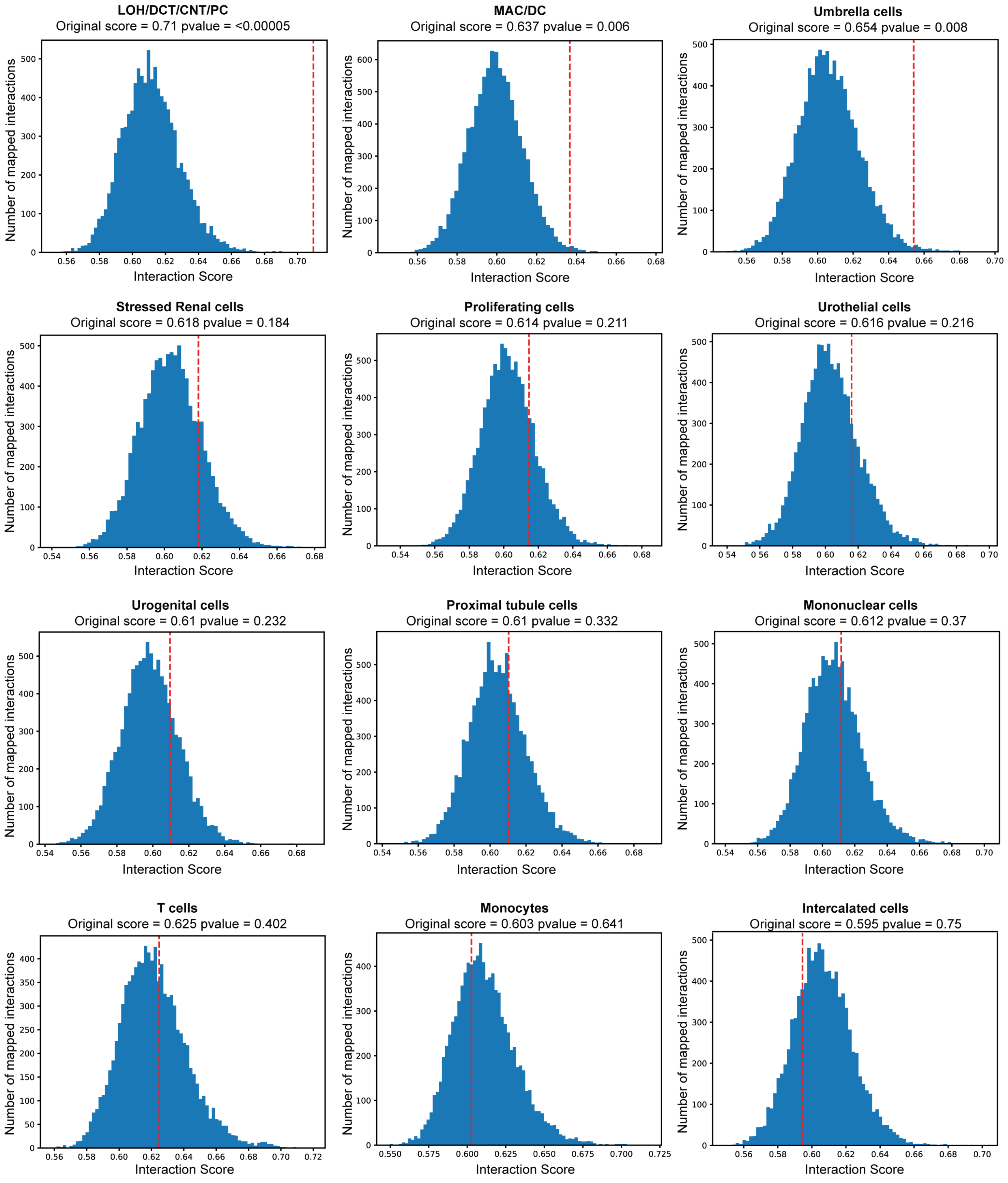

**Figure S17**: GOBP enrichment heatmap of DEGs in severe vs mild urine sediment scRNA-seq dataset LOH/DCT/CNT/PC cluster (hybrid cluster of loop of Henle, distal tubule, connecting tubule and principal cells)

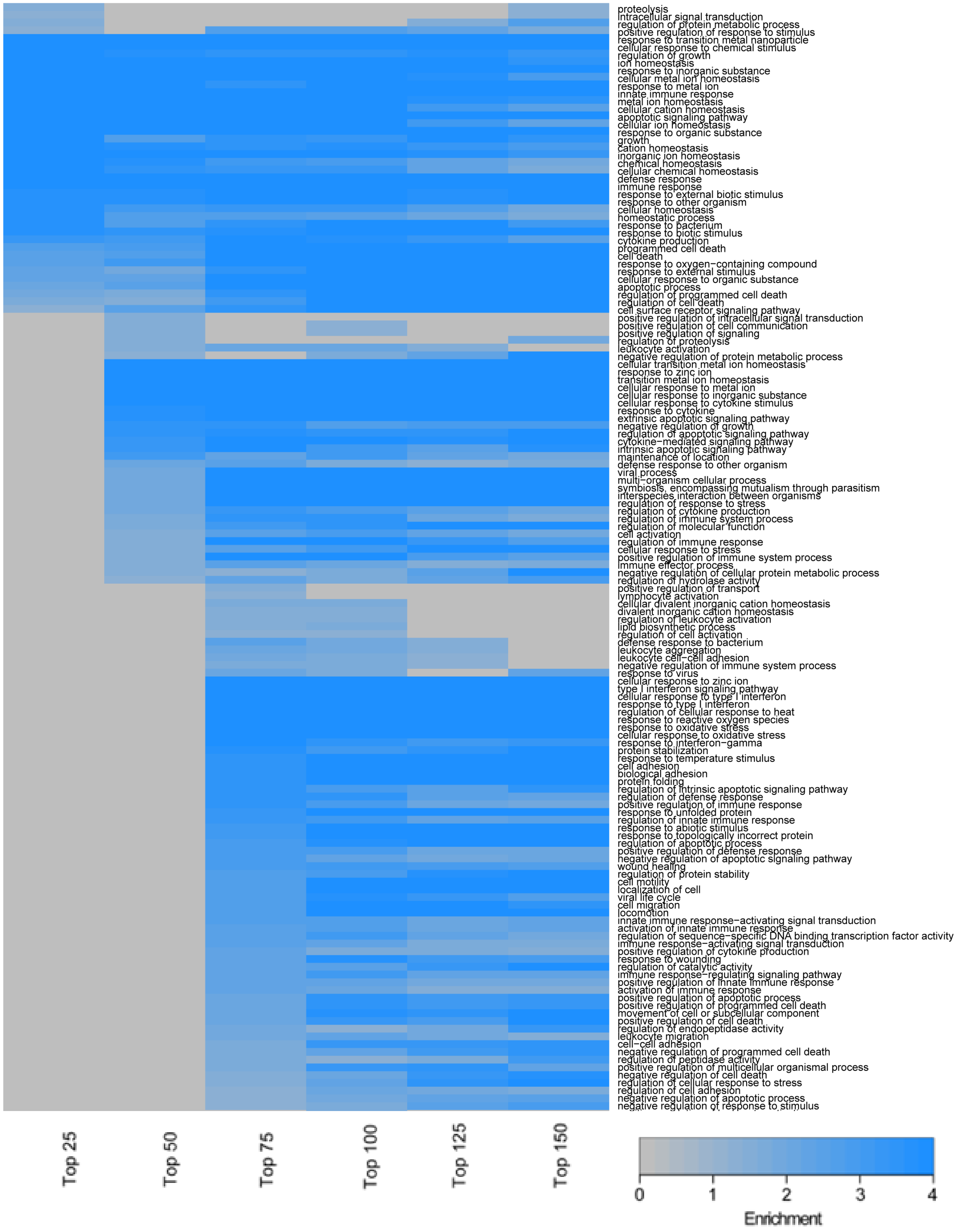

**Figure S18**: Interaction of top 150 DAPs from Urine and top 450 DAPs from plasma proteomics with top 150 DEGs from the Urine sediment scRNA-seq LOH cluster analyzed using MBCO standard enrichment. Pathways among top 25% interaction were connected by a dashed line

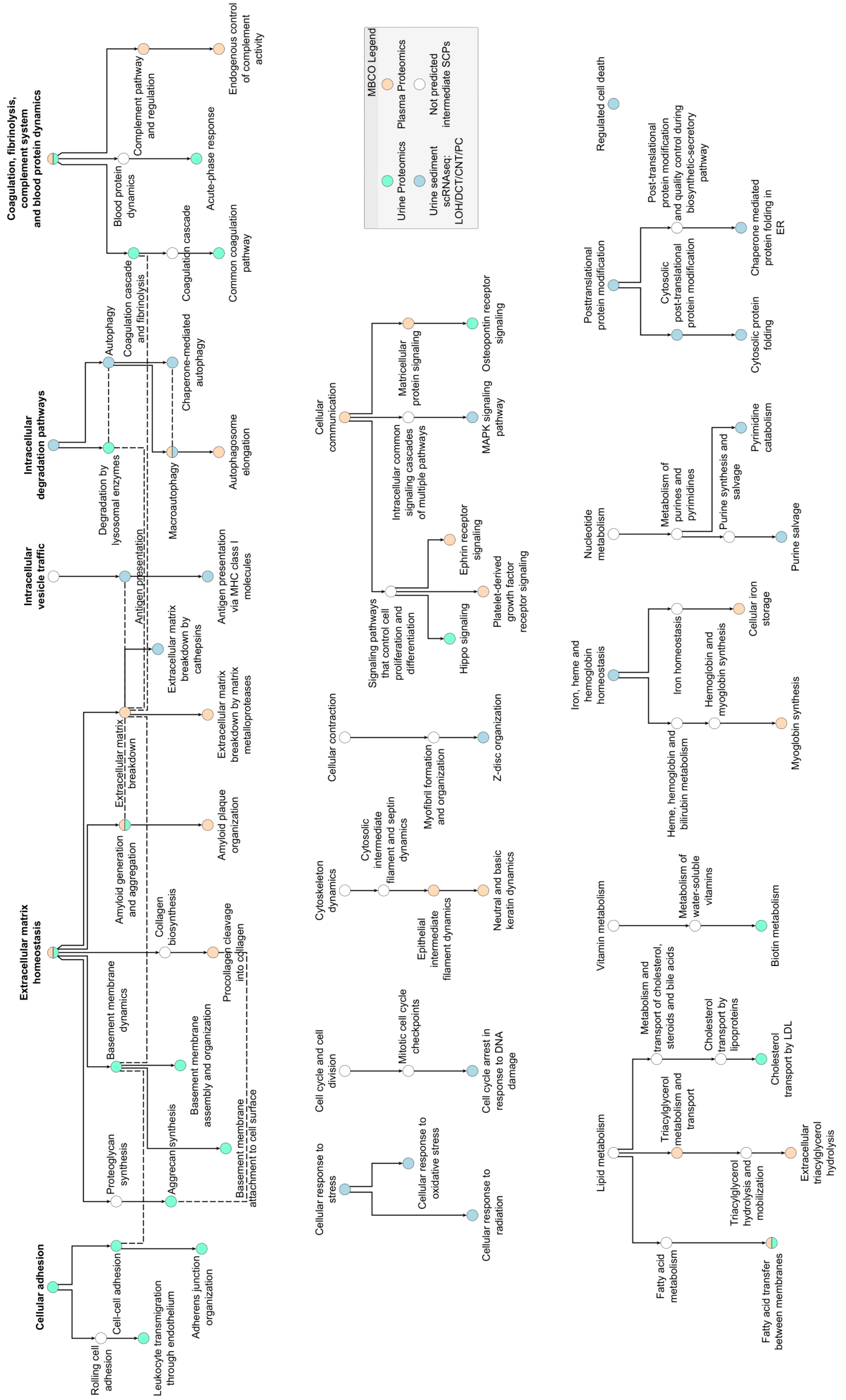

**Figure S19**: HumanBase kidney-specific functional module discovery analysis of top 150 DAPs from urine and top 450 from plasma proteomics and top 150 DEGs from the urine sediment scRNA-seq LOH/DCT/CNT/PC cluster

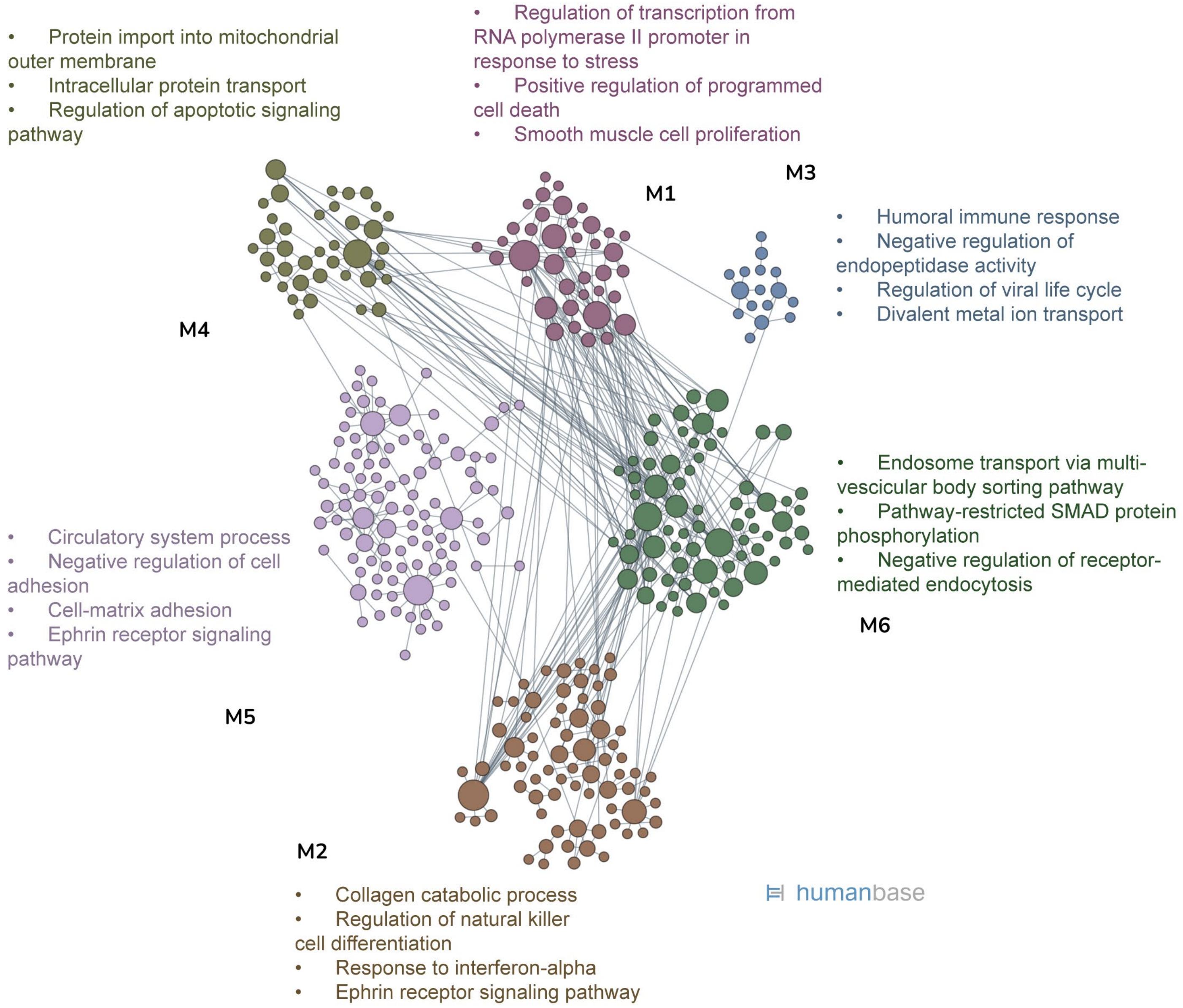

**Figure S20**: HumanBase kidney-specific functional module discovery analysis of overlapping proteins from the protein-protein interaction (PPI) network of urine and plasma proteomics datasets and urine scRNAseq LOH/DCT/CNT/PC cluster

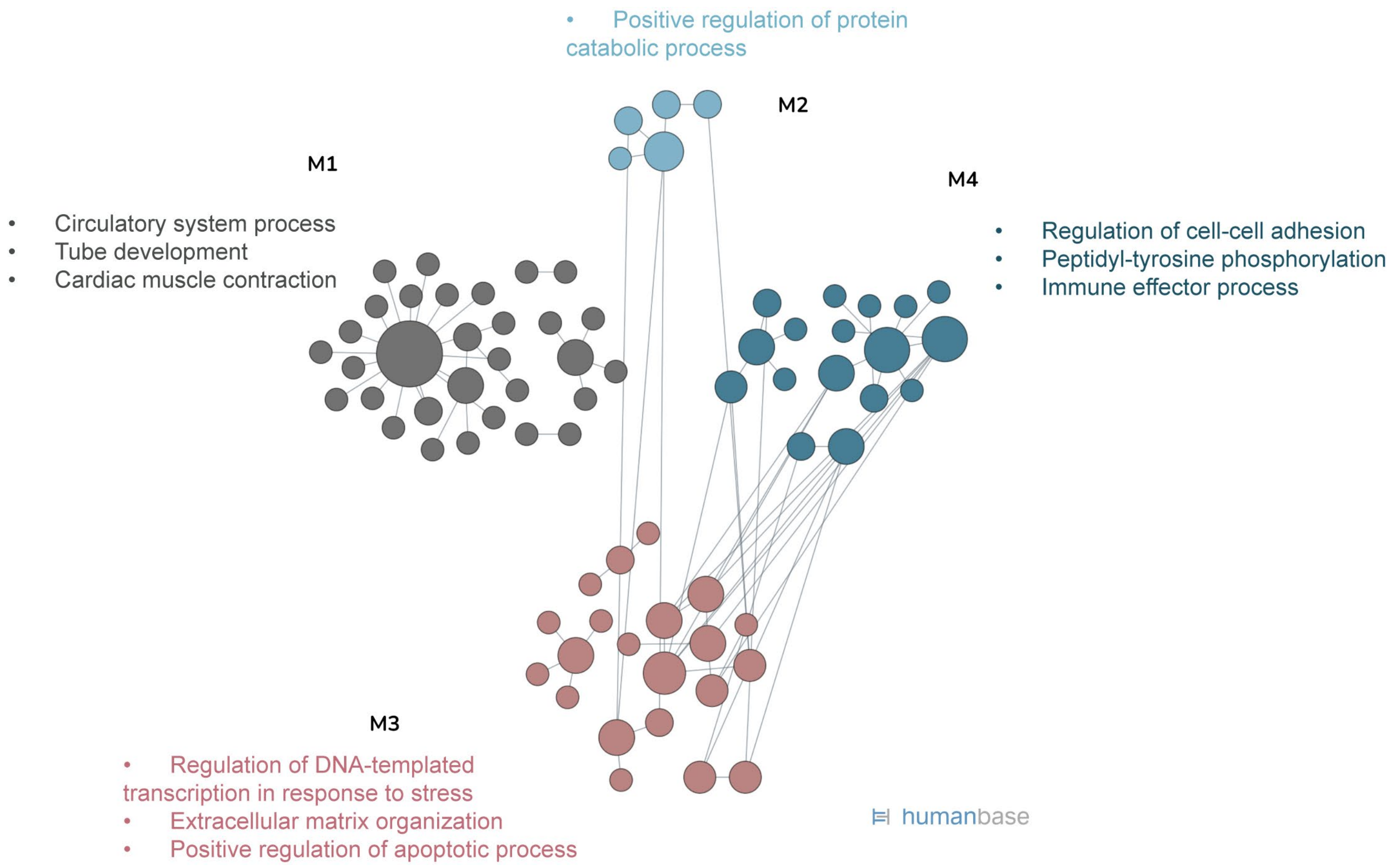

**Figure S21:** Viral titer quantification using supernatants from infected organoids on 2-, 4- and 6-days post infection (dpi)

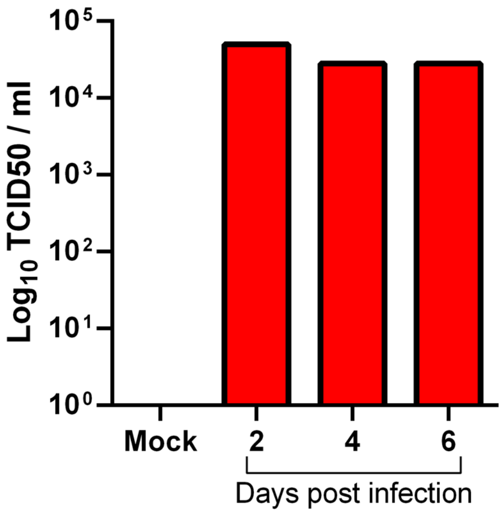

**Figure S22**: Cell type clusters identified from the kidney organoid scRNA-seq

A

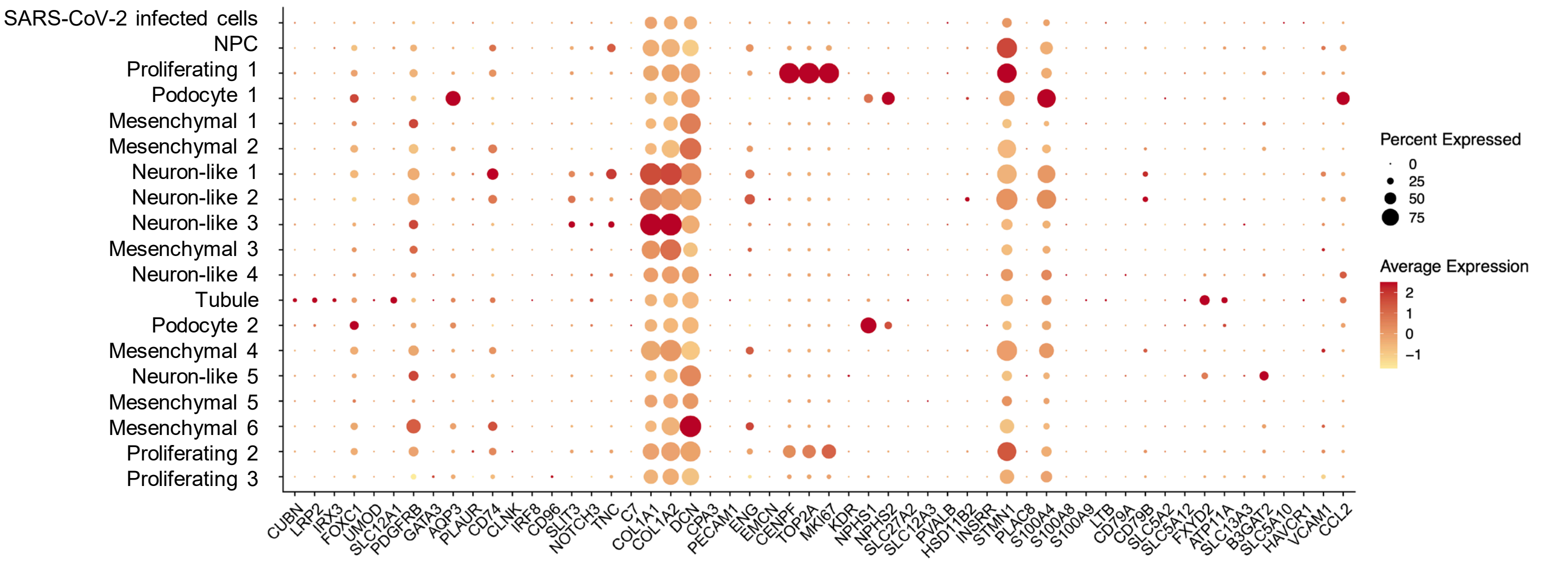


B

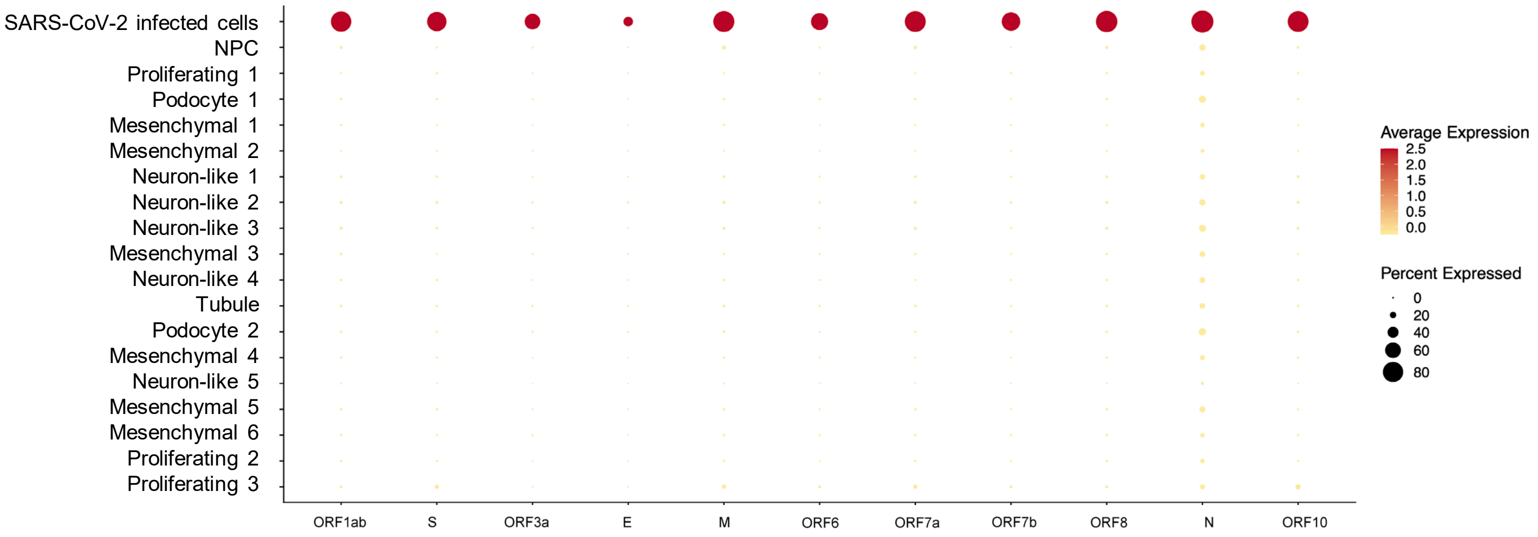

**Figure S23**: GOBP enrichment heatmap of top 150 DEGs in infected vs mock kidney organoids

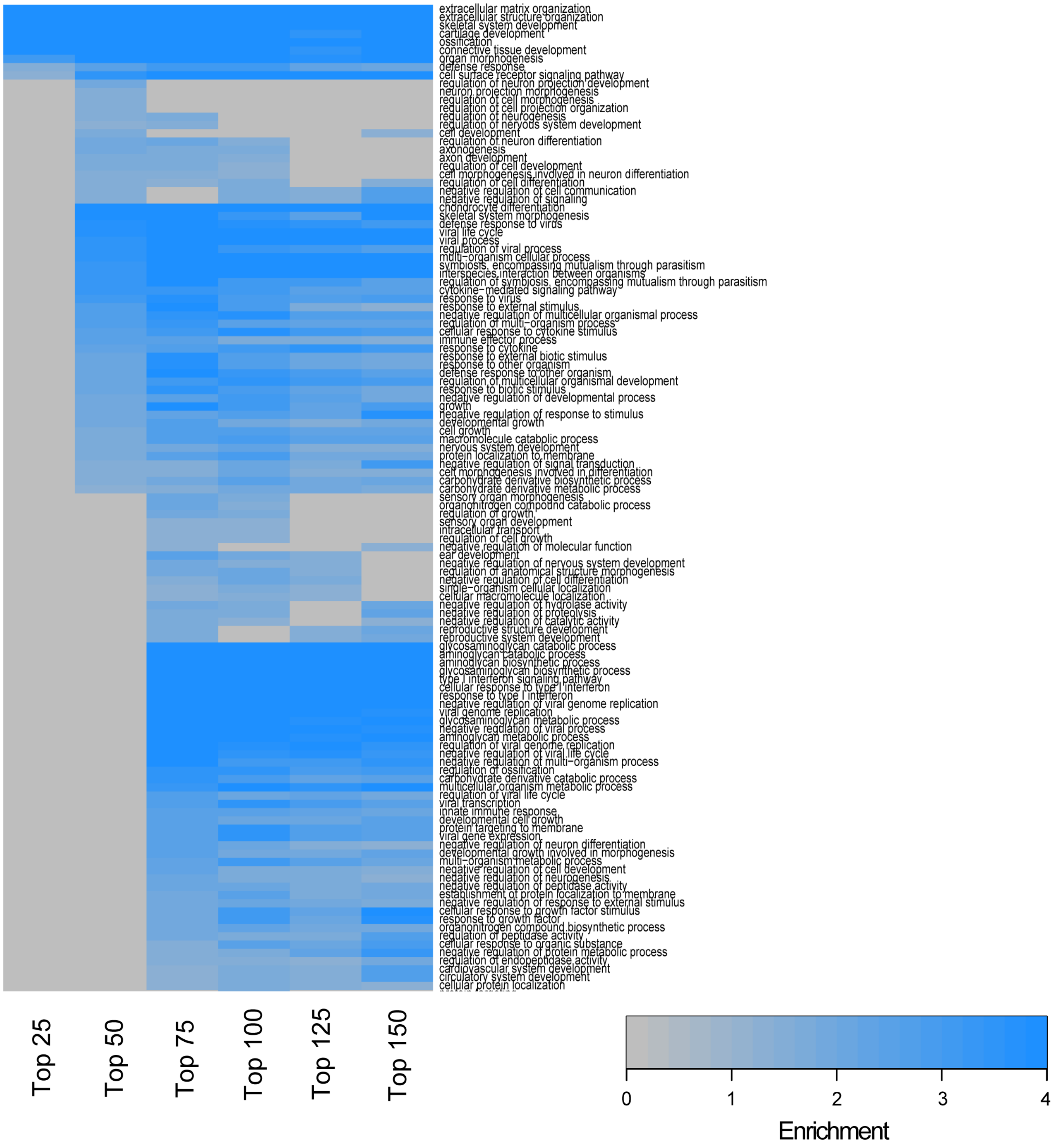

**Figure S24**: Interaction of top 150 DEGs from kidney organoid scRNAseq and the top 150 DAPs from urine and the top 450 DAPs from plasma proteomics datasets analyzed using MBCO standard enrichment. Pathways among the top 25% interaction were connected by a dashed line

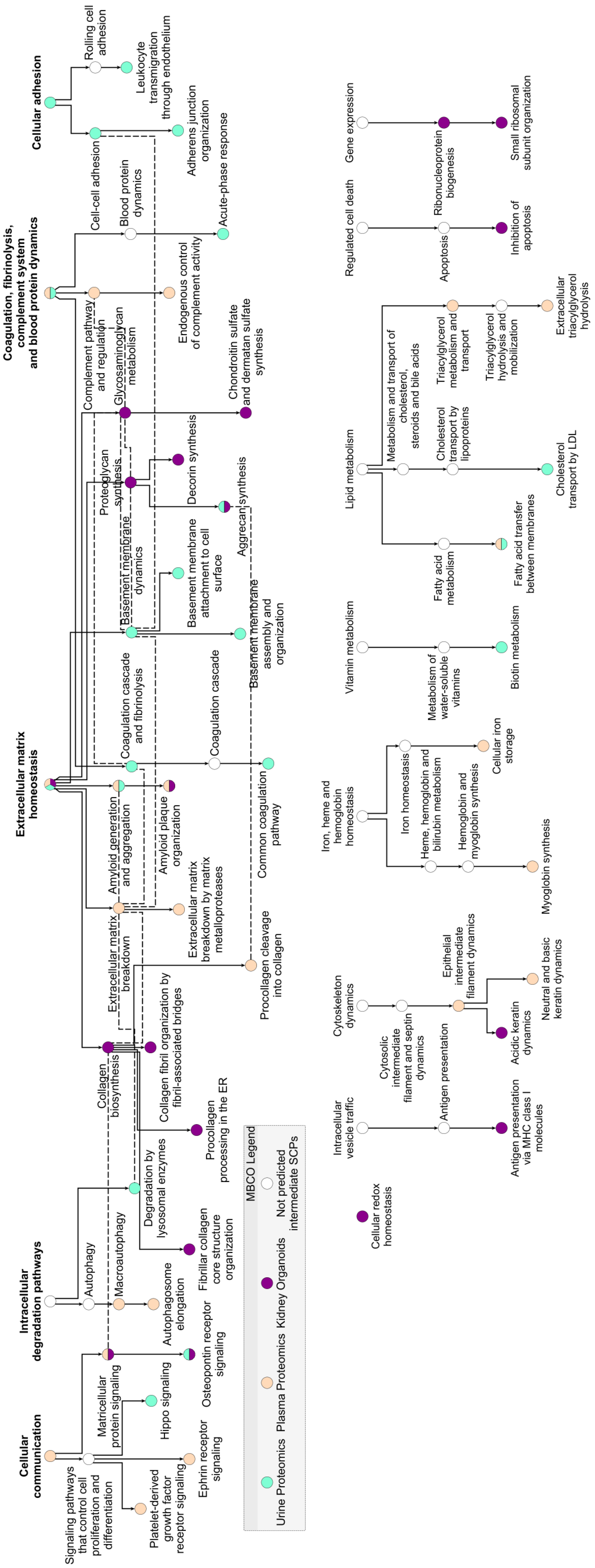

**Figure S25**: HumanBase kidney-specific functional module discovery analysis of top 150 DAPs from urine and plasma proteomics and top 150 DEGs from the kidney organoid scRNAseq dataset

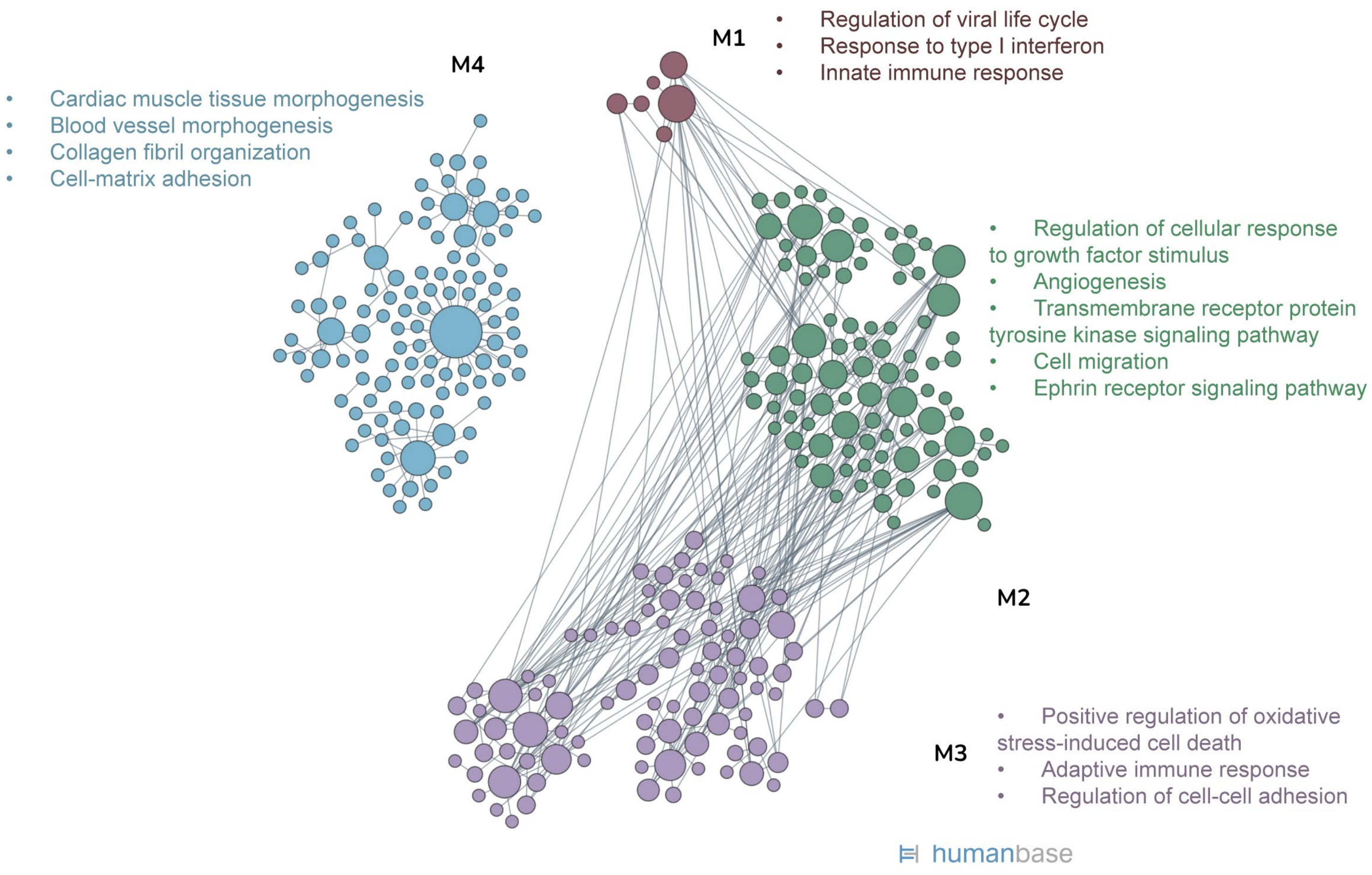

**Figure S26**: Abundance of TMPRSS2 in mild and severe COVID-19 samples

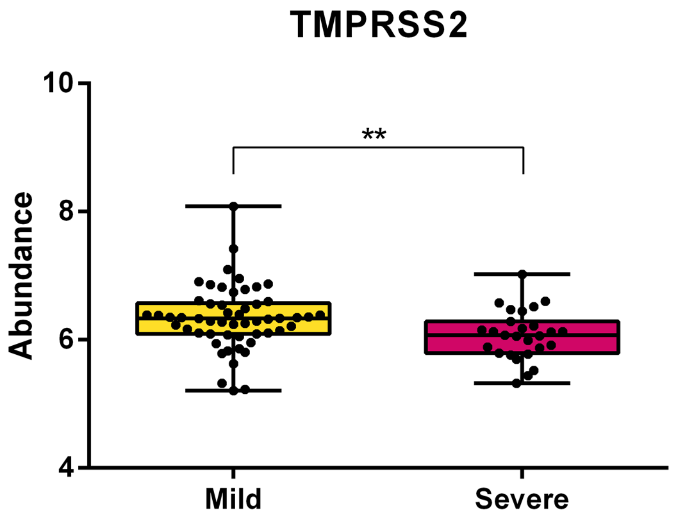

**Figure S27:** Receiver operating characteristics (ROC) curves of COVID-19 severity prediction using Mount Sinai Hospital cohort as (A) discovery set and University of Michigan cohort as (B) validation set

A

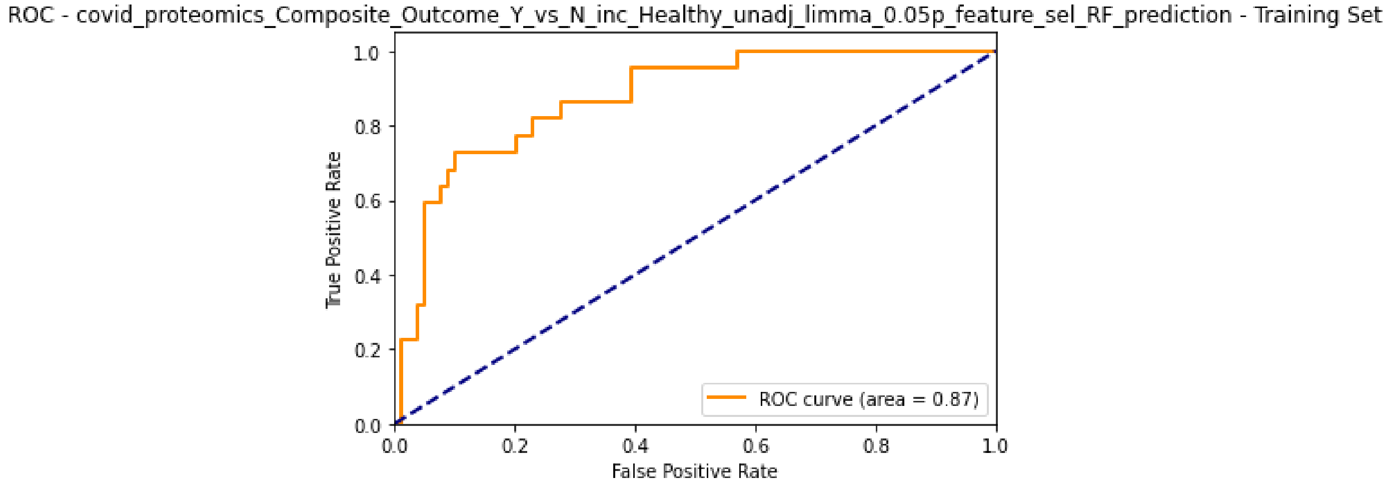


B

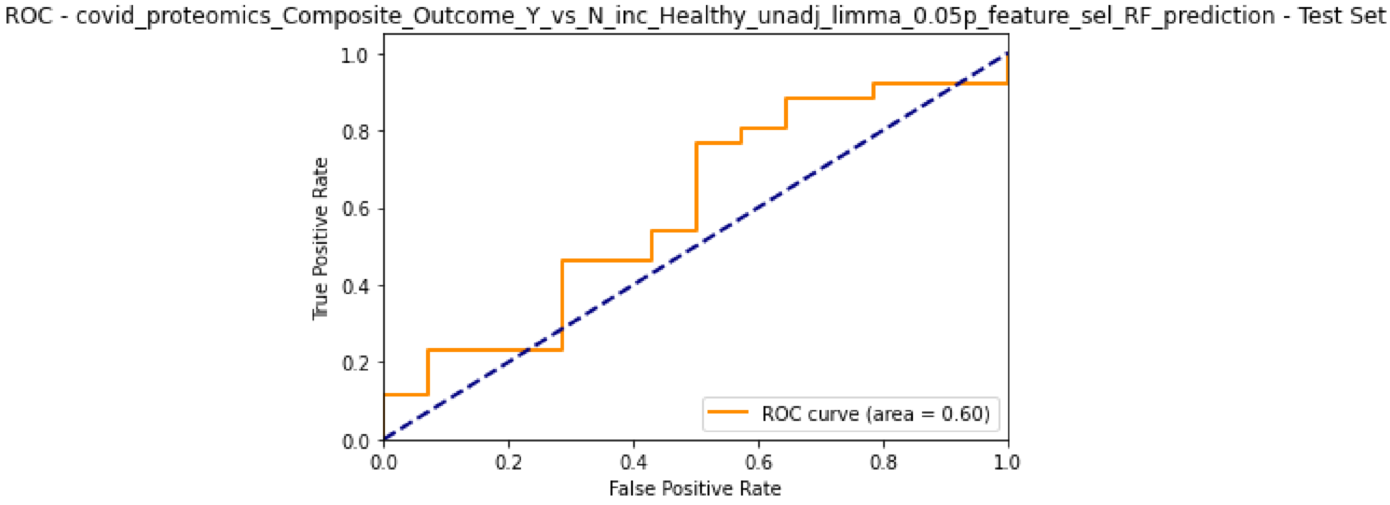

**Figure S28:** (A) List of ML features identified and (B) GOBP enrichment analysis of biomarkers from ML model using Sinai cohort as discovery set and Michigan cohort as validation set

A

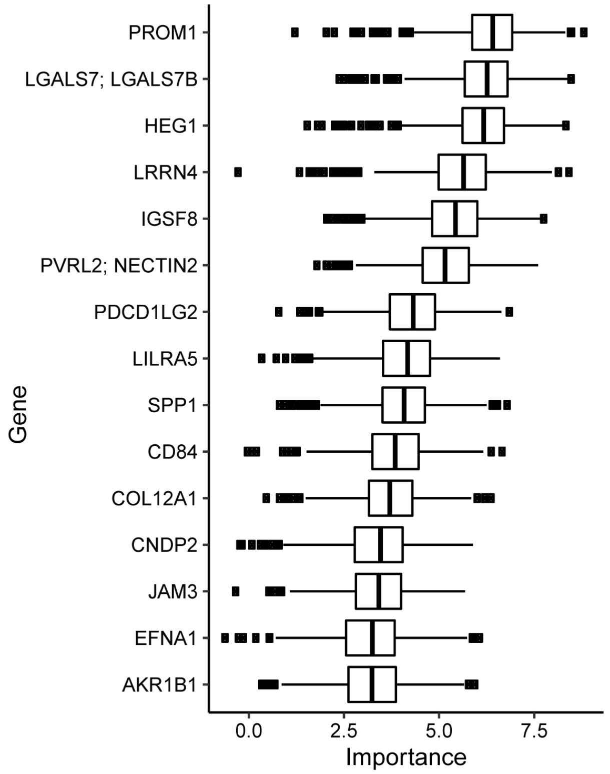


B

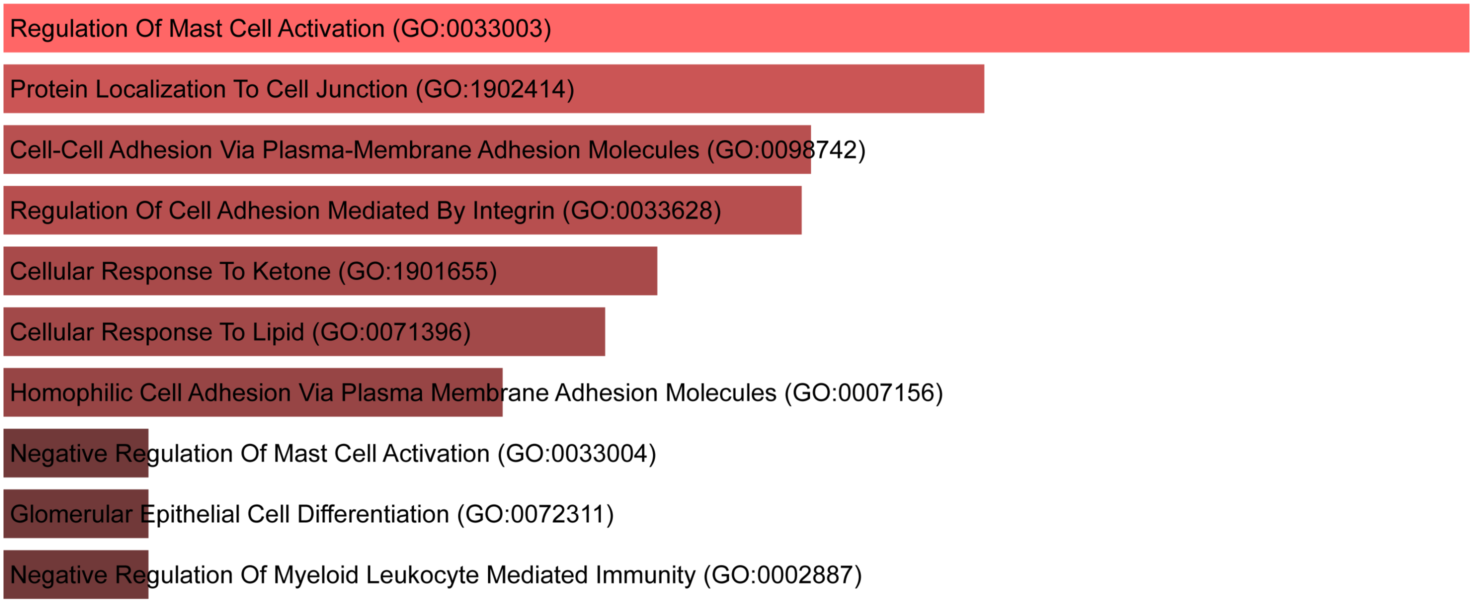

**Figure S29:** Schematic showing urine sample processing workflow for urinary proteomics

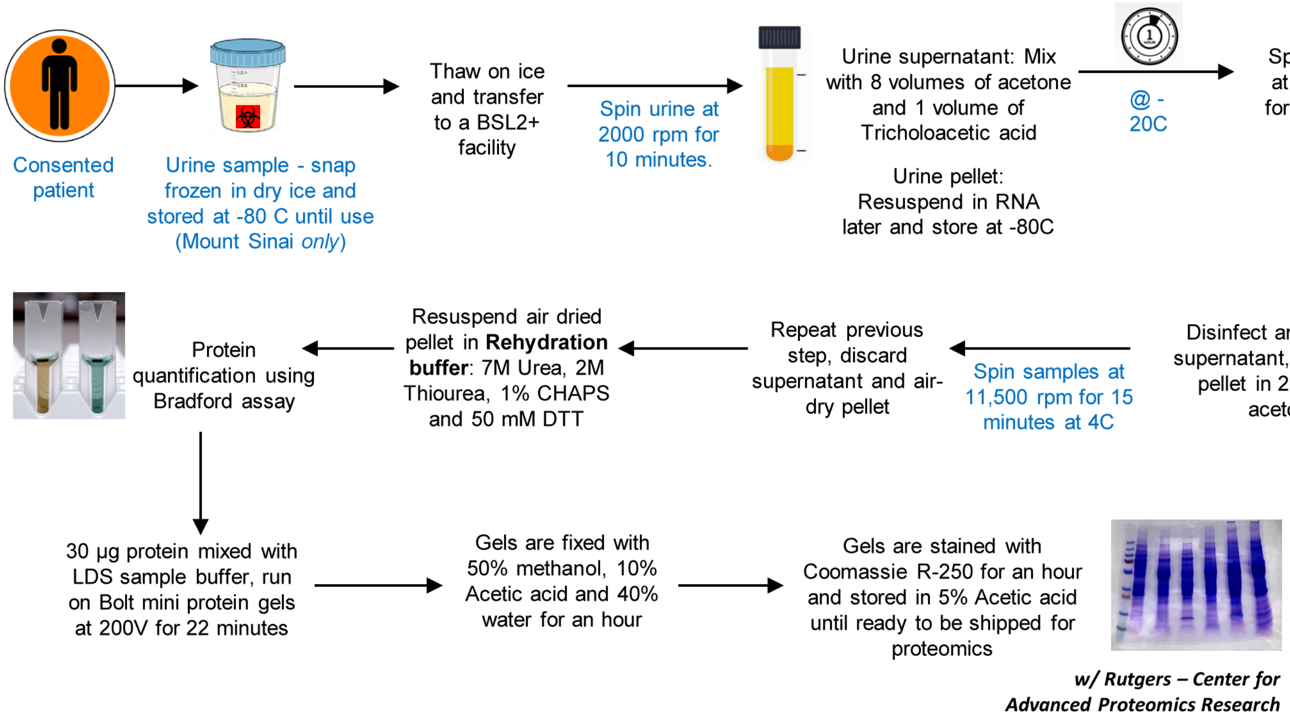

**Figure S30:** Schematic of iPSC-derived kidney organoid generation

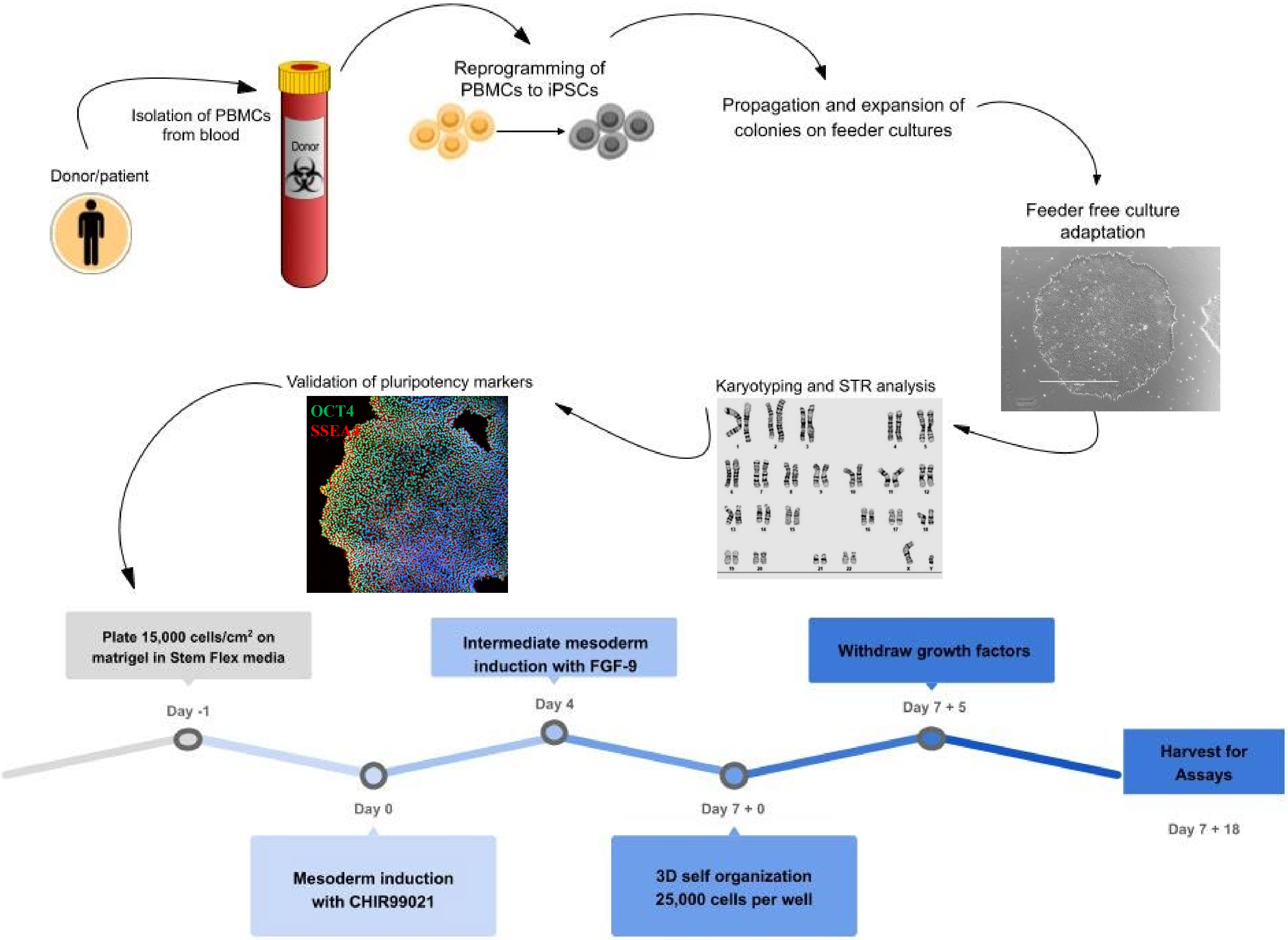
